# Exploring the Relationship between Caffeine Consumption, Caffeine Metabolism, and Sleep Behaviours: A Mendelian Randomisation Study

**DOI:** 10.1101/2025.04.30.25326725

**Authors:** Nilabhra R. Das, Benjamin Woolf, Stephanie Page, Rebecca C. Richmond, Jasmine Khouja

**Affiliations:** Population Health Sciences, University of Bristol, Bristol, UK; School of Psychological Sciences, University of Bristol, Bristol, UK; MRC Integrative Epidemiology Unit at the University of Bristol, Bristol, UK; MRC Biostatistics Unit at the University of Cambridge, Cambridge, UK; NIHR Oxford Health Biomedical Research Centre, University of Oxford, Oxford, UK

**Keywords:** Mendelian randomization, sleep, genetics, caffeine, metabolites

## Abstract

Higher consumption of caffeinated beverages is associated with disturbed sleep patterns. Using genetic variants as proxies for caffeine consumption, caffeine metabolism, and sleep traits, we investigated whether this association reflects a direct effect of caffeine. Genetic variants associated with caffeine consumption (n=407,072), caffeine metabolism (n=9,876), chronotype (n=449,734), daytime napping (n=452,633), daytime sleepiness (n=452,071), getting up in morning (n=385,949), insomnia (n=453,379), and sleep duration (n=446,118) identified in individuals from several studies, including the UK Biobank, were used to explore bi-directional causal relationship between caffeine and sleep using a series of univariable Mendelian Randomisation analyses. We used multivariable Mendelian Randomisation to explore the direct effects of caffeine consumption on sleep behaviours while adjusting for metabolism and vice-versa. Higher consumption decreased daytime sleepiness (β_univariable_=-0.044, 95%CI [-0.065,-0.023], p<0.001; β_multivariable_=-0.034, 95%CI [- 0.058,-0.009], p=0.010), while faster caffeine metabolism, indicative of less caffeine exposure per beverage consumed, decreased the likelihood of daytime napping (β_univariable_=-0.024, 95%CI [-0.037,-0.011], p<0.001; β_multivariable_=-0.021, 95%CI [-0.042,0.000], p=0.051). Being an evening person decreased caffeine consumption (β_univariable_=-0.044, 95%CI [-0.078,-0.010], p=0.010). Caffeine consumption/metabolism was not causal ly related to sleep duration or insomnia. We found no clear evidence for effects of caffeine consumption/metabolism on sleep among non-current caffeine consumers when assessing possible pleiotropy. Overall, sleep appears to be impacted by caffeine in a way that influences daytime alertness rather than night-time sleep characteristics. However, the presence of weak instruments for caffeine metabolism and significant heterogeneity warrants further research with larger and diverse samples to better understand the causal pathway between caffeine and sleep.

## Introduction

In the National Diet and Nutrition Survey 2008-10 (https://www.gov.uk/government/collections/national-diet-and-nutrition-survey), 95% of individuals in the UK reported caffeine consumption in the form of tea, coffee, energy drinks, chocolate, or other snacks and beverages (1), and in a 2013 study (2), 85% of US residents reported drinking at least one caffeinated beverage per day. Caffeine may increase alertness and concentration (3), although caffeine consumption has also been associated with several adverse effects, including caffeine addiction, poorer sleep quality, and reduced sleep efficiency (4,5). A recent review of 58 independent epidemiological studies and randomised controlled trials investigated the association of caffeine consumption with poor sleep including insomnia (5). The review found that adolescents and adults with high caffeine intake from tea, coffee, and soda were 1.9 times more likely to have trouble sleeping and 1.8 times more likely to feel sleepy in the morning compared to those with lower caffeine intake. Previous studies have highlighted strong associations between poor sleep quality or disturbed sleep and major health issues such as cardiovascular disease and metabolic disorders, diabetes mellitus, as well as respiratory disorders (6). Therefore, a deeper understanding of the impact of caffeine consumption on sleep behaviours could aid public health messaging and facilitate effective dietary advice on caffeine consumption.

The effects of caffeinated product consumption on sleep have previously been attributed to the presence of caffeine in the bloodstream (7). Following ingestion, caffeine is almost completely metabolised (_∼_95%). Less than 3% of caffeine remains unchanged and is excreted through urine (8). Of the 95% of caffeine (chemical name: 1,3,7-trimethylxanthine [137X]) that is metabolised, 70-80% is metabolised in the liver by the CYP1A2 enzyme to form paraxanthine (chemical name: 1,7-dimethylxanthine [17X]) (9–11). Caffeine exposure (presence of caffeine in bloodstream) for an individual depends on how quickly they metabolise caffeine, which can be quantified using the caffeine metabolite ratio (CMR) - the ratio of paraxanthine to caffeine (7) (**Supplementary Figure 1**). Those with a higher CMR are exposed to less caffeine given a fixed level of intake, therefore could be at lower risk of sleep issues due to caffeine exposure (12,13).

The extent to which caffeine affects sleep behaviours in the general population has not been fully established. Traditional epidemiological studies which have been used to explore the influence of caffeine consumption on sleep behaviour may be limited by reverse causality (i.e., where poor sleep and daytime sleepiness in turn influence caffeine intake (5)) as well as residual confounding (3) by factors including diet, physical activity, other substances like smoking or alcohol use, and socio-economic position (4). Additionally, caffeine consumption measured by self-reported cups of tea and/or coffee per day may be prone to measurement error, especially as individuals are more likely to misremember their consumption patterns (14). On the other hand, CMR is a more objective measure of caffeine exposure, calculated by profiling blood plasma levels of caffeine metabolites (7).

Mendelian Randomisation (MR) is a causal method that helps mitigate the limitations of observational study design by employing genetic variants as instruments (instrumental variables or IV) for exploring the relationship between exposure and outcome (15). However, MR results provide valid causal inference only when several MR assumptions are met (**Figure 1**). The core assumptions are (16):

1. IV1 (“relevance”): instruments must be associated with the exposure,
2. IV2 (“exchangeability” or “independence”): there are no confounders of the association between instruments and the outcome,
3. IV3 (“exclusion restriction”): instruments are not related to the outcome other than via the exposure.

**Figure 1.**
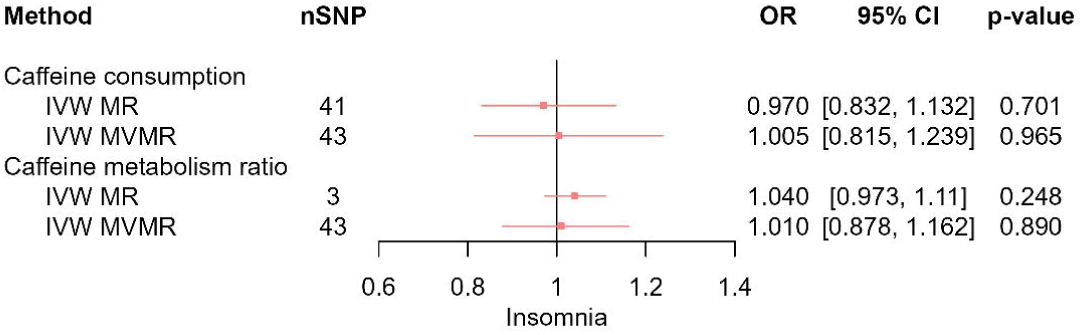
Assumption in Mendelian Randomisation (MR) framework. “G” represents the genetic variants, “X” represents the exposure, “Y” represents the outcome, and “U” represents the confounders confounding the relationship between exposure and outcome. “[1]” represents the first assumption (IV1) of the MR framework or “relevance” assumption, “[2]” represents the second assumption (IV2) or “exchangeability” assumption, and “[3]” represents the third assumption (IV3) or “exclusion restriction” assumption.

Genetic variants (single nucleotide polymorphisms or SNPs) identified from genome-wide association studies (GWASs) are used as instruments (i.e., a proxy for actual exposure) in MR. Genetic variants are randomly assigned during meiosis and fixed at conception and cannot be changed as a result of exposure to confounding variables post-conception. Hence, MR mitigates reverse causation, residual confounding, and tests for causal effects of exposure on outcome without bias given non-violation of MR assumptions.

A previous study using the MR approach found that higher plasma caffeine affected chronotype (morning/evening preference) but was not linked with other sleep traits, concluding that the associations between caffeine and poor sleep might be driven by shared environmental factors (3). However, using caffeine metabolism (substituting consumption) in an univariable MR framework to estimate the effects of caffeine on sleep behaviours, as was done in this previous study, may potentially lead to ambiguous results. Individuals with faster CMR, i.e., lower levels of caffeine in blood per beverage, consume more caffeinated products because the effects of caffeine wear off faster.

The investigation of caffeine consumption in an MR framework also remains challenging. A recent study by Woolf et. al. (17) has revealed that 16 SNPs associated with caffeine consumption act through caffeine metabolism, while the other half are linked with behavioural traits like smoking, alcohol consumption, education, and physical activity, which are likely to violate the IV2 assumption. This potentially renders caffeine consumption SNPs as invalid instruments. Moreover, evidence from a previous study suggests that individuals with higher genetically predicted caffeine metabolism rates, tend to consume higher amounts of caffeine on average to experience caffeine’s alertness-promoting effects (18). This may influence the estimated total effects of CMR because the genetic variants associated with faster metabolism may also lead to higher caffeine intake. Hence, estimated total effects of CMR may reflect the biological impact of faster caffeine metabolism combined with the behavioural impact of higher caffeine consumption.

One alternative is to employ genetic variants for caffeine consumption (cups per day, CPD) and caffeine metabolism (CMR) as instruments in a multivariable-MR (MVMR) framework. MVMR is an extension of MR that can be used to establish causal effects of two or more related exposures, estimating the direct causal effect of each exposure on an outcome, conditional on the other exposures included in the model (19). Using this approach, we aimed to explore the direct effects of caffeine consumption on sleep behaviours accounting for caffeine metabolism, and direct effects of caffeine metabolism on sleep behaviours accounting for consumption (**Figure 2**).

**Figure 2.**
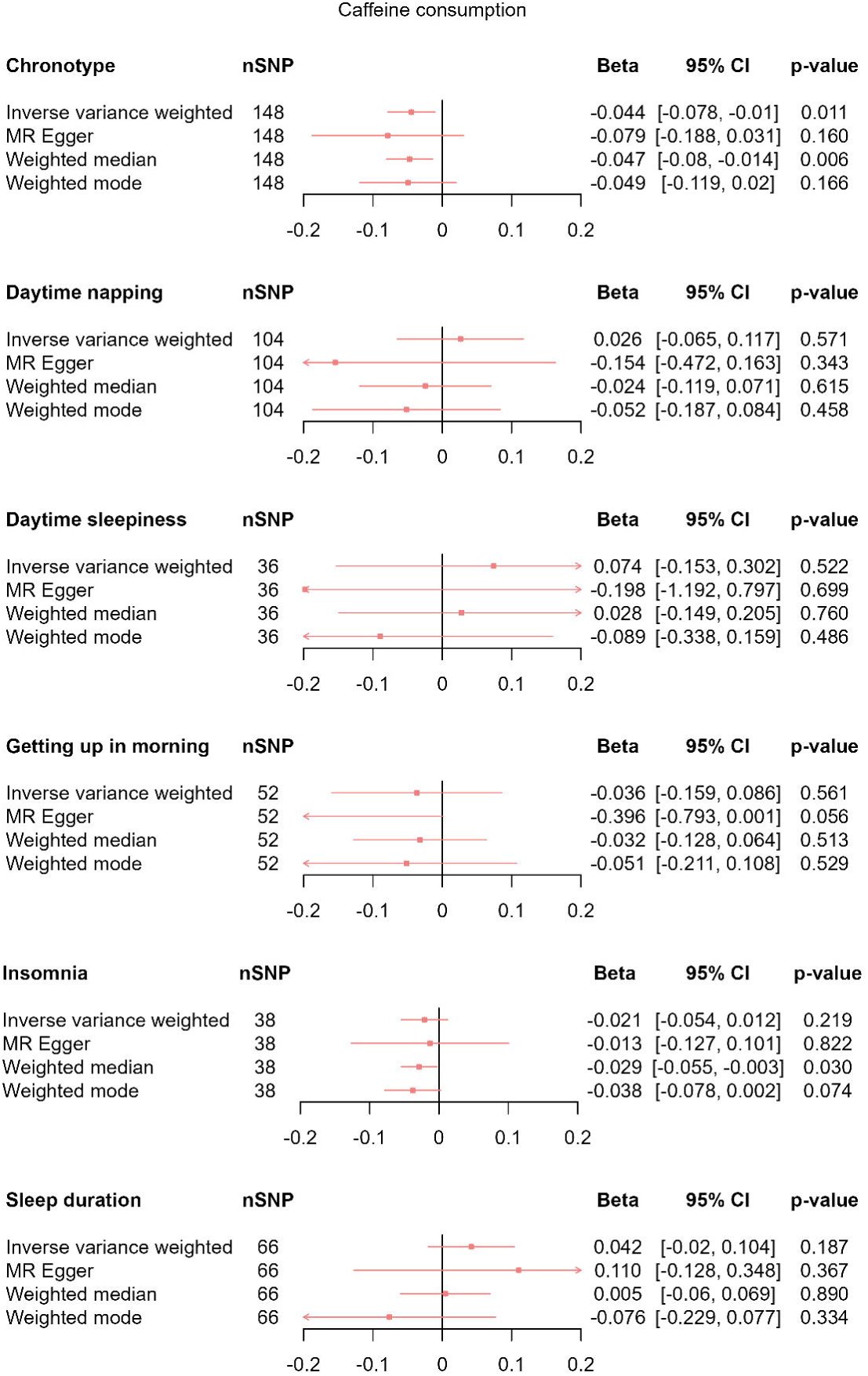
Multivariable Mendelian Randomisation (MVMR) framework. “G1”, “G2”, and “G3” represent the genetic variants. “U” represents the confounders confounding the relationship between exposures (caffeine consumption and caffeine metabolism ratio) and outcomes (sleep behaviours). “[1]” represents the first assumption (IV1) of the MVMR framework or “relevance” assumption, “[2]” represents the second assumption (IV2) or “exchangeability” assumption, and “[3]” represents the third assumption (IV3) or “exclusion restriction” assumption.

## Methods

### Data Sources

#### Caffeine consumption GWASs

Summary statistics were obtained from separate GWASs performed on estimated caffeine consumed from tea, coffee, and combined tea and coffee in a day based on 407,072 individuals of European ancestry in UK Biobank (UKB) (14). Each GWAS was adjusted for age, sex, genotyping array, and the first 30 genetic principal components (PCs) to adjust for population stratification. The UKB and the methods used for each GWAS are detailed in the **Supplementary Methods**.

### Plasma caffeine and caffeine metabolites GWAS

Summary statistics were obtained from a meta-analysis of six GWASs performed on caffeine metabolism (CMR) and blood plasma caffeine (BPC) based on 9,876 individuals of European ancestry (7). The GWASs were adjusted for smoking status, age, sex, site of the study, fasting status, and family structure and/or study specific PCs of ancestry. The meta-GWAS is discussed in the **Supplementary Methods**.

### Sleep behaviour GWASs

Summary statistics on sleep behaviours (chronotype (20), daytime napping (21), daytime sleepiness (22), getting up in the morning (20), sleeplessness/insomnia (23), and sleep duration (24)) were obtained from GWASs performed on self-reported traits from approximately 500,000 individuals of European ancestry in UKB available in the Sleep Disorders Knowledge Portal (SDKP) (sleepdisordergenetics.org). Details on the UKB data fields, sample sizes, questions and response choices for sleep behaviours are available in the **Supplementary Methods**. We used summary statistics from two separate insomnia GWASs (25,26) which meta-analysed sleep data from UKB and 23andMe data to cross-check primary results.

Further, summary statistics from GWAS performed using accelerometer-derived sleep traits from UKB (27) were obtained for “sleep duration”, “least active 5 hours” (L5) and “most active 10 hours” (M10) (chronotype equivalent), “number of nocturnal sleep episodes” and “sleep efficiency” (aspects of insomnia), and “diurnal inactivity” (consistent with daytime napping) available in the SDKP.

Additionally, we conducted our own GWAS for sleep behaviours stratified by caffeine consumption status (current/non-current) using UK Biobank Resource under application numbers 16391 and 81499. Stratified-GWASs were performed using the MRC IEU UKB GWAS pipeline (28) adjusting for sex, genotyping chip, relatedness, and population stratification. Details of the GWAS pipeline are provided in the **Supplementary Methods**.

### Statistical analysis

#### Univariable MR

First, we used a two-sample MR (2SMR) framework with inverse variance weighted (IVW), MR-Egger, weighted mode, and weighted median models to assess the total causal effects of caffeine consumption (CPD) and caffeine metabolism (CMR) on sleep behaviours. These methods are detailed in **Supplementary Methods**. Exposures were clumped at a linkage disequilibrium (LD) threshold of r^2^<0.001, with a clumping window of >10,000kb. Genome-wide (GW) significant (p<5×10^-8^) SNPs were selected and data were harmonised prior to performing MR to ensure SNP-exposure and SNP-outcome effects corresponded to the same effect allele (19,29,30). We performed 2SMR using Steiger-filtering to ensure the correct direction of causality i.e. to confirm that the SNPs used to instrument CMR/CPD are more strongly associated with caffeine metabolism/intake than sleep behaviour (15). We removed invalid CPD SNPs that were more strongly associated with sleep behaviours than caffeine intake (chronotype: 4, daytime napping: 2, getting up in the morning: 4, insomnia: 3, sleep duration: 2) prior to performing MR analyses. No CMR SNPs were removed from the analyses. Further, we performed the reciprocal MR, using genetic instruments associated with sleep behaviours to evaluate their effects on caffeine consumption and metabolism.

We used an F statistic exceeding the conventional threshold of 10 to indicate good instrument strength (IV1) (31). A non-zero intercept in MR-Egger regression was used to indicate directional pleiotropic effect (violation of IV2) (32). We used funnel plots to assess the presence of horizontal pleiotropy by visually inspecting asymmetry. Cochran’s Q statistic (33) exceeding the number of SNPs included in each model and the I^2^ statistic (34) was used to indicate heterogeneity. We performed leave-one-out analyses to assess where the causal effect was consistent with removal of individual SNPs. Since GWASs were restricted to individuals of European ancestry while including genetic principal components as covariates, potential confounding due to population stratification (IV2) was minimised.

#### Multivariable MR

We assessed the role of CMR in the relationship between tea/coffee consumption and sleep behaviours using a two-sample MVMR (2S-MVMR) framework. We selected exposure SNPs using a GW significance threshold of p<5×10^-8^, then clumped (LD r^2^<0.001, clumping window

>10,000kb) (19) and harmonised the data (19,29) prior to performing analyses using IVW, and MVMR-Egger models. These methods are detailed in the **Supplementary Methods**.

We used a conditional F statistic (cF) over 10 to indicate good instrument strength (29,31). We used weak instrument robust methods – Qhet (30), debias IVW (35), Grapple (36), GMM, and IVW ME (37) – to check consistency and robustness in cases of weak instrument strength. Weak instrument robust MR methods are detailed in **Supplementary Methods**. Cochran’s Q statistics larger than number of SNPs included in the model indicate horizontal pleiotropy. We used pleiotropy robust MVMR-Egger to check consistency of results (33).

### Sensitivity analyses

Additionally, we performed 2SMR and 2S-MVMR to assess total effects of CPD and CMR, as well as direct effects of CPD and CMR, on sleep behaviours in non-current drinkers of tea and coffee. This serves as a negative control to explore violations of IV2 and IV3 assumptions, since we would not expect to observe effects of caffeine consumption and metabolism among individuals who do not drink tea or coffee. We assumed any evidence of causal effects of genetically-predicted increased caffeine intake via tea/coffee consumption among non-current drinkers of tea and coffee would be due to pleiotropy or population stratification, which may bias our primary results.

Furthermore, we performed 2SMR and 2S-MVMR using accelerometer-derived sleep traits, multiple GWASs for insomnia, assessing the effects of blood plasma caffeine levels (as well as CMR), and substituting total caffeine consumption with only tea consumption and only coffee consumption to check for consistency in our primary results.

The meta-analysis GWAS for plasma caffeine and caffeine metabolism did not include UKB data. Hence, there was no overlap in participants between the plasma caffeine and caffeine metabolism GWAS with sleep traits GWASs. However, since the caffeine consumption GWASs were conducted on UKB data, there was overlap in participants with the sleep GWASs. To assess the impact of this, we used MRLap to check for consistency when accounting for sample overlap between exposures and outcomes in two-sample MR (38).

Analyses were performed in R (version 4.3.3) utilising MendelianRandomization (version 0.10.0), mr.divw (version 0.1.0), MRlap (version 0.0.3.3), MVMR (version 0.4), and TwoSampleMR (version 0.6.4) R packages. Study findings have been reported according to the Strengthening the Reporting of Observational Studies in Epidemiology using Mendelian Randomisation (STROBE-MR) guidelines (https://www.strobe-mr.org/) (**Supplementary Table 1**) (16,39). Lastly, the study protocol was not pre-registered on any centralised scientific research databases.

## Results

Estimated odds ratios (OR_IVW-MR_ and OR_IVW-MVMR_ from IVW MR and IVW MVMR, respectively) for binary traits and estimated betas (β_IVW-MR_ and β_IVW-MVMR_ from IVW MR and IVW MVMR, respectively) for continuous traits are reported in the main text. Presented βs correspond to a change of 1 standard deviation (SD) in outcome for 1 SD increase in exposure; ORs also represent the odds per SD increase in the exposure.

### Effect of caffeine on sleep behaviours

#### Instrument strength

F statistics for CPD (82.04-106.09) and CMR (84.97) and cFs for CPD (10.89-16.58) passed the conventional threshold of 10, suggesting good instrument strength (**Supplementary Tables 2 & 3**). However, cFs for CMR (4.71-5.01) indicated weak conditional instrument strength (**Supplementary Table 3**). Hence, results from weak instrument robust methods are also presented in the **Supplementary Material (Supplementary Figure 8)**. Cochran’s Q was greater than the number of SNPs across all sleep traits (**Supplementary Table 4**), suggesting the presence of SNP heterogeneity. Similarly, I^2^ indicated that 51-70% of the variability of MR effect estimates was due to heterogeneity (**Supplementary Table 4**). Results for pleiotropy-robust methods are also presented in the **Supplementary Material (Supplementary Figures 2, 4, & 8)**.

### Effects of CPD on sleep behaviours

In IVW analyses, we found evidence that higher CPD decreased the likelihood of daytime napping (β_IVW-MR_=-0.035, 95%CI [-0.059,-0.012], p=0.004), which attenuated upon accounting for CMR (β_IVW-MVMR_=-0.015, 95%CI [-0.046,0.015], p=0.332) (**Figure 3a**). We found evidence that higher CPD decreased the likelihood of daytime sleepiness (β_IVW-MR_=-0.044, 95%CI [-0.065,-0.023], p<0.001) which was slightly attenuated upon accounting for CMR (β_IVW-MVMR_=-0.034, 95%CI [-0.058,-0.009], p=0.010), however the confidence intervals still excluded the null (**Figure 3b**). We found evidence that higher CPD decreased ease of getting up in the morning (β_IVW-MR_=-0.045, 95%CI [-0.083,-0.007], p=0.021; β_IVW-MVMR_=-0.034, 95%CI [-0.087,0.018], p=0.209) (**Figure 3c**). We found no clear evidence of any total effects of CPD or any direct effects of CPD on sleep duration (β_IVW-MR_=0.019, 95%CI [-0.021,0.059], p=0.360; β_IVW-MVMR_=0.035, 95%CI [-0.019,0.089], p=0.208) (**Figures 3d**), chronotype (β_IVW-MR_=-0.024, 95%CI [-0.086,0.037], p=0.437; β_IVW-MVMR_=-0.012, 95%CI [-0.106,0.082], p=0.807) (**Figure 3e**), or insomnia (OR_IVW-MR_=0.970, 95%CI [0.832,1.132], p=0.701; OR_IVW-MVMR_=1.005, 95%CI [0.815,1.239], p=0.965) (**Figure 3f**). We observed a pattern of attenuated effect estimates across sleep behaviours upon adjusting for CMR, with the exception of the effect of CPD on sleep duration which strengthened when accounting for CMR (β_IVW-MR_=0.028, β_IVW-MVMR_=0.040), although the confidence intervals included the null (**Figure 3d**). Effect estimates were similar using pleiotropy robust and weak instrument robust MVMR methods across all sleep behaviours presented in the **Supplementary Results (Supplementary Figure 8).** Additionally, we found weak evidence that higher CPD decreased ease of getting up in the morning in pleiotropy-robust univariable MR methods, contrary to our primary results (**Supplementary Figure 2**).

**Figure 3a.**
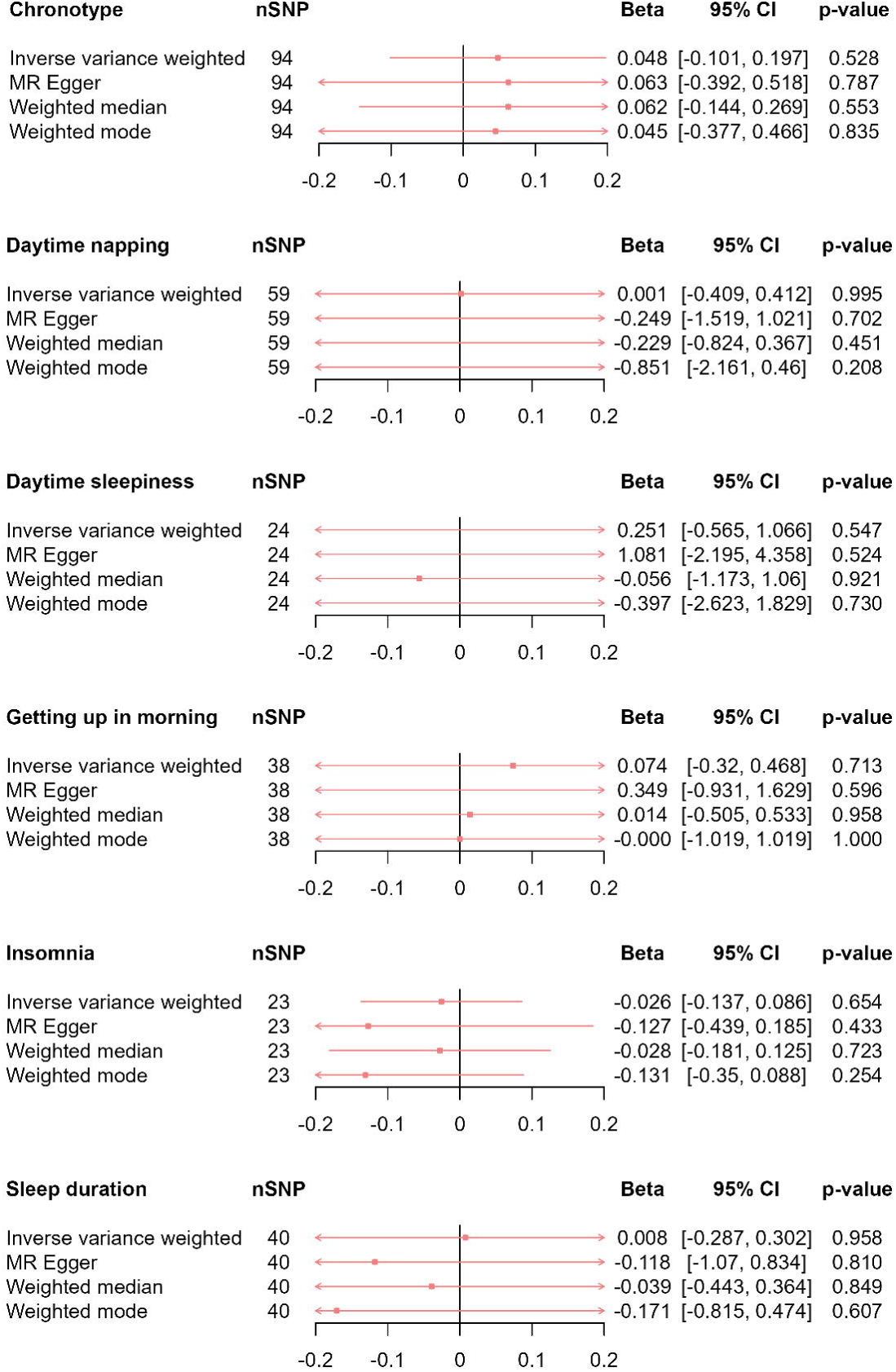
MR results of effects of caffeine consumption and caffeine metabolism ratio on daytime napping. Red squares represent the estimated effect sizes (betas or odds ratios (ORs)) for each individual estimation method. The red horizontal lines represent the 95% confidence intervals (95% CI) for the estimated effects. The black vertical line represents the point of no effect. “nSNP” gives the number of single nucleotide polymorphisms (SNPs) used in each estimation method. “IVW MR” gives the total effects estimated using inverse variance weighted (IVW) Mendelian Randomisation (MR). “IVW MVMR” gives the direct effects estimated using Multivariable Mendelian Randomisation (MVMR).

**Figure 3b.**
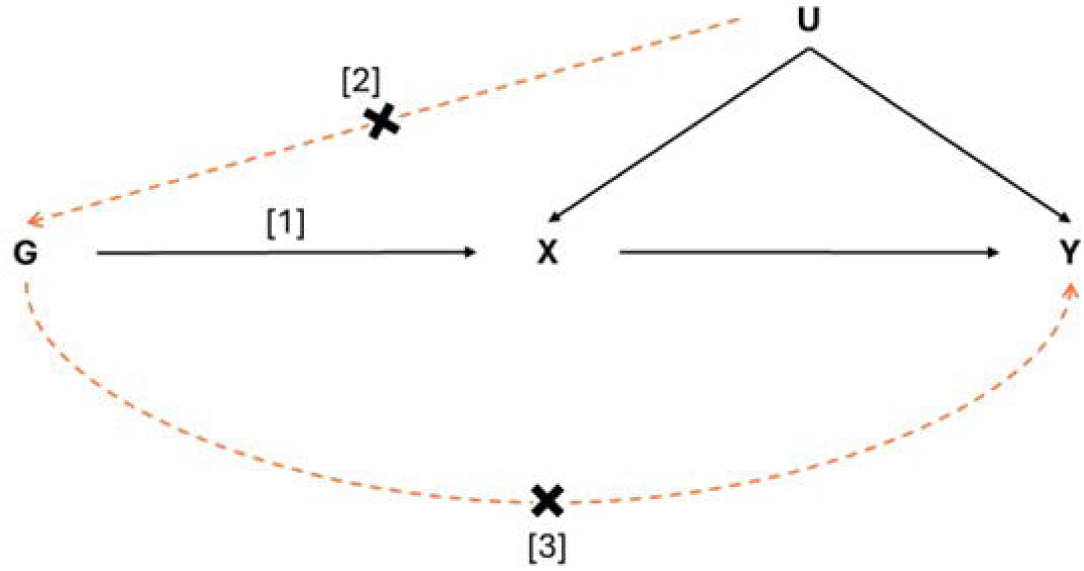
MR results of effects of caffeine consumption and caffeine metabolism ratio on daytime sleepiness. Red squares represent the estimated effect sizes (betas or odds ratios (ORs)) for each individual estimation method. The red horizontal lines represent the 95% confidence intervals (95% CI) for the estimated effects. The black vertical line represents the point of no effect. “nSNP” gives the number of single nucleotide polymorphisms (SNPs) used in each estimation method. “IVW MR” gives the total effects estimated using inverse variance weighted (IVW) Mendelian Randomisation (MR). “IVW MVMR” gives the direct effects estimated using Multivariable Mendelian Randomisation (MVMR).

**Figure 3c.**
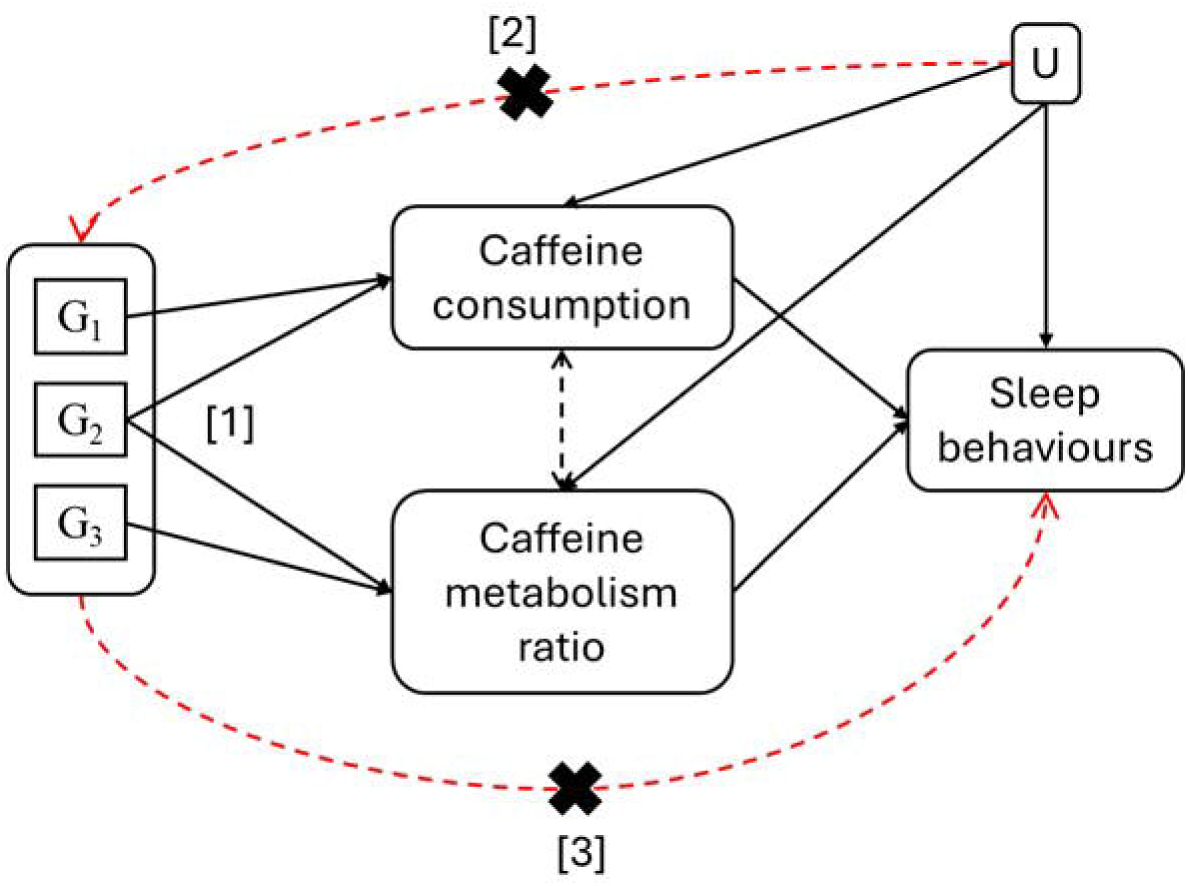
MR results of effects of caffeine consumption and caffeine metabolism ratio on sleep duration. Red squares represent the estimated effect sizes (betas or odds ratios (ORs)) for each individual estimation method. The red horizontal lines represent the 95% confidence intervals (95% CI) for the estimated effects. The black vertical line represents the point of no effect. “nSNP” gives the number of single nucleotide polymorphisms (SNPs) used in each estimation method. “IVW MR” gives the total effects estimated using inverse variance weighted (IVW) Mendelian Randomisation (MR). “IVW MVMR” gives the direct effects estimated using Multivariable Mendelian Randomisation (MVMR).

**Figure 3d.**
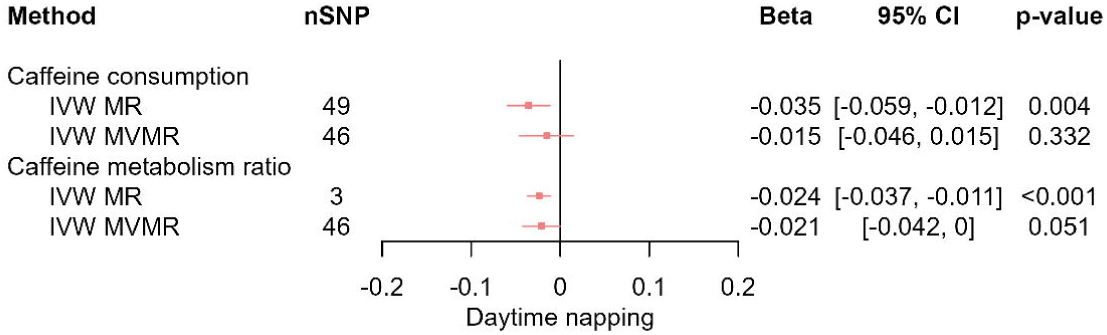
MR results of effects of caffeine consumption and caffeine metabolism ratio on getting up in morning. Red squares represent the estimated effect sizes (betas or odds ratios (ORs)) for each individual estimation method. The red horizontal lines represent the 95% confidence intervals (95% CI) for the estimated effects. The black vertical line represents the point of no effect. “nSNP” gives the number of single nucleotide polymorphisms (SNPs) used in each estimation method. “IVW MR” gives the total effects estimated using inverse variance weighted (IVW) Mendelian Randomisation (MR). “IVW MVMR” gives the direct effects estimated using Multivariable Mendelian Randomisation (MVMR).

**Figure 3e.**
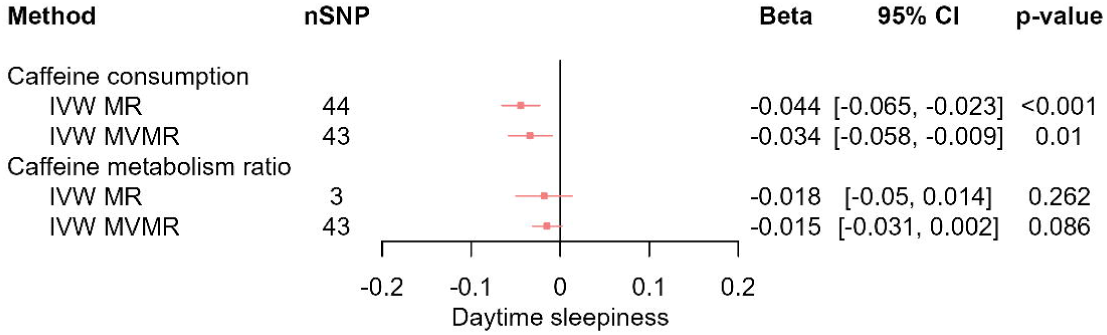
MR results of effects of caffeine consumption and caffeine metabolism ratio on chronotype. Red squares represent the estimated effect sizes (betas or odds ratios (ORs)) for each individual estimation method. The red horizontal lines represent the 95% confidence intervals (95% CI) for the estimated effects. The black vertical line represents the point of no effect. “nSNP” gives the number of single nucleotide polymorphisms (SNPs) used in each estimation method. “IVW MR” gives the total effects estimated using inverse variance weighted (IVW) Mendelian Randomisation (MR). “IVW MVMR” gives the direct effects estimated using Multivariable Mendelian Randomisation (MVMR).

**Figure 3f.**
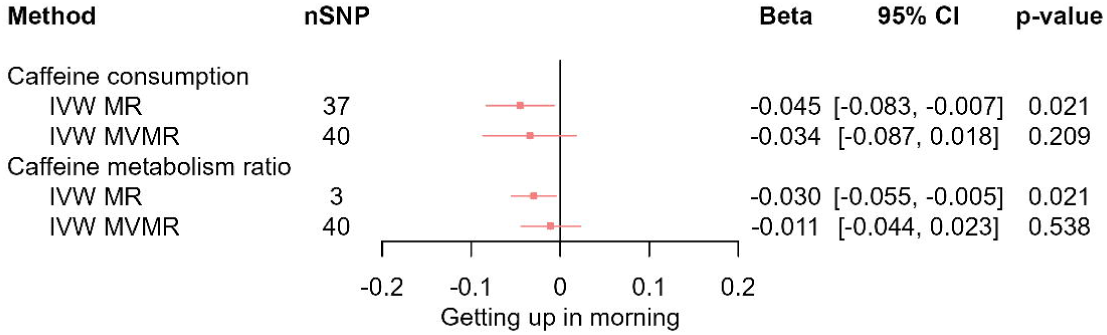
MR results of effects of caffeine consumption and caffeine metabolism ratio on insomnia. Red squares represent the estimated effect sizes (betas or odds ratios (ORs)) for each individual estimation method. The red horizontal lines represent the 95% confidence intervals (95% CI) for the estimated effects. The black vertical line represents the point of no effect. “nSNP” gives the number of single nucleotide polymorphisms (SNPs) used in each estimation method. “IVW MR” gives the total effects estimated using inverse variance weighted (IVW) Mendelian Randomisation (MR). “IVW MVMR” gives the direct effects estimated using Multivariable Mendelian Randomisation (MVMR).

### Effects of CMR on sleep behaviours

In IVW analyses, we found evidence that faster caffeine metabolism (CMR) decreased the likelihood of daytime napping (β_IVW-MR_=-0.024, 95%CI [-0.037,-0.011], p<0.001; β_IVW-MVMR_=-0.021, 95%CI [-0.042, 0.000], p=0.051) (**Figure 3a**). We found evidence that faster CMR decreased the ease of getting up (β_IVW-MR_=-0.030, 95%CI [-0.055,-0.005], p=0.021) which attenuated when accounting for CPD (β_IVW-MVMR_=-0.011, 95%CI [-0.044,0.023], p=0.538) (**Figure 3c**). We found weak evidence that faster CMR decreased the likelihood of being an evening person, but again this attenuated when accounting for CPD (β_IVW-MR_=-0.044, 95%CI [-0.09,0.002], p=0.058; β_IVW-MVMR_=-0.023, 95%CI [-0.086,0.04], p=0.481) (**Figure 3e**). Additionally, we found no clear evidence of any total effects of CMR or any direct effects of CMR on daytime sleepiness (β_IVW-MR_=-0.018, 95%CI [-0.05,0.014], p=0.262; β_IVW-MVMR_=-0.015, 95%CI [-0.031,0.002], p=0.086) (**Figure 3b**), sleep duration (β_IVW-MR_=0.005, 95%CI [-0.014,0.025], p=0.603; β_IVW-MVMR_=-0.010, 95%CI [-0.046,0.026], p=0.578) (**Figures 3d**), or insomnia (OR_IVW-MR_=1.040, 95%CI [0.973,1.11], p=0.248; OR_IVW-MVMR_=1.010, 95%CI [0.878,1.162], p=0.890) (**Figure 3f**). We observed patterns of attenuation for the effect of CMR on sleep behaviours after adjusting for CPD.

Total effect estimates as well as direct effect estimates of CMR on other sleep traits were consistent across pleiotropy robust and weak instrument robust methods detailed in the **Supplementary Results (Supplementary Figure 8)**. However, in our pleiotropy robust univariable MR models, we found evidence that a faster CMR (total effect) increased the likelihood of daytime sleepiness (**Supplementary Figure 4**), with some supporting evidence from the MVMR models (**Supplementary Figure 8c**).

### Effects of sleep behaviours on caffeine consumption and metabolism

We next evaluated the effects of sleep behaviours (exposure) on CPD and CMR (outcome). F statistics for sleep behaviours were between 40.76 and 52.79 (**Supplementary Table 5**). Q statistics were greater than number of SNPs across all sleep traits, indicating the presence of SNP heterogeneity which was further supported by high I^2^ statistics (61-80%) when considering CPD as the outcome (**Supplementary Table 6**). However, I^2^ statistics suggested that no amount of the variation (0%) was due to heterogeneity when considering CMR as the outcome (**Supplementary Table 6**). Results for pleiotropy-robust methods are also presented (**Figure 4**).

Using IVW-MR, we found evidence that being an evening person decreased the amount of CPD (β_IVW-MR_=-0.044, 95%CI [-0.078,-0.010], p=0.011). We observed consistent direction of effect estimates with wider confidence intervals in pleiotropy robust methods (**Figure 4a**). We found no clear evidence for effects of daytime napping (β_IVW-MR_=0.026, 95%CI [- 0.065,0.117], p=0.571), daytime sleepiness (β_IVW-MR_=-0.074, 95%CI [-0.153,0.302], p=0.699), ease of getting up in morning (β_IVW-MR_=-0.036, 95%CI [-0.159,0.086], p=0.561), insomnia (β_IVW-MR_=-0.021, 95%CI [-0.054, 0.012], p=0.219), or sleep duration (β_IVW-MR_=0.042, 95%CI [- 0.02,0.104], p=0.187) on CPD (**Figure 4a**), or any effects of sleep behaviours on CMR (chronotype: β_IVW-MR_=0.048, 95%CI [-0.101,0.197], p=0.528; daytime napping: β_IVW-MR_=0.001, 95%CI [-0.409,0.412], p=0.995; daytime sleepiness: β_IVW-MR_=-0.251, 95%CI [-0.565,1.066], p=0.547; getting up in morning: β_IVW-MR_=0.074, 95%CI [-0.320,0.468], p=0.713; insomnia: β_IVW-MR_=-0.026, 95%CI [-0.137,0.086], p=0.654; sleep duration: β_IVW-MR_=0.008, 95%CI [- 0.287,0.302], p=0.958) (**Figure 4b**). Effect estimates were consistent across pleiotropy robust methods (**Figure 4**).

**Figure 4a.**
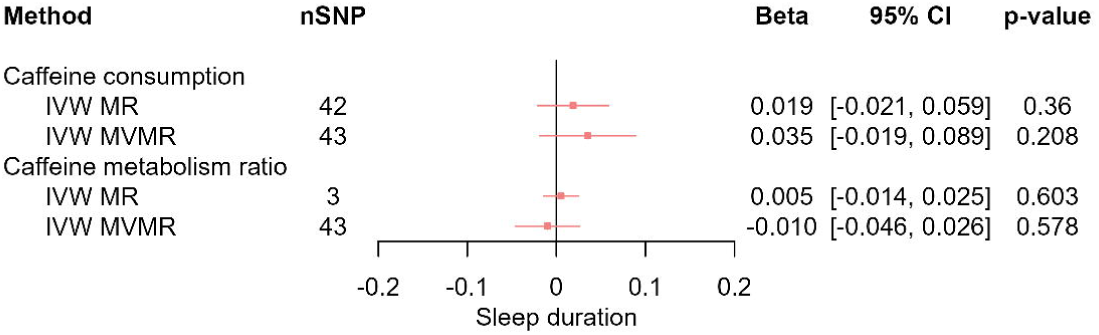
MR results of total effect of sleep behaviours on caffeine consumption. Red squares represent the estimated effect sizes (betas) for each individual estimation method. The red horizontal lines represent the 95% confidence intervals (95% CI) for the estimated effects. The black vertical line represents the point of no effect. “nSNP” gives the number of single nucleotide polymorphisms (SNP) used in each estimation method.

**Figure 4b.**
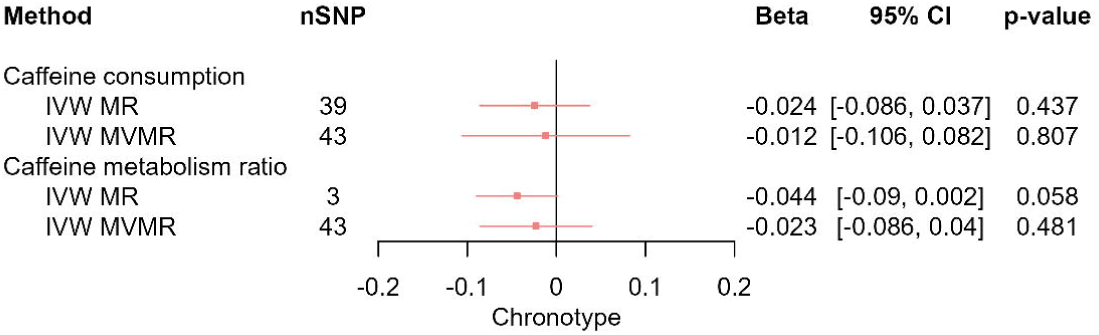
MR results of total effect of sleep behaviours on caffeine metabolism ratio. Red squares represent the estimated effect sizes (betas) for each individual estimation method. The red horizontal lines represent the 95% confidence intervals (95% CI) for the estimated effects. The black vertical line represents the point of no effect. “nSNP” gives the number of single nucleotide polymorphisms (SNP) used in each estimation method.

Lastly, we did not observe any violations of assumptions in any of our bi-directional MR frameworks as detailed **Supplementary Results**. Sensitivity analyses results and bi-directional MR assumptions are further detailed in **Supplementary Results (Supplementary Figures 3, 5, 6, & 7)**.

### Sensitivity analyses

For the most part, primary MR and MVMR results were replicated among tea consumers (**Supplementary Figure 15a**), coffee consumers (**Supplementary Figure 15b**), and current caffeine consumers (**Supplementary Figures 13 & 14**), using different insomnia GWASs (**Supplementary Figures 9g & 9h**), and when CMR was substituted for BPC (**Supplementary Figure 10**). However, the total effects of CMR (including substitution with BPC) on chronotype were not consistent across our sensitivity analyses, where we found evidence for a positive effect (evening preference) in current caffeine consumers (β_IVW-MR_=0.034, 95%CI [0.010,0.057], p=0.005), and no evidence of any effects when substituted with BPC (β_IVW-MR_=0.020, 95%CI [-0.011,0.051], p=0.209) compared to evidence for a weak inverse effect in our primary analysis (β_IVW-MR_=-0.044, 95%CI [-0.09,0.002], p=0.058) (**Supplementary Figure 14a**).

For accelerometer-derived sleep traits, we found evidence of an inverse effect of CPD on diurnal inactivity, which remained following adjustment for CMR (β_IVW-MR_=-0.051, 95%CI [- 0.095,-0.007], p=0.023; β_IVW-MVMR_=-0.060, 95%CI [-0.111,-0.008], p=0.028) (**Supplementary Figure 9a**). Further, we found evidence that higher CPD (unadjusted for CMR) increased sleep efficiency, which slightly attenuated on adjustment for CMR (OR_IVW-MR_=1.007, 95%CI [1.001, 1.013], p=0.019, OR_IVW-MVMR_=1.005, 95%CI [0.999, 1.012], p=0.116) (**Supplementary Figure 9f**). However, we did not find any total or direct effects of CPD on L5 time (**Supplementary Figure 9b**), M10 time (**Supplementary Figure 9c)**, number of nocturnal sleep episodes (**Supplementary Figure 9d)**, and accelerometer-derived sleep duration (**Supplementary Figure 9e)**, or any effect of CMR on the accelerometer-derived sleep traits (**Supplementary Figures 9a-f**).

As part of the negative control analysis, we found no clear evidence of any total effects of CPD or CMR, or direct effects of CPD or CMR on the sleep traits among non-current caffeine consumers (**Supplementary Figures 11 & 12**).

Additionally, our primary results were replicated after adjusting for sample overlap between CPD and the sleep traits of interest (**Supplementary Figure 16**).

Sensitivity analyses results are further detailed in **Supplementary Results (Supplementary Figures 9, 10, & 13-16)**.

## Discussion

In this study, we explored total and direct effects of caffeine consumption measured as cups per day (CPD) on sleep behaviours, adjusting for caffeine metabolism (CMR). Furthermore, we explored the total and direct effects of caffeine measured by CMR on sleep behaviours, adjusting for CPD. In our univariable and multivariable MR analyses, we observed inverse effects of increased caffeine consumption from tea/coffee on daytime napping, daytime sleepiness, and getting up in the morning. We also observed inverse effects of increased caffeine metabolism on chronotype, daytime napping, daytime sleepiness, and getting up in the morning. Additionally, we observed an inverse effect of evening chronotype on caffeine consumption. Consistent with previous MR study findings, we found no clear evidence for effects of caffeine consumption or metabolism on sleep duration or insomnia (3,40). Our findings suggest that the effects of caffeine might be more pronounced in daytime alertness phenotypes (chronotype, daytime napping, and daytime sleepiness) over nocturnal sleep phenotypes (ease of getting up in the morning, insomnia, and sleep duration).

Findings that higher caffeine consumption decreased the likelihood of daytime napping, daytime sleepiness, and the ease of getting up are consistent with previous observational studies (41,42). We observed a pattern of attenuation of the effects of caffeine consumption when accounting for caffeine metabolism, suggesting that observed effects of consumed caffeine on sleep behaviours are primarily due to increased circulating caffeine. However, the direct effects of caffeine consumption were often non-zero, indicating presence of residual effects from additional constituents of caffeinated drinks such as milk, antioxidants, and sugar, or residual (genetic) confounding by lifestyle/environmental factors as previously suggested (3,17). Caffeine tolerance may also interact with the effect of caffeine consumption independently of metabolism (4), contributing to observed residual effects of CPD. Further, potential horizontal pleiotropy may stem from genetic variations in adenosine (a sleep-promoting neurotransmitter blocked by caffeine) receptor genes. However, we did not observe any evidence of horizontal pleiotropy in our analyses. We found evidence that a faster caffeine metabolism (CMR) decreased the likelihood of daytime napping, daytime sleepiness, and the ease of getting up (3). Individuals with genetic variants associated with faster CMR are more efficient at breaking down and eliminating caffeine (lesser caffeine exposure for a given consumed amount). Faster CMR may lead to more rapid onset and offset of caffeine’s stimulant effects, resulting in acute alertness benefits without prolonged exposure to caffeine (less interference with adenosine) that could lead to sleep disturbances – allowing for a smoother transition between periods of wakefulness and sleep, thus reducing the physiological need for daytime napping (43).

Similar to Treur and colleagues, we found weak evidence that faster CMR (unadjusted for CPD) decreased the likelihood of being an evening person (3). Contrarily, we also found evidence that faster CMR (unadjusted for CPD) increased the likelihood of being an evening person among current consumers. However, these were attenuated to the null in the CPD-adjusted MVMR models. Moreover, observed effects of CMR in our analyses were in the same (inverse) direction as the effects of CPD.

Contrary to a previous study that employed a similar framework to explore the stimulating effects of nicotine on sleep while adjusting for nicotine metabolism rate (NMR) (44), our findings indicate contrasting effects of CMR on certain sleep traits (chronotype, daytime napping, daytime sleepiness, ease of getting up in the morning), where the observed CMR effects appear to be in the same direction (negative) as that of CPD implying that both a faster CMR and a higher CPD increase alertness. Differences in direction of effects of stimulant metabolism rates could stem from different half-lives of the stimulants. The stimulating effects of caffeine (half-life: 5 hours) are likely to last longer and potentially into the night compared to nicotine which has a shorter half-life (2 hours). Moreover, while NMR captures the rate of inactivation of nicotine, CMR is measured as the ratio of paraxanthine to caffeine, where paraxanthine is also an active stimulant. Evidence from functional studies suggests that paraxanthine (the primary metabolite of caffeine) imparts a similar and potentially more efficacious stimulating effect to caffeine (45). The effects of paraxanthine likely contribute to the inverse direction of observed direct effects of CMR in our analyses. A faster CMR leads to a more rapid metabolism of caffeine into paraxanthine, resulting in sustained alertness benefits of paraxanthine without exposure to caffeine for an extended period. Nonetheless, potential physiological effects of paraxanthine are likely to be relatively brief, as its half-life is about 25% shorter than that of caffeine (46).

Additionally, Treur and colleagues did not observe any effects of sleep behaviours on caffeine consumption (3). However, we found evidence that identifying as an evening person decreased CPD.

### Strengths and limitations

This study has several notable strengths. We isolated the effects of caffeine consumption and metabolism on sleep behaviours by using a novel design which includes caffeine metabolism ratio (CMR) as a proxy measure for caffeine in MVMR analyses, while accounting for caffeine consumption (CPD). Additionally, the large sample sizes of GWASs for caffeine consumption and sleep behaviours increase statistical power to detect effects in our analyses, compared with previous MR studies (3,40).

However, the study also has some limitations. The small sample size of the caffeine metabolism GWAS relative to caffeine consumption GWAS reduced power to detect direct effects of CMR on sleep behaviours. Additionally, we observed presence of heterogeneity, and pleiotropy in some of our analyses, and weak instrument strength for CPD and CMR in our MVMR analyses. However, this was largely overcome with the use of pleiotropy-, weak instrument-, and sample overlap-robust methods (31–33,38), where we observed largely consistent results. Our primary results were also supported by similar effect estimates observed among tea consumers, coffee consumers, and current caffeine consumers. Further, we found no evidence of any effects of CPD or CMR on sleep traits among non-current caffeine consumers. This analysis serves as a negative control, suggesting evidence against pleiotropy (19,29).

Participants misremembering or incorrectly reporting caffeine consumption and sleep behavioural patterns may have introduced measurement error in the self-reported caffeine consumption and sleep traits (47). While we checked consistency of findings using accelerometer-derived sleep behaviour data, small sample sizes for accelerometer-derived sleep trait GWASs resulted in low power to detect effects. However, while MR estimates can be biased by differential measurement error, they are less affected than observational estimates from standard regression (37).

MR and MVMR analyses were performed using GWAS summary statistics based on cohorts primarily of European ancestry. Hence, study findings suffer from a lack of generalisability to populations with non-European ancestry (19). Moreover, CPD GWAS included exposure to both caffeinated and de-caffeinated tea/coffee, which may have diluted GWAS signals. Additionally, there was overlap in participants since GWASs for caffeine consumption and sleep behaviours were conducted using UKB data which may have led to some biased estimates (48). However, a simulation study has shown that two-sample MR methods used in a one-sample setting perform similarly to when used in a two-sample setting (49). The study further concluded that two-sample MR methods, except for MR-Egger, can be safely used for one-sample MR performed on large biobanks. Further, we used a sample overlap-robust method (38) to account for the overlap and observed consistent results.

Future studies should utilise larger GWAS on CMR to cross-check our study findings. UKB is a non-representative study with about 5.5% participant response rate (50). Future assessment of the effects of caffeine on sleep traits should be conducted on studies other than UKB to validate our findings. GWASs based on individuals of non-European ancestry should be used in future studies to improve generalisability of our study findings or potentially reveal evidence for differences in the effect of caffeine on sleep behaviours between ancestries. Finally, using GWASs performed on caffeinated coffee consumption might further strengthen evidence for effects. Although we observed consistent results in our sensitivity analyses separating out tea and coffee consumption (contrary to our primary analyses using combined tea and coffee consumption), the inclusion of decaffeinated coffee in the coffee consumption measurement may have resulted in a slight underestimation of the observed effects of caffeine.

### Implications

Exploring the relationship between sleep and caffeine consumption is important for informing public health guidelines for caffeine consumption and sleep hygiene, while knowledge of genetic predisposition to caffeine metabolism could improve personalised advice regarding how caffeine consumption might affect sleep. Our findings support the role of caffeine on certain sleep behaviours (specifically daytime napping and daytime sleepiness), which could have important implications for public health initiatives aimed at better sleep hygiene, diet, and overall quality of life. However, the limited evidence for a (long-term) effect of caffeine consumption and metabolism on sleep duration and insomnia, indicates that the stimulating effects of caffeine are typically acute, and suggests that the association between habitual caffeine consumption and poor sleep seen observationally may be attributed to residual confounding by shared environmental factors.

### Conclusion

In conclusion, we found evidence for effects of higher caffeine consumption levels on certain sleep behaviours (decreased daytime napping and daytime sleepiness, and to a lesser extent, decreased ease of getting up), and evidence of faster caffeine metabolism on chronotype (morning preference), and decreased daytime napping and ease getting up. In our MVMR analyses, we found evidence for effects of higher consumption levels on reduced daytime sleepiness and evidence for effects of faster metabolism on reduced daytime napping and daytime sleepiness, suggesting that caffeine plays an important role in determining daytime alertness. A higher caffeine metabolism rate may allow an individual to experience caffeine’s alertness-promoting effects within a shorter period of time. Thus, the sleep homeostasis is not disrupted for an extended period of time. This, in turn, could reduce the need for compensatory sleep which is experienced by individuals with slower caffeine metabolism. Our findings suggest that faster metabolism of caffeine may not result in reduced stimulating effects when accounting for the amount of caffeine consumed per day. Since the stimulating effects of caffeine may be sustained by paraxanthine, which is the primary metabolite of caffeine, further exploration of the impact of the metabolites of caffeine on sleep traits is warranted. Moreover, we observed some residual effects of caffeine consumption after adjusting for caffeine metabolism, suggesting effects due to additional constituents of tea/coffee, or lifestyle/environmental factors on sleep. We found no clear evidence for effects of caffeine consumption or metabolism ratio on sleep duration or insomnia. Finally, we observed effects of being an evening/morning person on caffeine consumption levels suggesting the presence of a sleep-caffeine-sleep behavioural cycle. Our findings suggest that the observed relationships between genetically predicted caffeine consumption, caffeine metabolism, and daytime alertness traits can potentially aid in catering dietary advice to individuals suffering from sleep disorders. However, weak strength of the caffeine metabolism instruments could have affected the reliability of our results and biased our results towards the null. Although we employed multiple weak instrument-robust methods to improve the robustness of our findings, we suggest that future studies use better-powered GWASs to validate them.

## Data Availability

All data (GWAS summary statistics) used in the study are publicly available from the GWAS Catalog at https://www.ebi.ac.uk/gwas/.

## Acknowledgements

This research was conducted using the UK Biobank Resource under Application Number 16391 and 81499. Quality Control filtering of the UK Biobank data was conducted by R. Mitchell, G. Hemani, T. Dudding, L. Corbin, S. Harrison, L. Paternoster as described in the published protocol (doi:10.5523/bris.1ovaau5sxunp2cv8rcy88688v). The MRC IEU UK Biobank GWAS pipeline was developed by B. Elsworth, R. Mitchell, C. Raistrick, L. Paternoster, G. Hemani, T. Gaunt (doi:10.5523/bris.1ovaau5sxunp2cv8rcy88688v).

## Funding

This work was supported by Cancer Research UK (grant number C18281/A29019). BW works in the Medical Research Council Biostatistics Unit at the University of Cambridge which is supported by the Medical Research Council and the University of Cambridge (grant code MC_UU_00040/1). JK (grant code MC_UU_00011/7) and RCR (grant code MC_UU_00011/1) work in the Medical Research Council Integrative Epidemiology Unit at the University of Bristol which is supported by the Medical Research Council and the University of Bristol. RCR is supported by the National Institute for Health and Care Research (NIHR) Oxford Health Biomedical Research Centre. The views expressed are those of the author(s) and not necessarily those of the NIHR or the Department of Health and Social Care. NIHR Oxford Health Biomedical Research Centre grant reference number: NIHR203316.

## Conflicts of interest

The authors declare no conflicts of interest related to this study.

## Notes

### Competing Interest Statement

The authors have declared no competing interest.

### Author Declarations

The study used only openly available GWAS summary statistics from the GWAS Catalog at https://www.ebi.ac.uk/gwas/.

### Summary of Updates

The Discussion and Conclusion sections have been updated to include a more detailed mechanistic interpretation of the results.

