## Supplementary Text for "Exploring the Relationship between Caffeine Consumption, Caffeine Metabolism, and Sleep Behaviours: A Mendelian Randomisation Study"

**Supplementary Material**

**Supplementary Methods**

*Data sources*

*UK Biobank*

As stated in the MRC IEU UK Biobank Genome-Wide Association Study (GWAS) pipeline documentation (1), UK Biobank (UKB) is a population-based health research resource comprising almost 500,000 individuals aged between 38 years and 73 years. Participants were recruited between the years 2006 and 2010 from across the UK (2). UKB primarily aims to identify factors contributing to human diseases in middle-aged and older adults. Participants provided various types of information, including demographics, health status, lifestyle measures, cognitive assessments, personality self-reports, and physical and mental health measures through questionnaires and interviews. Additionally, anthropometric measurements, blood pressure readings, and samples of blood, urine, and saliva were collected (data available at [www.ukbiobank.ac.uk](http://www.ukbiobank.ac.uk)) (3). Detailed description of the study design, participant characteristics, and quality control (QC) methods have been described in detail previously (4). UKB obtained ethical approval from the Research Ethics Committee (REC reference for UK Biobank is 11/NW/0382).

GWASs are primarily aimed at identifying genetic variants that predict a trait or disease and pinpoint genes or loci relevant to the trait’s aetiology (5-7). GWASs have identified genetic variants associated with caffeine consumption from tea/coffee (8) as well as sleep traits such as chronotype, daytime napping, daytime sleepiness, getting up in morning, sleeplessness, sleep duration, etc. in UKB (9–13). However, blood plasma levels of caffeine metabolites and caffeine metabolism rate/ratio have not yet been identified in UKB. Hence, we used genetic variants identified in a much smaller meta-GWAS included less the 10,000 individuals of European ancestry (14) and procured statistical power from the large sleep trait GWAS by using a two-sample multivariable mendelian randomisation (2S-MVMR) framework.

*Caffeine consumption GWASs*

We used summary statistics from GWAS conducted by Said et al. available at <https://doi.org/10.17632/d8nwkm7p9p.1> (8). The GWAS included 407,072 individuals of European ancestry in UKB. Caffeine consumption was measured as self-reported answers to the following questions:

1. “How many cups of coffee do you drink each day? (Include decaffeinated coffee).” (UKB data-field 1498).
2. “How many cups of tea do you drink each day? (Include black and green tea).” (UKB data-field 1488).

Caffeine consumption was calculated by multiplying a standard caffeine content per cup (30 mg for tea and 60 mg for coffee) to the reported number of consumed cups of tea and/or coffee. Total caffeine intake was calculated as the sum of caffeine consumption from both tea and coffee from participants who reported data on both. Participants who reported “Less than one” (-10), “Do not know” (-1), or “Prefer not to answer” (-3) were set to missing and participants who had non-missing phenotype and covariates data were used in performing GW analyses. A total of 362,316 individuals were included in GWAS for combined tea and coffee, 373,522 for coffee, and 395,866 for tea.

*Plasma caffeine and caffeine metabolism GWAS*

We used summary statistics from the meta-analysis conducted by Cornelis and colleagues available at <https://digitalhub.northwestern.edu/users/mcc340> (14). The meta-GWAS included 9876 participants of European ancestry from 6 cohorts, namely, the Prospective Study of the Vasculature in Uppsala Seniors (PIVUS) (15), the Study of Health in Pomerania TREND (SHIP-TREND) (16), the Swiss Kidney Project on Genes in Hypertension (SKIPOGH) (17), TwinGene (18), TwinsUK (19), and the Uppsala Longitudinal Study of Adult Men (ULSAM) (20). Study sample was comprised of fasting blood samples from adults. Plasma 13X, 17X, 37X, 137X, and 137U were profiled independently by each cohort. Since cohorts used different methods for normalising distribution of metabolites and 17X/137X, all data were rescaled to a distribution having a mean of 0 and SD of 1 prior to performing GW analyses. The formation of paraxanthine (17X) from caffeine (137X) is entirely exclusively catalysed by the CYP1A2 gene and is responsible for metabolism of almost 80% of ingested caffeine. Hence, this ratio of paraxanthine to caffeine (17X/137X) was included in the GWAS along with plasma levels of other metabolites (13X, 17X, 37X, 137X, and 137U).

*Sleep behaviour GWASs*

We used summary statistics from the large GWAS of self-reported sleep traits in UKB available at <https://sleep.hugeamp.org/dinspector.html?dataset=GWAS_UKBB_eu>. Phenotypes of interest were chronotype, daytime napping, daytime sleepiness, getting up in morning, insomnia, and sleep duration.

The chronotype and getting up in morning meta-GWASs including 451,454 participants of European ancestry in UKB and 1,248,100 participants of European ancestry in 23andMe due to public non-availability and time-constraints. We only included GWAS summary statistics from UKB individuals in our analyses and excluded GWAS summary data from 23andMe cohort due to public unavailability of summary data. Participant chronotype was self-reported. The measure of chronotype in UKB was defined as “Morning/evening person (chronotype)” in UKB (UKB data-field 1180). Chronotype was ascertained using responses to the question: “Do you consider yourself to be…” Possible response choices were restricted to “Definitely a ‘morning’ person” (coded as 0), “More a ‘morning’ than ‘evening’ person” (1), “More an ‘evening’ than a ‘morning’ person” (2), “Definitely an ‘evening’ person” (3), “Do not know”, “Prefer not to answer”. The measure of ease getting up in morning (UKB data-field 1170) in UKB was defined as “Getting up in morning” in UKB. Participant morning-ness was self-reported. Ease of getting up in the morning was ascertained using responses to the question: “On an average day, how easy do you find getting up in the morning?” Possible response choices were restricted to “Very easy”, “Fairly easy”, “Not very easy”, “Not at all easy”, “Do not know”, and “Prefer not to answer”. Participants who reported “Do not know” or “Prefer not to answer” were set to missing and participants who had non-missing phenotype and covariates data were treated as a continuous trait for performing GW analyses (10).

The daytime napping GWAS included 452,633 participants of European ancestry in UKB. Participant daytime napping (UKB data-field 1190) frequency was self-reported. Daytime napping frequency was measured as the response to the question: “Do you have a nap during the day?” Possible response choices were restricted to “Never/rarely”, “Sometimes”, “Usually”, and “Prefer not to answer”. “Prefer not to answer” responses were set to missing and all other responses were treated as a continuous trait when performing GW analyses (12).

The daytime sleepiness GWAS included 452,071 participants of European ancestry in UKB. Participant daytime sleepiness (UKB data-field 1220) was self-reported. Daytime dozing/sleeping was measured using responses to the question: “How likely are you to doze off or fall asleep during the daytime when you don’t mean to? (e.g. when working, reading or driving)” Possible response choices were restricted to “Never/rarely”, “sometimes”, “often”, “all of the time”, “do not know”, and “prefer not to answer”. Participants who reported “do not know” or “prefer not to answer” were set to missing. All other responses were treated as a continuous trait when performing GW analyses (11).

The insomnia GWAS included 453,379 participants of European ancestry in UKB. Participant sleeplessness/insomnia (UKB data-field 1200) were self-reported. Insomnia symptoms in UKB were assessed from responses to the question: “Do you have trouble falling asleep at night or do you wake up in the middle of the night?” Possible response choices were “never/rarely”, “sometimes”, “usually”, and “prefer not to answer”. Participants who responded “prefer not to answer” were set to missing. Other participants were dichotomised into controls (participants who responded “never/rarely”) and cases (participants who responded “sometimes” and “usually”) prior to performing GW analyses (13). Summary statistics were also obtained from two separate GWASs performed similarly including 1,331,010 individuals of European ancestry combined from UKB and 23andMe (21) and 386,533 individuals of European ancestry in the UKB (22).

The sleep duration GWAS included 446,118 participants of European ancestry in UKB. Participant sleep duration (UKB data-field 1160) was self-reported. To measure sleep duration, participants were asked the question: “About how many hours sleep do you get in every 24h? (please include naps).” Responses were recorded in hours. “Do not know” or “Prefer not to answer” responses were set to missing. Further, participants who reported extreme responses (less than 3 hours or more than 18 hours) and participants who self-reported use of any sleep-medication were excluded prior to performing GW analyses (9).

The accelerometer derived sleep behaviours GWAS included 103,711 individuals of European ancestry in UKB. Individuals were asked to wear a triaxial accelerometer device (Axivity AX3) for a continuous period of up to 7 days. Of these, 11,067 individuals were excluded for having data problems, poor wear time, poor calibration, or unable to calibrate activity data on the device, data recording errors, interrupted recording periods. Final GW analysis was performed on a maximum of 85,723 individuals. Accelerometer derived sleep duration (n=85449), L5 (n=85205) and M10 (n=85670), number of sleep episodes (n=84810) and sleep efficiency (n=84810), and diurnal inactivity (n=84757) were used as substitutes for self-reported sleep duration, chronotype, insomnia, and daytime napping respectively. Sleep duration was estimated based on the total sleep episodes (periods of at least 5 minutes) within a pre-defined sleep-period-time (SPT) window, L5 was defined as the 5-hour period with the minimum average acceleration, M10 as the midpoint of the most-active 10 hours of each day, number of nocturnal sleep episodes as the number of sleep episodes within the SPT-window, sleep efficiency as sleep duration divided by time elapsed between first inactivity bout start and last inactivity bout end, and diurnal inactivity was estimated by the total duration of inactivity bouts outside SPT-window. All traits were treated as continuous during GW analyses (23).

Additionally, we performed a stratified-GWAS of sleep behaviours stratified by caffeine consumption status (current/non-current) using the MRC IEU UKB GWAS pipeline. This pipeline contains the full cohort of successfully genotyped samples in UKB (n=488,377). Pre-imputation QC, phasing, and imputation are described elsewhere (3). The sample was restricted to individuals of European ancestry defined by a cluster analysis utilising first 4 PCs provided by UKB in the statistical software environment R. The analysis included a cluster of 464,708 samples (1). This pipeline utilises BOLT-LMM (v2.3) software package to account for relatedness and population stratification (first 10 PCs) via a linear mixed model (1,24). We also adjusted for sex and genotyping chip while performing GWAS of stratified sleep traits.

In our stratified-GWASs, except for daytime sleepiness and insomnia, all other sleep behaviours were treated as continuous outcomes when running GW analyses. Daytime sleepiness and insomnia were dichotomised prior to running GW analyses because there was not enough heritability (variation) present in the non-current consumers stratum after stratification of these two traits by caffeine consumption status.

*Statistical analyses*

*Univariable MR*

We used a two-sample mendelian randomisation (2S-MR) framework, specifically inverse variance weighted (IVW), MR-Egger, weighted mode, and weighted median models to assess total effects of caffeine per day (CPD) and caffeine metabolite ratio (CMR) (independently) on sleep behaviours. IVW is a fixed effects estimate where Wald ratio or ratio estimates are combined together in a fixed effects meta-analysis. Weight of each ratio is given by the inverse of the variance of SNP-outcome association. As in a traditional meta-analysis, each SNP (IV) is treated as an independent study and Wald ratios estimated for each SNP are subsequently meta-analysed under a fixed effects model (25,26). MR-Egger is a two-sample specific MR model that combines Wald ratio, or ratio estimates into a meta-regression giving an intercept term and a slope term. A non-null intercept term from MR-Egger regression is an indication of horizontal pleiotropy. However, the slope term provides a test for a causal effect and a consistent estimate of causal effect even when the intercept is non-zero. Thus, MR-Egger estimates directional pleiotropy adjusted causal effect (27–31). Weighted mode and weighted median methods are extensions of two-sample MR whereby the estimate is given by the mode and the 50^th^ percentile of the empirical density function of Wald ratio or ratio estimates respectively (27,32,33).

To test for bi-directionality of the relationship between caffeine intake and sleep behaviours, we conducted a 2S-MR to investigate causal effects of sleep traits on CPD and CMR (34). Bi-directional MR is only valid under the condition that no marginal association exists between two instruments, i.e., SNP for CPD is not in LD with SNP for sleep behaviour (r^2^>0.001) (35). Hence, instrumental SNPs were chosen while ensuring GW significance (p<5x10^-8^), biological plausibility, and independence. Monotonicity of associations between instruments and exposure were validated. Furthermore, data was harmonised to ensure two samples were identically coded, i.e., reflecting same effect allele, prior to performing MR analyses (35). MR analyses were performed using Steiger filtered SNPs to minimise bias due to reverse causal instruments (36).

*Multivariable MR*

We used 2S-MVMR to assess direct effect of caffeine metabolism on sleep for a constant CPD, and direct effect of CPD on sleep for a given CMR. We performed MVMR analyses using MVMR-IVW. We used MVMR-Egger to check for consistency with IVW results and ensure robustness (25–27,30,31,37).

We tested for violations of MR and MVMR assumptions. For both frameworks, we tested instrument-exposure strength using an F statistic. F statistic for MR and cF for MVMR exceeding 10 was used to indicate good instrument strength (38). We used weak instrument robust methods like Qhet (39), debias IVW (40), Grapple (41), GMM, and IVW ME (42) to ensure consistency of effect estimates in situations of weak instrument strength. Reliable estimates of causal effects of each exposure on outcome can be obtained in the presence of weak instruments and pleiotropy, by repurposing commonly used heterogeneity Q-statistic as an estimating equation (39,44). Debiased IVW estimator is a simple modification of IVW estimator robust to many weak instruments and does not require screening. Multivariable debiased IVW (MV-dIVW) estimator effectively reduces asymptotic bias from weak instruments in MV-IVW (40). GRAPPLE (Genome-wide mR Analysis under Pervasive PLEiotropy) is a comprehensive MR framework to analyse causal effects of a target risk factor with heterogeneous genetic instruments and identify possible pleiotropic patterns in data. GRAPPLE uses summary statistics from GWASs to efficiently detect causal effects from MR using both strong and weak genetic instruments, detect existence of multiple pleiotropic pathways, adjust for confounding risk factors, as well as determine causal direction (41). GMM MR framework performs MVMR and produces robust causal inference in 2S-MVMR using generalised method of moments by accounting for overdispersion heterogeneity in genetic variant-outcome associations. Similarly, IVW ME framework mitigates bias in MR estimates due to causal differential measurement error by including the variable causing the error in an MVMR analysis (42).

We used a non-zero intercept term in MR-Egger regression as indicative of presence of horizontal pleiotropy (IV3 violation) (44). Finally, Cochran’s Q exceeding number of SNPs included in the model as instruments indicates heterogeneity (27).

We stratified sleep behaviour GWASs by caffeine consumption to explore effects of caffeine on sleep behaviours among both current and non-current consumers separately. We expect effect sizes of CPD on sleep behaviours to be more pronounced among current caffeine consumers of tea/coffee than among combined non-current/current consumers. Finally, we used evidence of effects among non-current consumers (which should not be possible because this serves as negative control) to indicate influence of pleiotropy or population stratification.

**Supplementary Results**

Among participants who reported sleep behaviours, current drinkers of tea and/or coffee largely outnumbered non-current drinkers as observed in chronotype (97.43% vs 2.57%). The distribution was similar (97.42% vs 2.58%) in daytime napping, daytime sleepiness, getting up in morning, insomnia, and sleep duration traits (**Supplementary Table 7**).

*Effects of CPD on sleep behaviours*

Similar to our primary results, we found no clear evidence of any effects of CPD on sleep traits in our pleiotropy robust and weak instrument robust MVMR methods (**Supplementary Figure 8**). We observed effect estimates similar to our primary analyses among tea consumers (**Supplementary Figure 15a**), coffee consumers (**Supplementary Figure 15b**), current caffeine (total tea and/or coffee) consumers (**Supplementary Figure 14**), and using accelerometer derived sleep traits (**Supplementary Figures 9a-f**). Further, the direction of the total effect of CPD on insomnia were not replicated across all our sensitivity analyses methods (OR_IVW-MR_=0.970, 95%CI [0.832,1.132], p=0.701; Jansen: OR_IVW-MR_=0.970, 95%CI [0.856,1.1], p=0.639; Watanabe: OR_IVW-MR_=1.074, 95%CI [0.95,1.215], p=0.253; SEFF: OR_IVW-MR_=1.007, 95%CI [1.001,1.013], p=0.019; NNSE: OR_IVW-MR_=0.849, 95%CI [0.627,1.15], p=0.290; Coffee: OR_IVW-MR_=1.020, 95%CI [0.768,1.354], p=0.891; Tea: OR_IVW-MR_=1.074, 95%CI [0.832,1.386], p=0.584; Current: OR_IVW-MR_=1.025, 95%CI [0.929,1.131], p=0.621) (**Supplementary Figures 2, 9d, 9f-h, 14e, 15**). Nonetheless, most of the confidence intervals included the null. However, we found evidence that higher CPD increased sleep efficiency (OR_IVW-MR_=1.007, 95%CI [1.001,1.013], p=0.019), though by a negligible amount (increase of 0.7% relative to those with lower CPD). Additionally, in our sample overlap-robust analyses, we observed more than a twofold increase in the estimated total effect of CPD on daytime napping (Corrected β_MRLap-IVW_ =-0.078, 95%CI [-0.134,-0.022], p=0.006) and daytime sleepiness (Corrected β_MRLap-IVW_=-0.106, 95%CI [-0.154,-0.058], p<0.001) (**Supplementary Figure 16**). However, the confidence intervals were consistent with our primary analyses. Likewise, we observed similar effect estimates as our primary results for all other sleep traits in our sample overlap-robust MR analysis (**Supplementary Figure 16**).

*Effects of CMR on sleep behaviours*

We found no clear evidence of any effects of CMR on sleep traits in our pleiotropy robust and weak instrument robust methods. This is similar to our primary results. However, effects estimates were not consistent across robust MR methods (**Supplementary Figure 8**). Additionally, total effect of CMR on chronotype from our primary analysis was not replicated in our sensitivity analyses (β_IVW-MR_=-0.044, 95%CI [-0.09,0.002], p=0.058; L5: β_IVW-MR_=0.035, 95%CI [-0.009,0.079], p=0.124; M10: β_IVW-MR_=0.055, 95%CI [-0.042,0.151], p=0.267; Current: β_IVW-MR_=0.034, 95%CI [0.01,0.057], p=0.005; Tea: β_IVW-MR_=-0.080, 95%CI [-0.195,0.036], p=0.177; Coffee: β_IVW-MR_=-0.047, 95%CI [-0.182,0.088], p=0.495) (**Supplementary Figures 2**, **9b-c, 14a, 15**). We observed effect estimates of CMR on sleep traits similar to our primary MR and MVMR results among tea consumers (**Supplementary Figure 15a**), coffee consumers (**Supplementary Figure 15b**), current caffeine (total tea and/or coffee) consumers (**Supplementary Figure 14**), and using accelerometer derived sleep traits (**Supplementary Figures 9a-f**). However, unlike in our primary analyses, we found evidence that faster CMR increased likelihood of being an evening person among current consumers (β_IVW-MR_=0.034, 95%CI [0.01,0.057], p=0.005) which attenuated after adjusting for CPD (β_IVW-MVMR_=0.008, 95%CI [-0.036,0.052], p=0.715) (**Supplementary Figure 14a**). We also replicated our primary results when substituting BPC for CMR (**Supplementary Figure 10**). Further, we observed similar effects of CMR on insomnia symptoms from different GWASs (**Supplementary Figure 9g-h**).

*Effects of sleep behaviours on caffeine*

We found evidence for a negative effect chronotype in our primary analysis (β_IVW-MR_=-0.044, 95%CI [-0.078,-0.010], p=0.011). However, this was not replicated in our pleiotropy robust MR analyses (β_MR-Egger_=-0.079, 95%CI [-0.188,0.031], p=0.160; β_MR-Weighted-median_=-0.047, 95%CI [-0.080,-0.014], p=0.006; β_MR-Weighted-mode_=-0.049, 95%CI [-0.119,0.020], p=0.166) (**Figure 4**). Additionally, in our pleiotropy robust MR methods, we found some evidence that experiencing insomnia symptoms decreased amount of CPD (β_MR-Egger_=-0.013, 95%CI [-0.127,0.101], p=0.822; β_MR-Weighted-median_=-0.029, 95%CI [-0.055,-0.003], p=0.030; β_MR-Weighted-mode_=-0.038, 95%CI [-0.078,0.002], p=0.074). Although we observed a similar effect estimate in IVW (β_IVW-MR_=-0.021, 95%CI [-0.054,0.012], p=0.219), there was not enough evidence for a conclusive negative effect (**Figure 4**). Similar to our primary MR results, we found no evidence for effect of daytime napping, daytime sleepiness, getting up in morning, or sleep duration on CPD in our sensitivity analyses (**Figure 4**). We also found no evidence for any effect of sleep behaviours on CMR in our sensitivity analyses (**Figure 5**). Additionally, we did not observe violations of bi-directional MR assumptions in funnel plots and leave-one-out analyses (**Supplementary Figures 3, 5,6, & 7**).

*Negative control*

Among non-current caffeine (total tea and/or coffee) consumers, we found to clear evidence for effect of CPD on chronotype (β_IVW-MR_=-0.025, 95%CI [-0.198,0.147], p=0.773; β_IVW-MVMR_=-0.097, 95%CI [-0.303,0.108], p=0.359), daytime napping (β_IVW-MR_=0.007, 95%CI [-0.109,0.123], p=0.907; β_IVW-MVMR_=0.042, 95%CI [-0.095,0.18], p=0.548), daytime sleepiness (OR_IVW-MR_=1.224, 95%CI [0.532,2.817], p=0.634; OR_IVW-MVMR_=1.378, 95%CI [0.58,3.28], p=0.473), getting up in morning (β_IVW-MR_=-0.054, 95%CI [-0.19,0.082], p=0.436; β_IVW-MVMR_=-0.025, 95%CI [-0.174,0.124], p=0.744), insomnia (OR_IVW-MR_=1.165, 95%CI [0.804,1.688], p=0.419; OR_IVW-MVMR_=1.233, 95%CI [0.824,1.847], p=0.310), and sleep duration (β_IVW-MR_=-0.077, 95%CI [-0.282,0.128], p=0.461; β_IVW-MVMR_=0.018, 95%CI [-0.215,0.251], p=0.879). Similarly, we found no evidence for effect of CMR on chronotype (β_IVW-MR_=0.034, 95%CI [-0.092,0.16], p=0.596; β_IVW-MVMR_=0.099, 95%CI [-0.04,0.239], p=0.171), daytime napping (β_IVW-MR_=-0.007, 95%CI [-0.176,0.162], p=0.936; β_IVW-MVMR_=-0.074, 95%CI [-0.167,0.019], p=0.128), daytime sleepiness (OR_IVW-MR_=0.901, 95%CI [0.486,1.667], p=0.739; OR_IVW-MVMR_=0.696, 95%CI [0.386,1.254], p=0.234), getting up in morning (β_IVW-MR_=-0.067, 95%CI [-0.167,0.034], p=0.192; β_IVW-MVMR_=-0.047, 95%CI [-0.148,0.054], p=0.369), insomnia (OR_IVW-MR_=0.956, 95%CI [0.662,1.381], p=0.810; OR_IVW-MVMR_=0.996, 95%CI [0.757,1.31], p=0.9877), and sleep duration (β_IVW-MR_=-0.114, 95%CI [-0.268,0.04], p=0.148; β_IVW-MVMR_=-0.110, 95%CI [-0.268,0.048], p=0.178) among non-current caffeine consumers (**Supplementary Figure 12**).

**Supplementary Tables**

**Supplementary Table 1.** Strengthening the Reporting of Observational Studies in Epidemiology using Mendelian Randomization (STROBE-MR) checklist of recommended items to address in reports of Mendelian randomisation studies.

| **Item No.** | **Section** | **Checklist item** | **Page No.** | **Relevant text from manuscript** |
| --- | --- | --- | --- | --- |
| 1 | **TITLE and ABSTRACT** | Indicate Mendelian randomization (MR) as the study’s design in the title and/or the abstract if that is a main purpose of the study | 1, 2 | Title page, Abstract |
|  | **INTRODUCTION** |  |  |  |
| 2 | **Background** | Explain the scientific background and rationale for the reported study. What is the exposure? Is a potential causal relationship between exposure and outcome plausible? Justify why MR is a helpful method to address the study question | 3-4 | Introduction section |
| 3 | **Objectives** | State specific objectives clearly, including pre-specified causal hypotheses (if any). State that MR is a method that, under specific assumptions, intends to estimate causal effects | 4 | Final paragraph |
|  | **METHODS** |  |  |  |
| 4 | **Study design and data sources** | Present key elements of the study design early in the article. Consider including a table listing sources of data for all phases of the study. For each data source contributing to the analysis, describe the following: | 4 | Data Sources section |
|  | a) | Setting: Describe the study design and the underlying population, if possible. Describe the setting, locations, and relevant dates, including periods of recruitment, exposure, follow-up, and data collection, when available. | 4-5 | Data Sources section/supplement |
|  | b) | Participants: Give the eligibility criteria, and the sources and methods of selection of participants. Report the sample size, and whether any power or sample size calculations were carried out prior to the main analysis | 4-5 | Data Sources section/supplement |
|  | c) | Describe measurement, quality control and selection of genetic variants | 5-6 | Statistical Analysis section/supplement |
|  | d) | For each exposure, outcome, and other relevant variables, describe methods of assessment and diagnostic criteria for diseases | 4-5 | Data Sources section/supplement |
|  | e) | Provide details of ethics committee approval and participant informed consent, if relevant | NA |  |
| 5 | **Assumptions** | Explicitly state the three core IV assumptions for the main analysis (relevance, independence and exclusion restriction) as well assumptions for any additional or sensitivity analysis | 4 | Introduction section |
| 6 | **Statistical methods: main analysis** | Describe statistical methods and statistics used | 5-6 | Statistical Analysis section/supplement |
|  | a) | Describe how quantitative variables were handled in the analyses (i.e., scale, units, model) | 4-5 | Data Sources section/supplement |
|  | b) | Describe how genetic variants were handled in the analyses and, if applicable, how their weights were selected | Supplementary methods |  |
|  | c) | Describe the MR estimator (e.g. two-stage least squares, Wald ratio) and related statistics. Detail the included covariates and, in case of two-sample MR, whether the same covariate set was used for adjustment in the two samples | Supplementary methods |  |
|  | d) | Explain how missing data were addressed | NA |  |
|  | e) | If applicable, indicate how multiple testing was addressed | NA |  |
| 7 | **Assessment of assumptions** | Describe any methods or prior knowledge used to assess the assumptions or justify their validity | 5-6 | Statistical Analysis section/supplement |
| 8 | **Sensitivity analyses and additional analyses** | Describe any sensitivity analyses or additional analyses performed (e.g. comparison of effect estimates from different approaches, independent replication, bias analytic techniques, validation of instruments, simulations) | 6-7 | Sensitivity analyses section/supplement |
| 9 | **Software and pre-registration** |  |  |  |
|  | a) | Name statistical software and package(s), including version and settings used | 7 | Final paragraph of Statistical Analysis section |
|  | b) | State whether the study protocol and details were pre-registered (as well as when and where) | 7 | Final paragraph of Statistical Analysis section |
|  | **RESULTS** |  |  |  |
| 10 | **Descriptive data** |  |  |  |
|  | a) | Report the numbers of individuals at each stage of included studies and reasons for exclusion. Consider use of a flow diagram | NA |  |
|  | b) | Report summary statistics for phenotypic exposure(s), outcome(s), and other relevant variables (e.g. means, SDs, proportions) | NA |  |
|  | c) | If the data sources include meta-analyses of previous studies, provide the assessments of heterogeneity across these studies | NA |  |
|  | d) | For two-sample MR:  i.  Provide justification of the similarity of the genetic variant-exposure associations between the exposure and outcome samples  ii.  Provide information on the number of individuals who overlap between the exposure and outcome studies | NA |  |
| 11 | **Main results** |  |  |  |
|  | a) | Report the associations between genetic variant and exposure, and between genetic variant and outcome, preferably on an interpretable scale | 7 | Instrument strength section |
|  | b) | Report MR estimates of the relationship between exposure and outcome, and the measures of uncertainty from the MR analysis, on an interpretable scale, such as odds ratio or relative risk per SD difference | 7-9 | Results section |
|  | c) | If relevant, consider translating estimates of relative risk into absolute risk for a meaningful time period | NA |  |
|  | d) | Consider plots to visualize results (e.g. forest plot, scatterplot of associations between genetic variants and outcome versus between genetic variants and exposure) | Figures 3 & 4 |  |
| 12 | **Assessment of assumptions** |  |  |  |
|  | a) | Report the assessment of the validity of the assumptions | 7, 9 | Instrument strength section, final paragraph of primary results section/supplement |
|  | b) | Report any additional statistics (e.g., assessments of heterogeneity across genetic variants, such as *I^2^*, Q statistic or E-value) | 7 | Instrument strength section/supplement |
| 13 | **Sensitivity analyses and additional analyses** |  |  |  |
|  | a) | Report any sensitivity analyses to assess the robustness of the main results to violations of the assumptions | 9 | Sensitivity analyses section/supplement |
|  | b) | Report results from other sensitivity analyses or additional analyses | 9 | Sensitivity analyses section/supplement |
|  | c) | Report any assessment of direction of causal relationship (e.g., bidirectional MR) | 8-9 | Final section (Effects of sleep behaviours on caffeine consumption and metabolism) of primary results |
|  | d) | When relevant, report and compare with estimates from non-MR analyses | NA |  |
|  | e) | Consider additional plots to visualize results (e.g., leave-one-out analyses) | Supplementary Figures |  |
|  | **DISCUSSION** |  |  |  |
| 14 | **Key results** | Summarize key results with reference to study objectives | 10 | First part of the Discussion section |
| 15 | **Limitations** | Discuss limitations of the study, taking into account the validity of the IV assumptions, other sources of potential bias, and imprecision. Discuss both direction and magnitude of any potential bias and any efforts to address them | 11 | Strengths and limitations section |
| 16 | **Interpretation** |  |  |  |
|  | a) | Meaning: Give a cautious overall interpretation of results in the context of their limitations and in comparison with other studies | 12 | Implications section & conclusion section |
|  | b) | Mechanism: Discuss underlying biological mechanisms that could drive a potential causal relationship between the investigated exposure and the outcome, and whether the gene-environment equivalence assumption is reasonable. Use causal language carefully, clarifying that IV estimates may provide causal effects only under certain assumptions | 12 | Conclusion section |
|  | c) | Clinical relevance: Discuss whether the results have clinical or public policy relevance, and to what extent they inform effect sizes of possible interventions | 12 | Implication section |
| 17 | **Generalizability** | Discuss the generalizability of the study results (a) to other populations, (b) across other exposure periods/timings, and (c) across other levels of exposure | 11 | Strengths and limitations section |
|  | **OTHER INFORMATION** |  |  |  |
| 18 | **Funding** | Describe sources of funding and the role of funders in the present study and, if applicable, sources of funding for the databases and original study or studies on which the present study is based | 1 | Acknowledgements section |
| 19 | **Data and data sharing** | Provide the data used to perform all analyses or report where and how the data can be accessed, and reference these sources in the article. Provide the statistical code needed to reproduce the results in the article, or report whether the code is publicly accessible and if so, where | Supplementary methods |  |
| 20 | **Conflicts of Interest** | All authors should declare all potential conflicts of interest | Title page |  |

This checklist is copyrighted by the Equator Network under the Creative Commons Attribution 3.0 Unported (CC BY 3.0) license.

**Supplementary Table 2.** F statistics for MR of caffeine consumption (CPD) and caffeine metabolism (CMR) on sleep behaviours

| **Exposure** | **Chronotype** | **Daytime napping** | **Daytime sleepiness** | **Getting up in morning** | **Insomnia** | **Sleep duration** |
| --- | --- | --- | --- | --- | --- | --- |
| CPD | 106.09 | 96.08 | 105.05 | 82.04 | 105.05 | 105.05 |
| CMR | 84.97 | 84.97 | 84.97 | 84.97 | 84.97 | 84.97 |

**Supplementary Table 3.** Conditional F statistics for MVMR of caffeine consumption (CPD) and caffeine metabolism (CMR) on sleep behaviours

| **Exposure** | **Chronotype** | **Daytime napping** | **Daytime sleepiness** | **Getting up in morning** | **Insomnia** | **Sleep duration** |
| --- | --- | --- | --- | --- | --- | --- |
| CPD | 11.67 | 10.89 | 11.67 | 16.58 | 11.67 | 11.67 |
| CMR | 5.01 | 4.71 | 5.01 | 4.84 | 5.01 | 5.01 |

**Supplementary Table 4.** Q and I^2^ statistics for MR and MVMR of caffeine consumption (CPD) and caffeine metabolism (CMR) on sleep behaviours

| **Exposure** | **MR (CPD)** | | **MR (CMR)** | | **MVMR** | |
| --- | --- | --- | --- | --- | --- | --- |
|  | **Q** | **I^2^ (%)** | **Q** | **I^2^ (%)** | **Q** | **I^2^ (%)** |
| Chronotype | 125.41 | 69.70 | 7.68 | 73.95 | 240.41 | 82.95 |
| Daytime napping | 127.50 | 62.35 | 3.03 | 33.89 | 131.72 | 66.59 |
| Daytime sleepiness | 120.13 | 64.21 | 25.13 | 92.04 | 106.29 | 61.43 |
| Getting up in morning | 81.07 | 55.59 | 5.68 | 64.80 | 144.14 | 73.64 |
| Insomnia | 105.84 | 62.21 | 0.24 | 0 | 151.10 | 72.87 |
| Sleep duration | 82.96 | 51.17 | 1.32 | 0 | 111.09 | 63.09 |

**Supplementary Table 5.** F statistics for MR of sleep behaviours on caffeine consumption (CPD) and caffeine metabolism (CMR)

| **Exposure** | **F statistic** | |
| --- | --- | --- |
|  | **CPD** | **CMR** |
| Chronotype | 47.59 | 51.41 |
| Daytime napping | 46.44 | 52.79 |
| Daytime sleepiness | 42.16 | 44.79 |
| Getting up in morning | 42.42 | 44.29 |
| Insomnia | 42.55 | 45.74 |
| Sleep duration | 40.76 | 43.07 |

**Supplementary Table 6.** Q and I^2^ statistics for MR of sleep behaviours on caffeine consumption (CPD) and caffeine metabolism (CMR)

| **Exposure** | **CPD** | | **CMR** | |
| --- | --- | --- | --- | --- |
|  | **Q** | **I^2^ (%)** | **Q** | **I^2^ (%)** |
| Chronotype | 416.19 | 64.68 | 24.59 | 0 |
| Daytime napping | 293.32 | 64.88 | 25.17 | 0 |
| Daytime sleepiness | 138.26 | 74.69 | 9.94 | 0 |
| Getting up in morning | 242.84 | 79 | 1.025 | 0 |
| Insomnia | 187.19 | 80.23 | 6.29 | 0 |
| Sleep duration | 166.12 | 60.87 | 8.28 | 0 |

**Supplementary Table 7.** UK Biobank participants with sleep behaviour data stratified by caffeine consumption status.

| **Caffeine consumption status** | **Chronotype** | **Daytime napping** | **Daytime sleepiness** | **Getting up in morning** | **Insomnia** | **Sleep duration** |
| --- | --- | --- | --- | --- | --- | --- |
| Non-current | 10047 (2.57%) | 11241 (2.58%) | 11206 (2.58%) | 11217 (2.58%) | 11252 (2.58%) | 11155 (2.58%) |
| Current | 380410 (97.43%) | 424901 (97.42%) | 423462 (97.42%) | 424518 (97.42%) | 425161 (97.42%) | 423265 (97.42%) |

**Supplementary Figures**

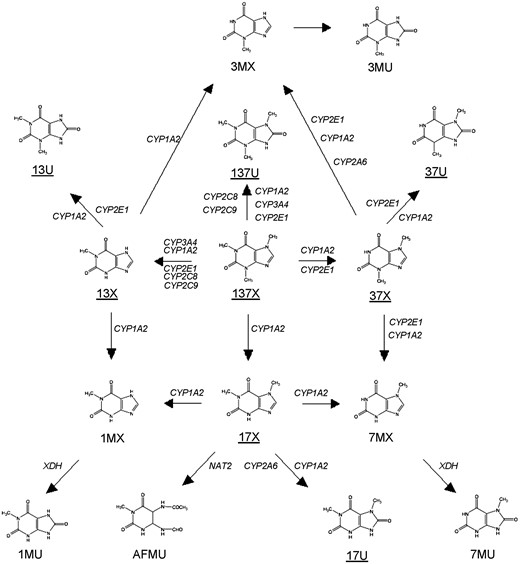

**Supplementary Figure 1.** Current understanding of the metabolic pathways of caffeine adapted from Cornelis et. al. (14).

Directional arrows signify metabolites and sub-metabolites produced from metabolism of caffeine and its metabolites. 137X: 1,3,7-trimethylxanthine (caffeine); 17X: 1,7-dimethylxanthine (paraxanthine); 13X: 1,3-dimethylxanthine (theophylline); 37X: 3,7-dimethylxanthine (theobromine); 137U:1,3,7-trimethyluric acid; 17U: 1,7-dimethyluric acid; 13U:1,3-dimethyluric acid; 37U: 3,7-dimethyluric acid; 1MX: 1-methylxanthine; 3MX: 3-methylxanthine; 7MX: 7-methylxanthine; 1MU: 1-methyluric acid; 3MU: 3-methyluric acid; 7MU: 7-methyluric acid; AFMU: 5-acetylamino-6-formylamino-3-methyluracil.

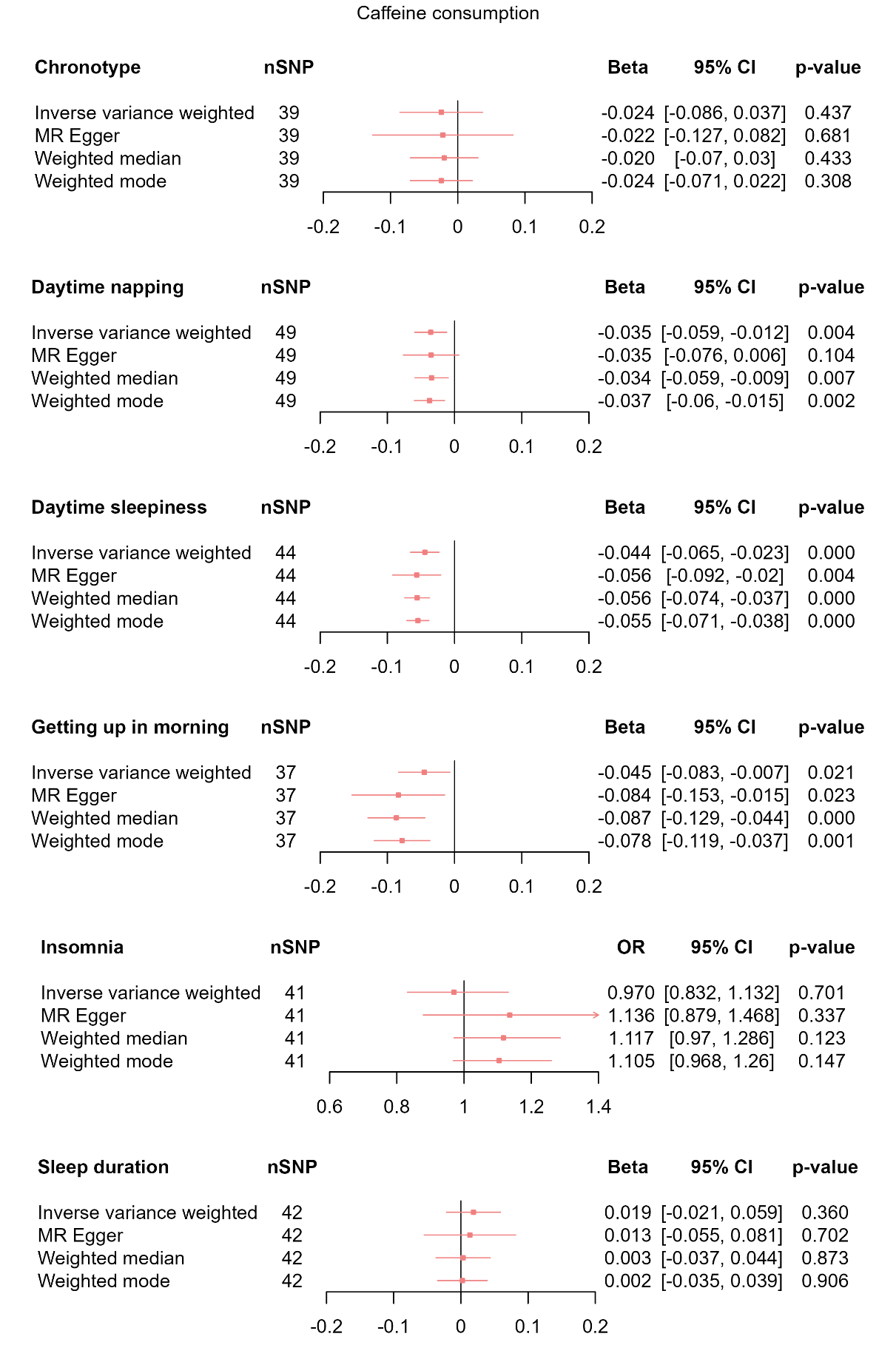

**Supplementary Figure 2.** MR results of total effect of caffeine consumption on sleep behaviours.

Red squares represent the estimated effect sizes (betas and OR) for each individual estimation method. The red horizontal lines represent the 95% confidence intervals (95% CI) for the estimated effects. The black vertical line represents the point of no effect. “nSNP” gives the number of single nucleotide polymorphisms (SNP) used in each estimation method.

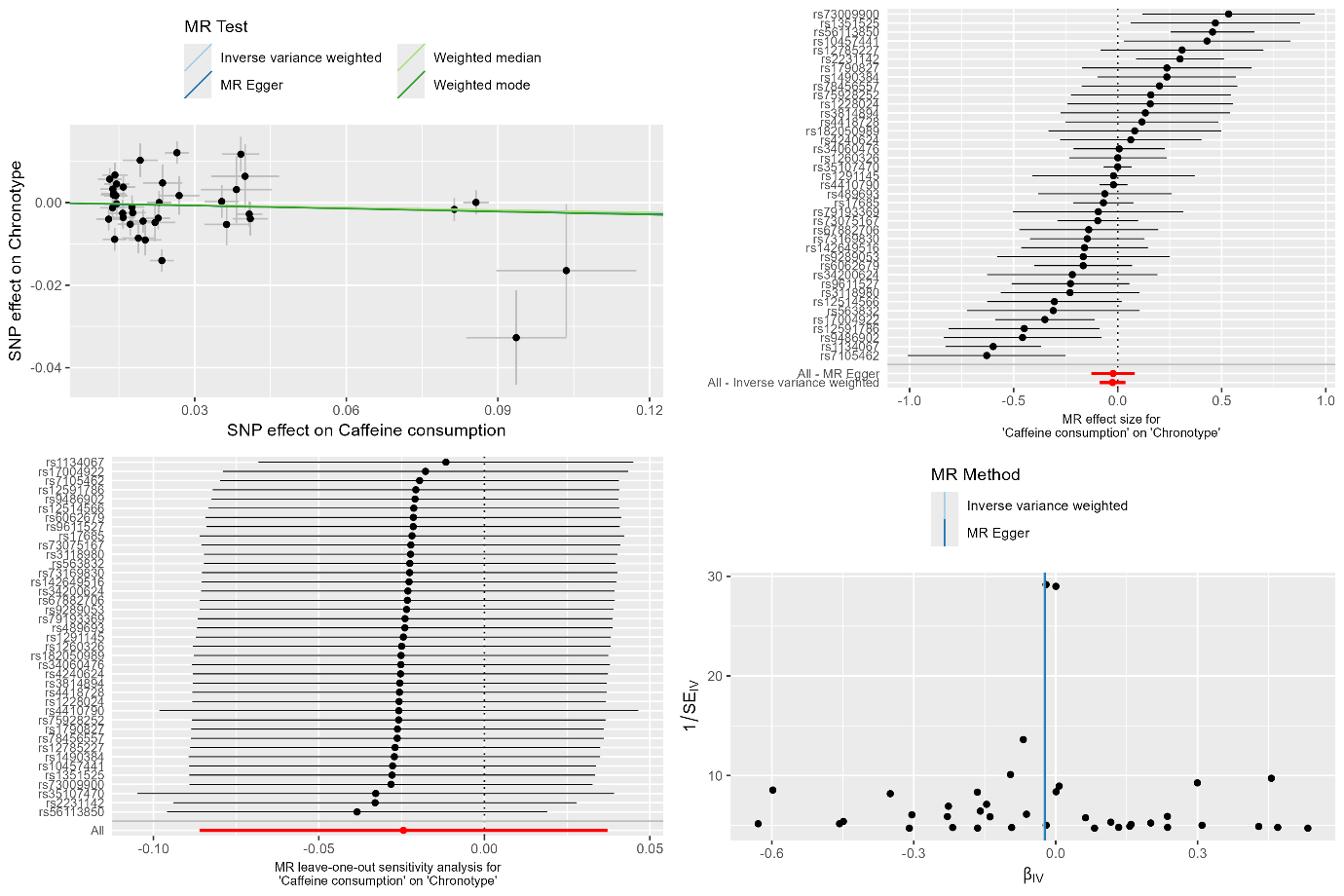

**Supplementary Figure 3a.** MR results and sensitivity analyses of caffeine consumption on chronotype.

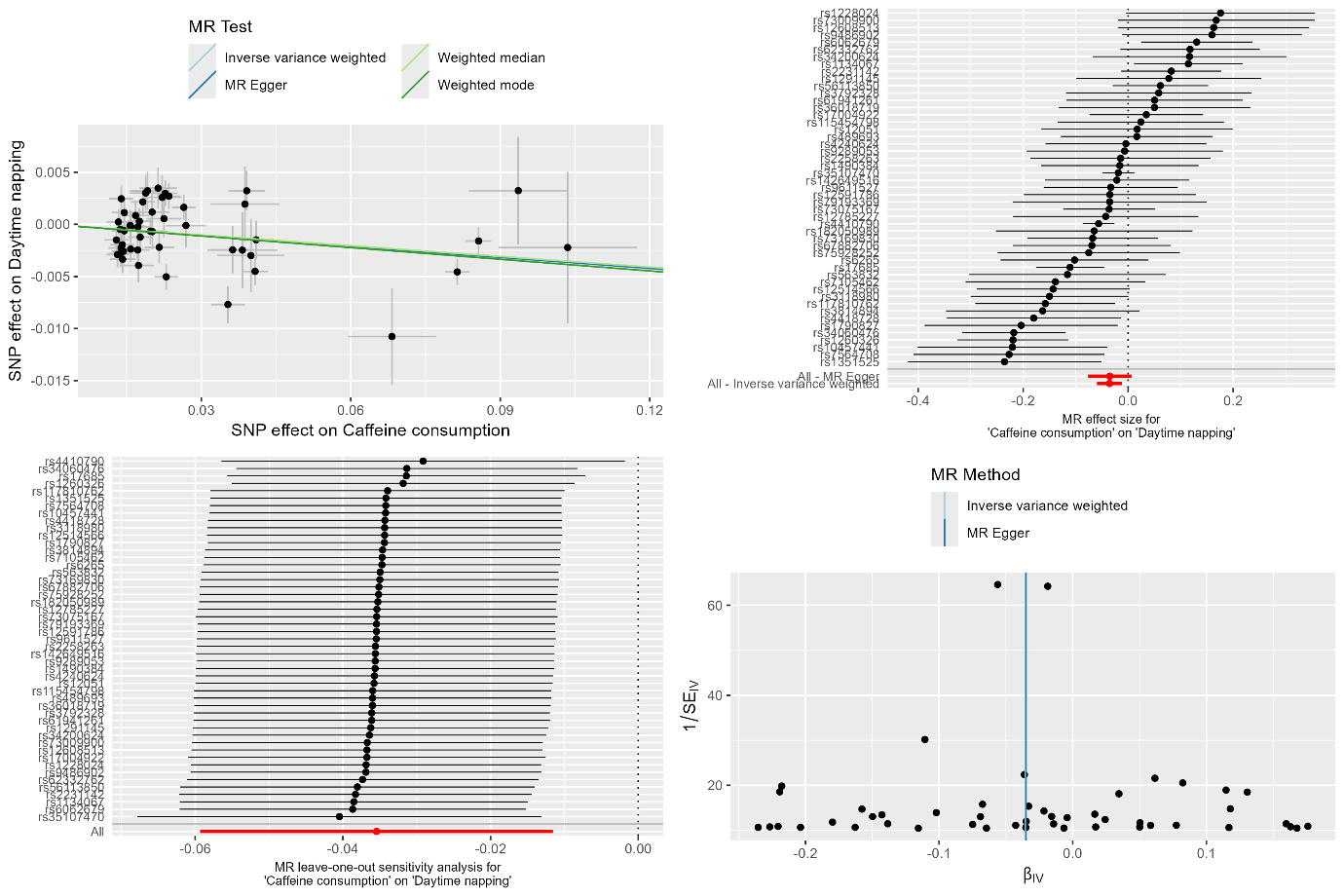

**Supplementary Figure 3b.** MR results and sensitivity analyses of caffeine consumption on daytime napping.

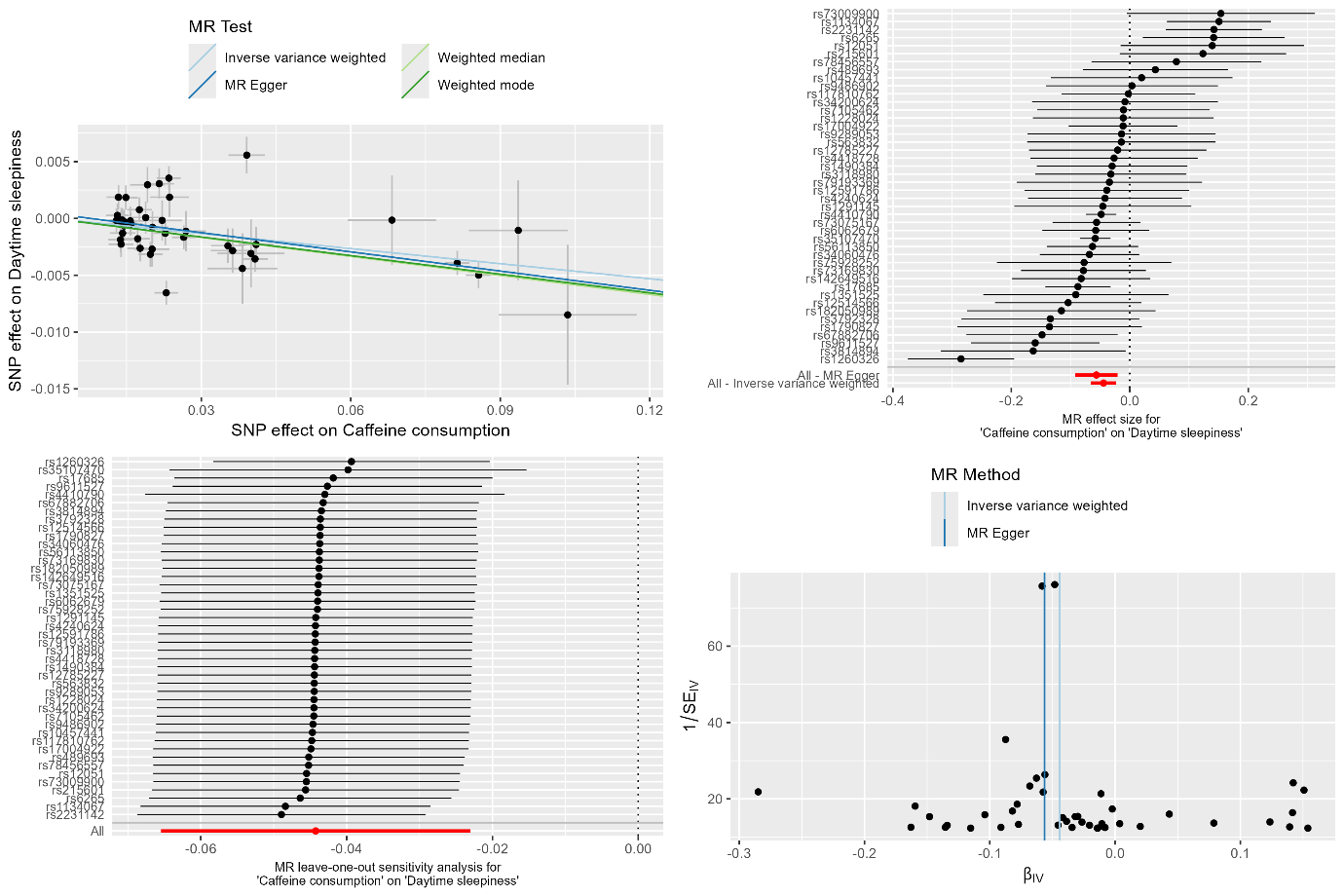

**Supplementary Figure 3c.** MR results and sensitivity analyses of caffeine consumption on daytime sleepiness.

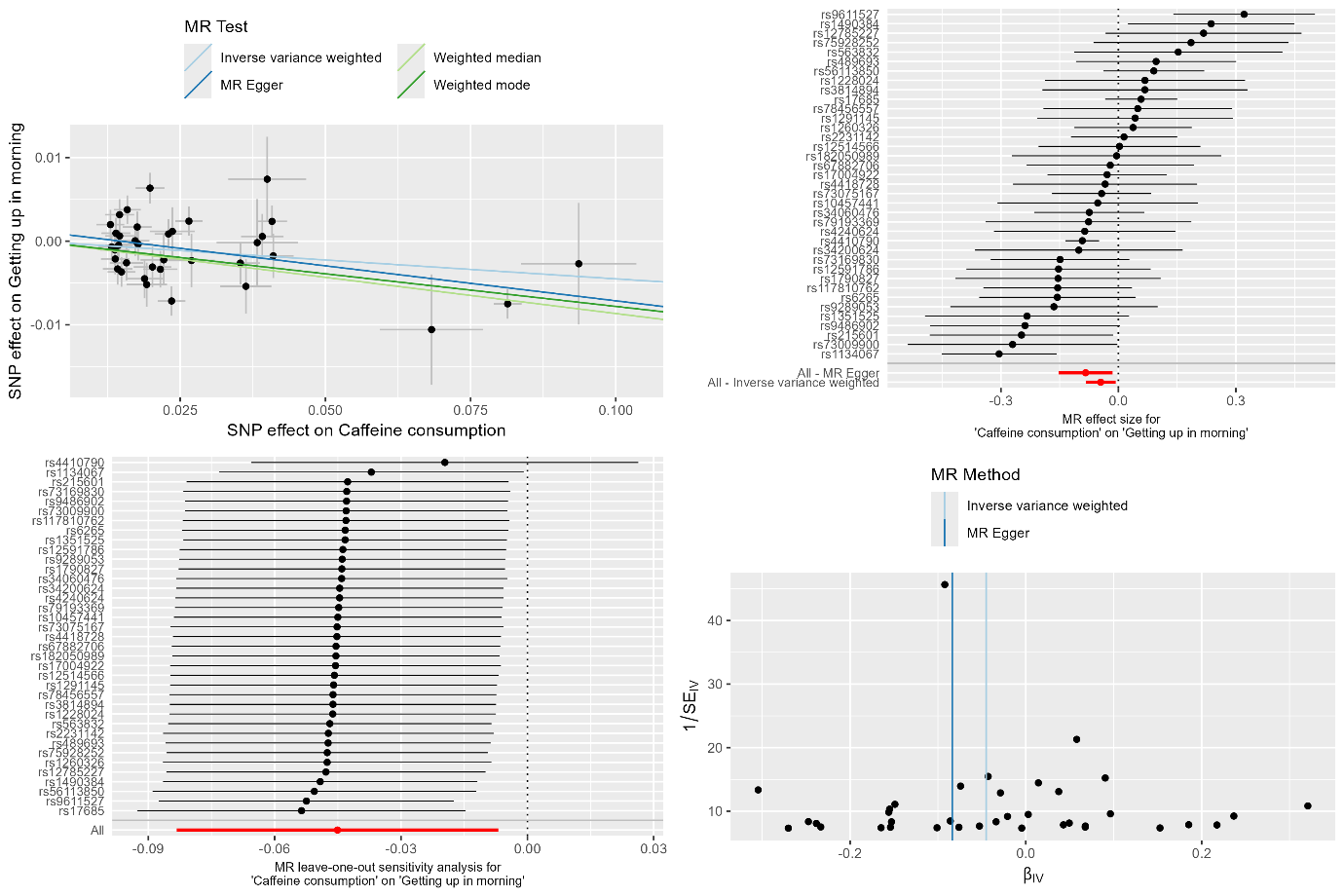

**Supplementary Figure 3d.** MR results and sensitivity analyses of caffeine consumption on getting up in morning.

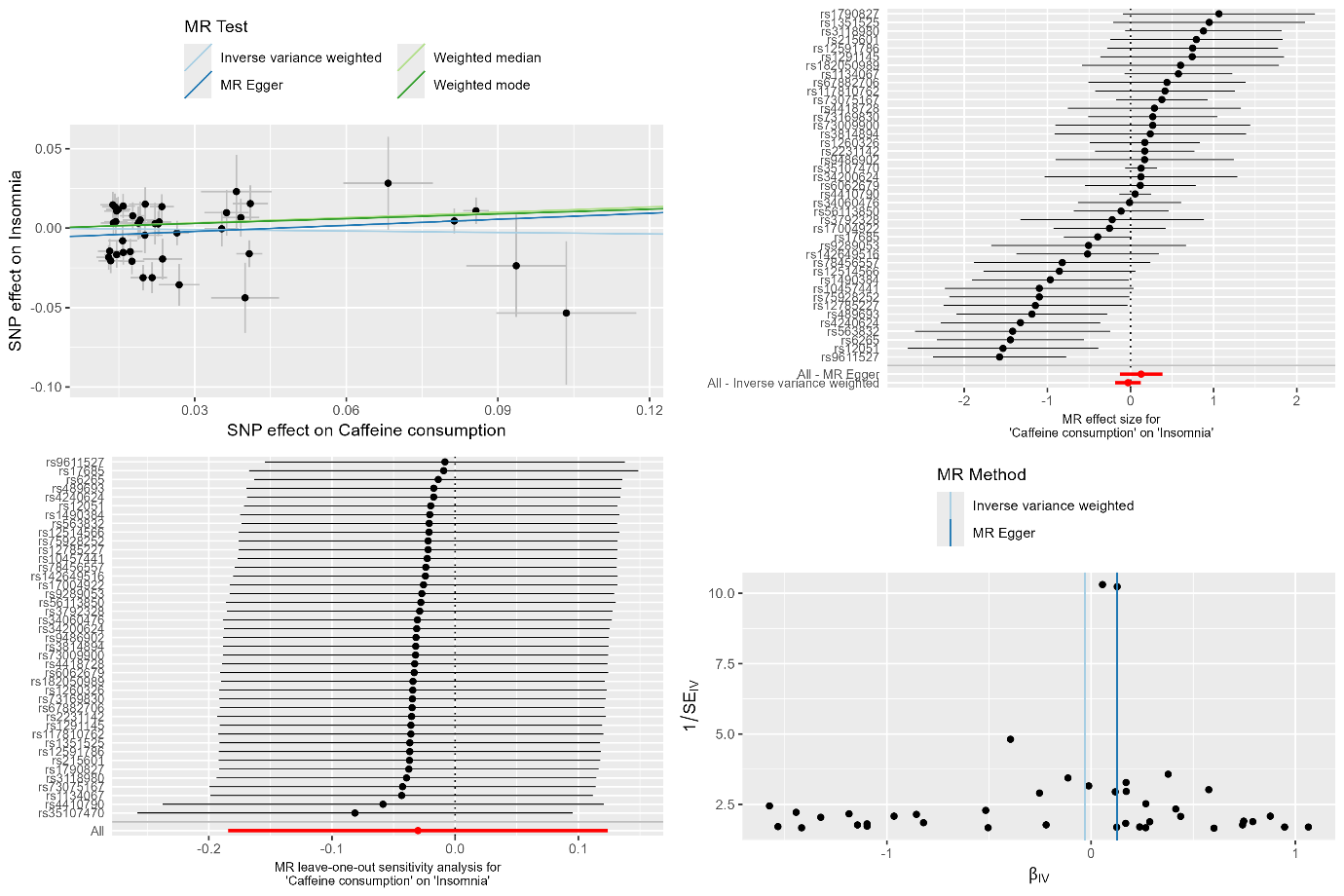

**Supplementary Figure 3e.** MR results and sensitivity analyses of caffeine consumption on insomnia.

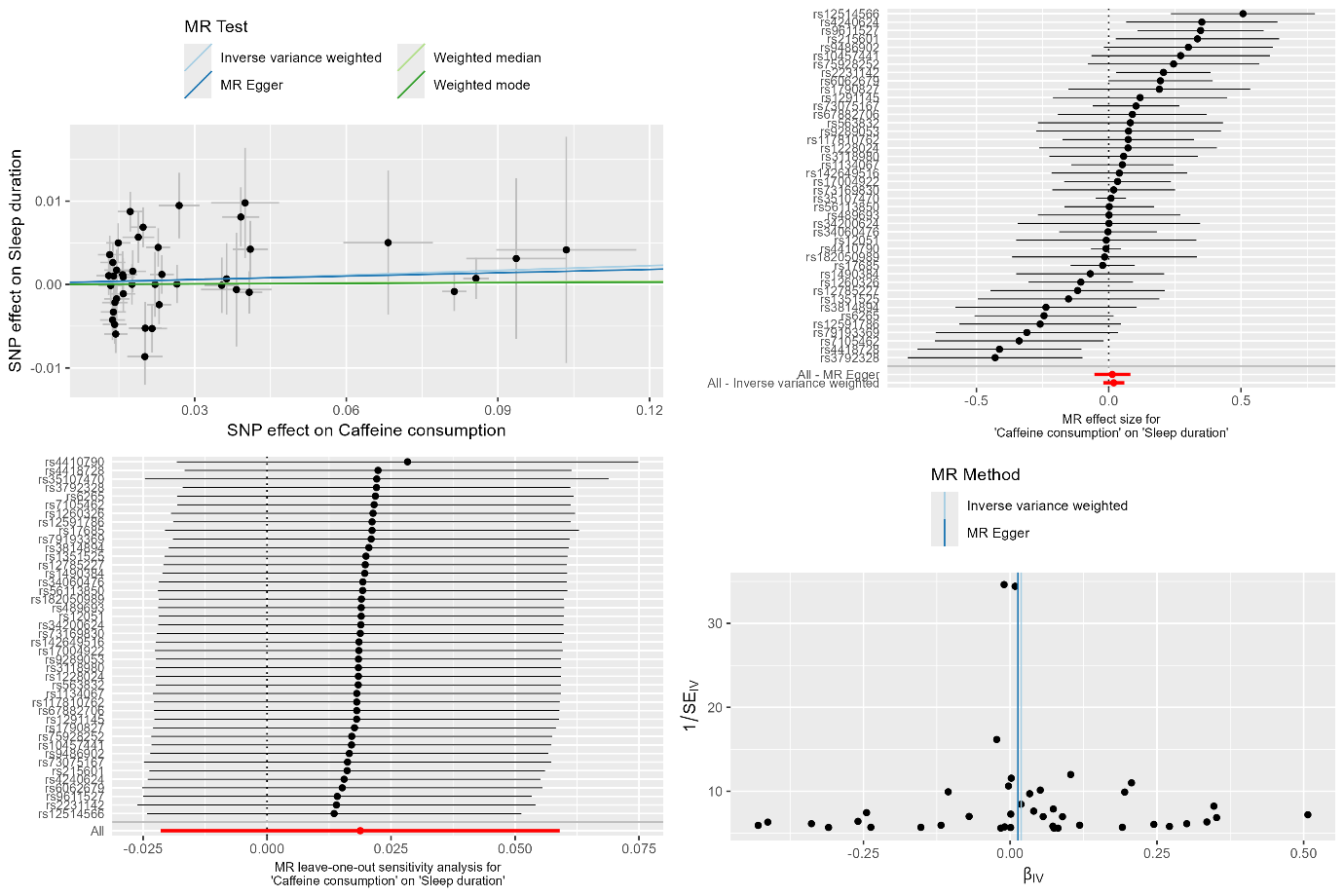

**Supplementary Figure 3f.** MR results and sensitivity analyses of caffeine consumption on sleep duration.

Top left: MR results. Black dots represent individual SNP effects. Vertical and horizontal black lines correspond to standard deviations of effects. Top right: Single SNP MR. Black dots represent individual SNP effects arranged in decreasing effect sizes. Horizontal black lines illustrate 95% confidence intervals for SNP effects. Red dots represent overall effect. Horizontal red lines illustrate 95% confidence intervals for overall effect. Bottom left: Leave-one-out MR results. Black dots represent overall effects when corresponding SNP is excluded from analysis. Horizontal black lines illustrate 95% confidence intervals for each leave-one-out MR. Red dots represent total effect when including all SNPs. Horizontal red lines illustrate 95% confidence intervals for overall effect of all SNPs. Bottom right: Funnel plot. Black dots represent individual SNP effects.

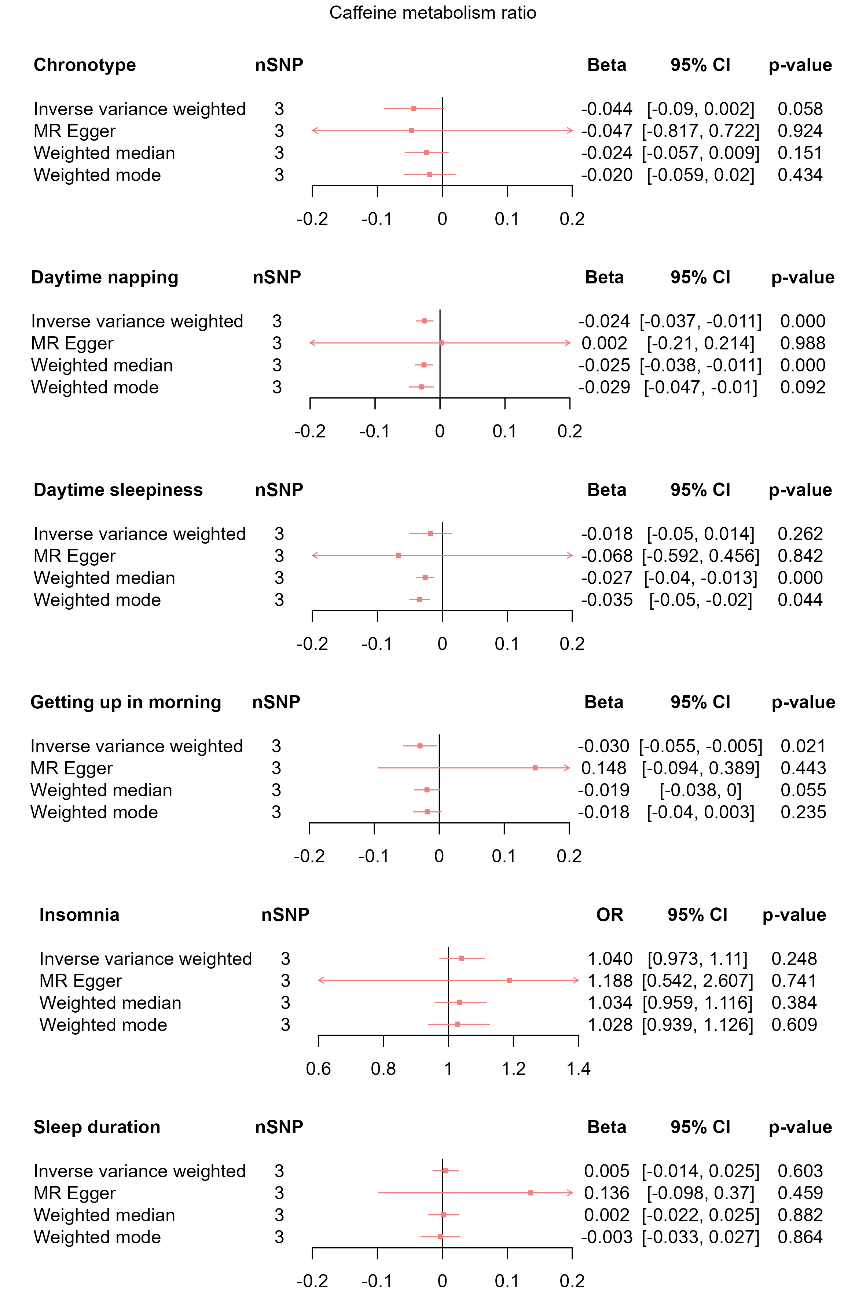

**Supplementary Figure 4.** MR results of total effect of caffeine metabolism ratio on sleep behaviours.

Red squares represent the estimated effect sizes (betas and ORs) for each individual estimation method. The red horizontal lines represent the 95% confidence intervals (95% CI) for the estimated effects. The black vertical line represents the point of no effect. “nSNP” gives the number of single nucleotide polymorphisms (SNP) used in each estimation method.

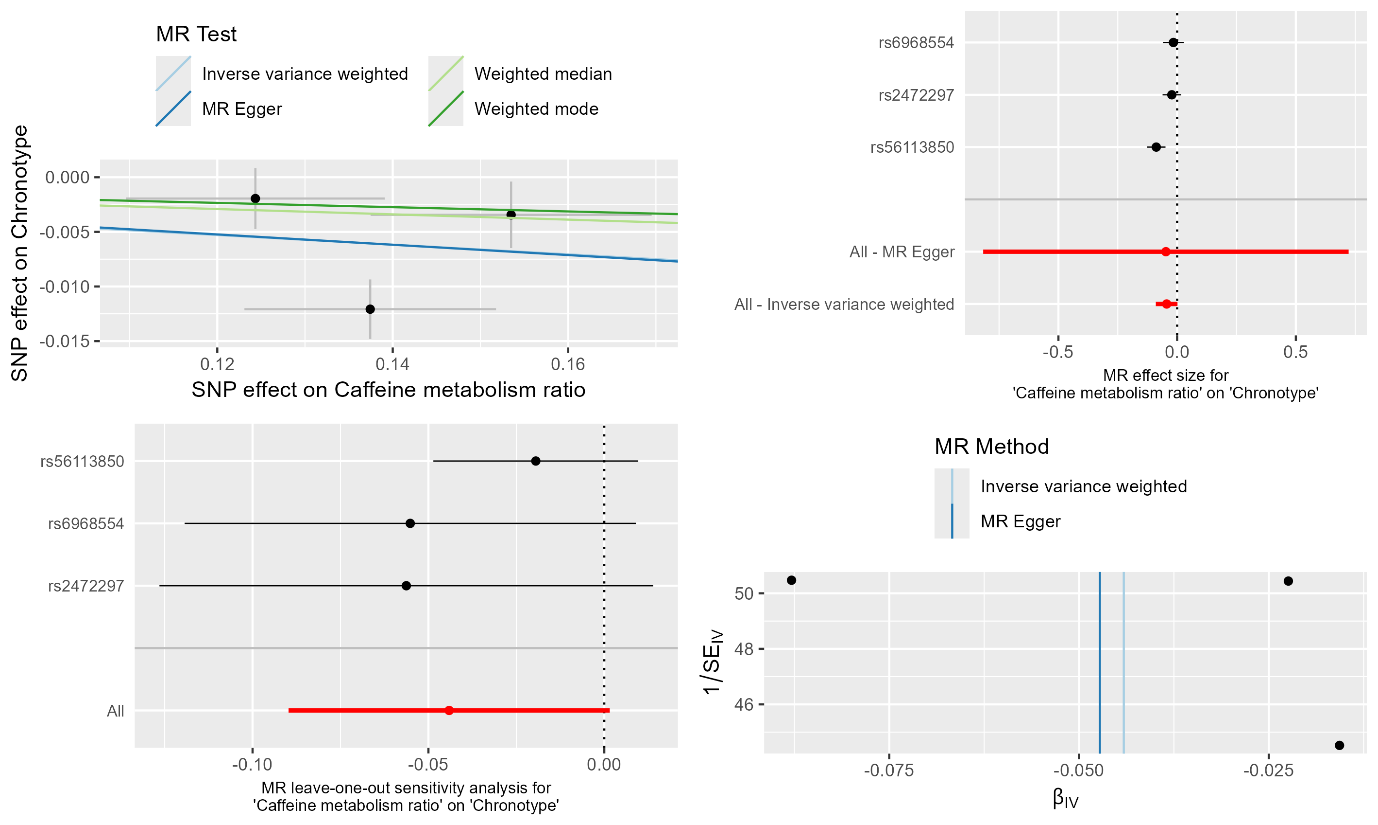

**Supplementary Figure 5a.** MR results and sensitivity analyses of caffeine metabolism ratio on chronotype.

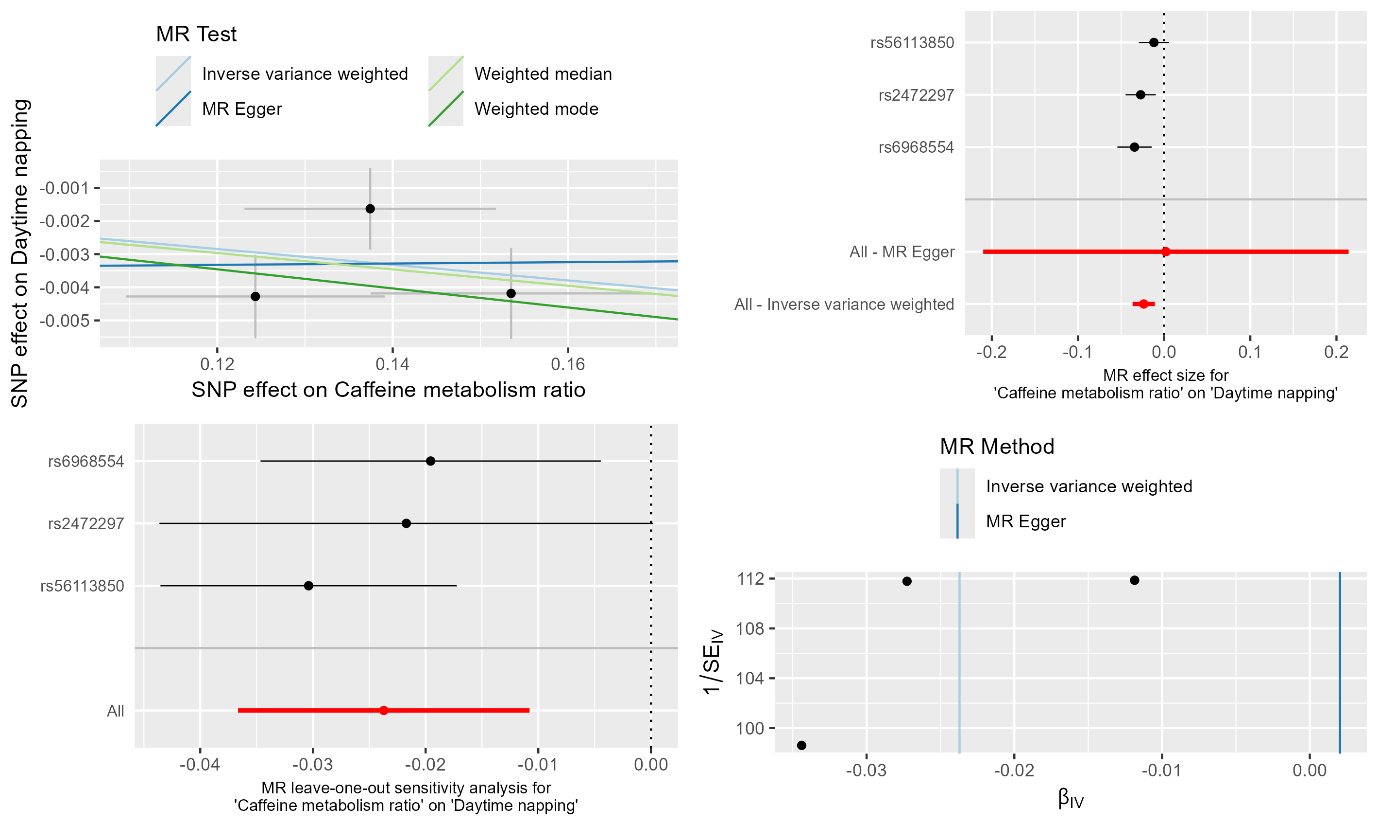

**Supplementary Figure 5b.** MR results and sensitivity analyses of caffeine metabolism ratio on daytime napping.

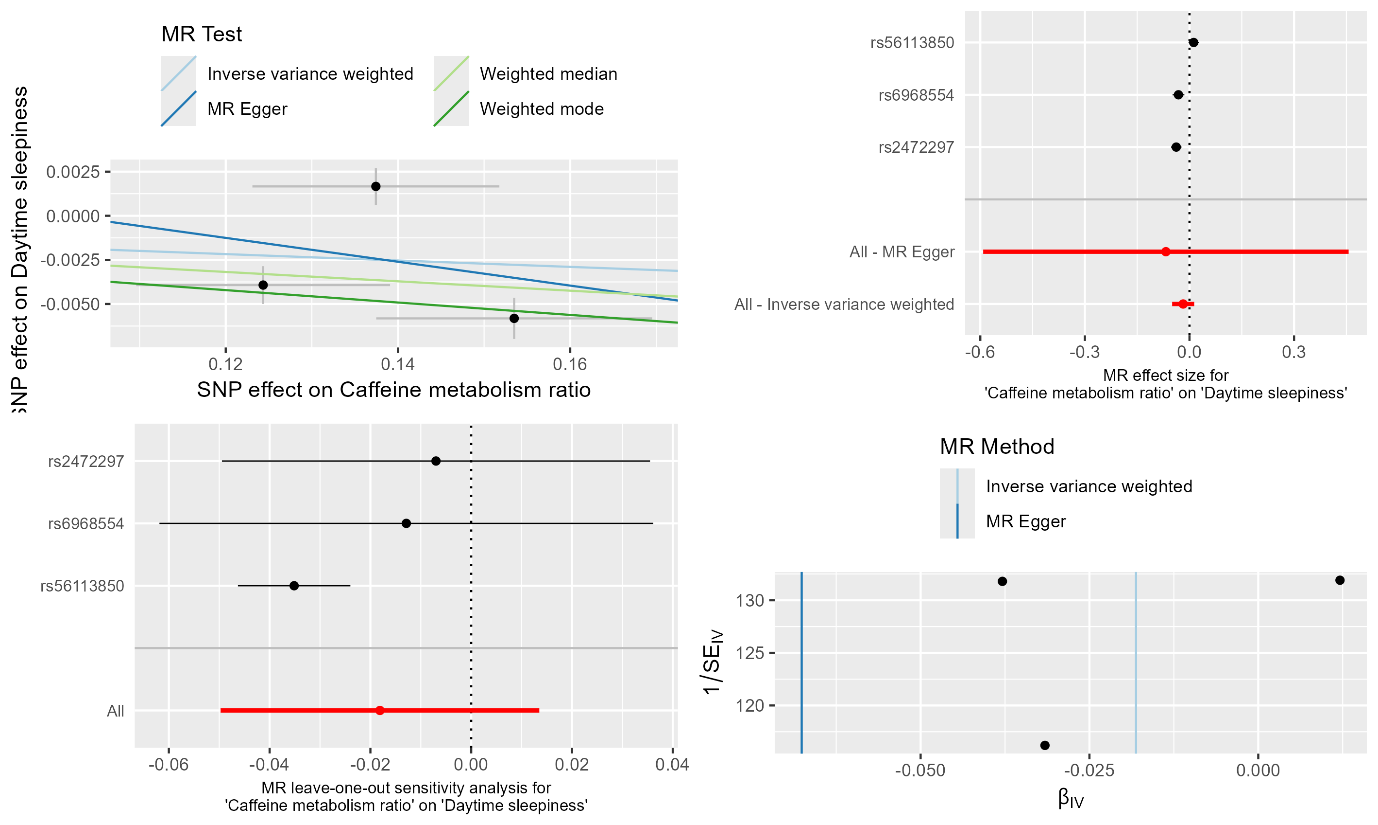

**Supplementary Figure 5c.** MR results and sensitivity analyses of caffeine metabolism ratio on daytime sleepiness.

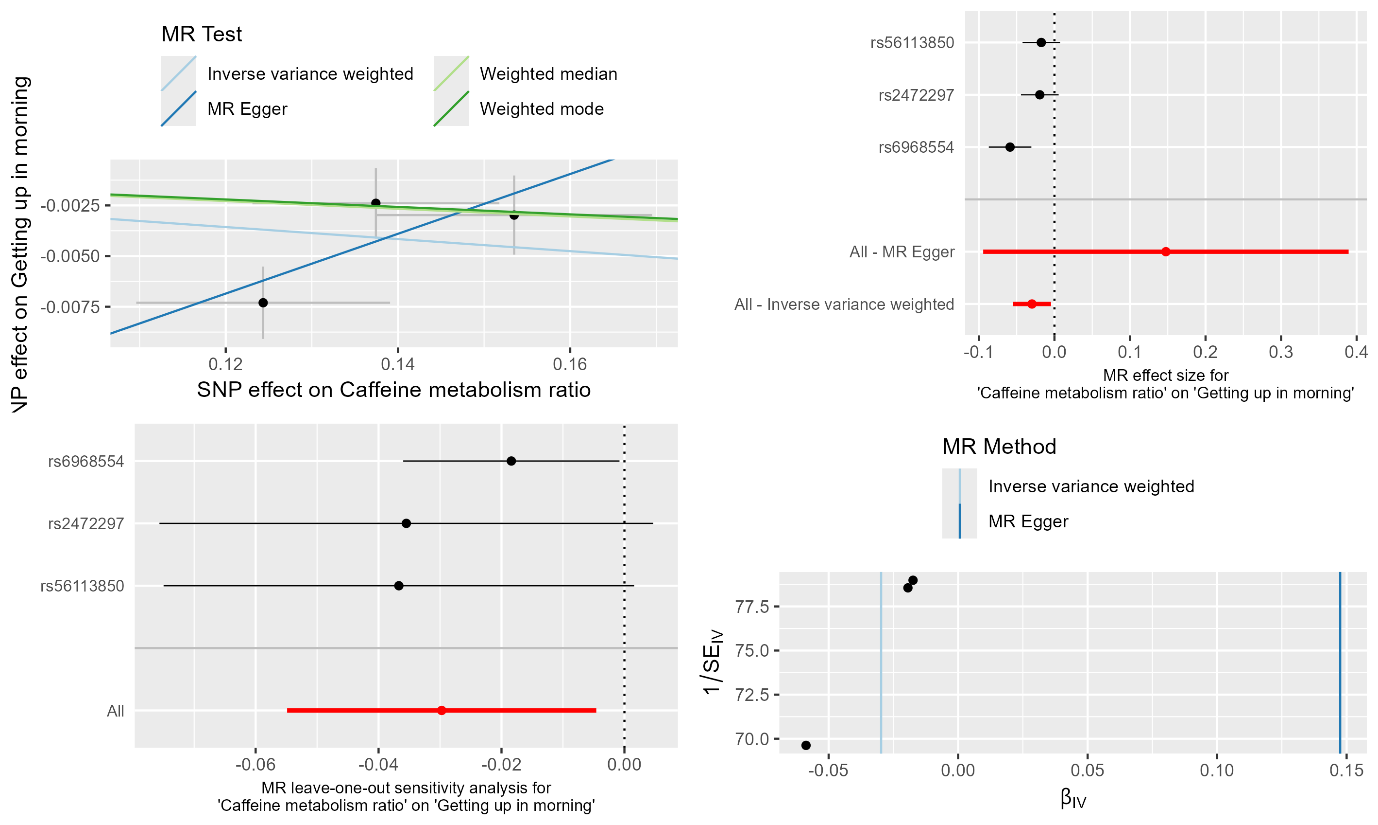

**Supplementary Figure 5d.** MR results and sensitivity analyses of caffeine metabolism ratio on getting up in morning.

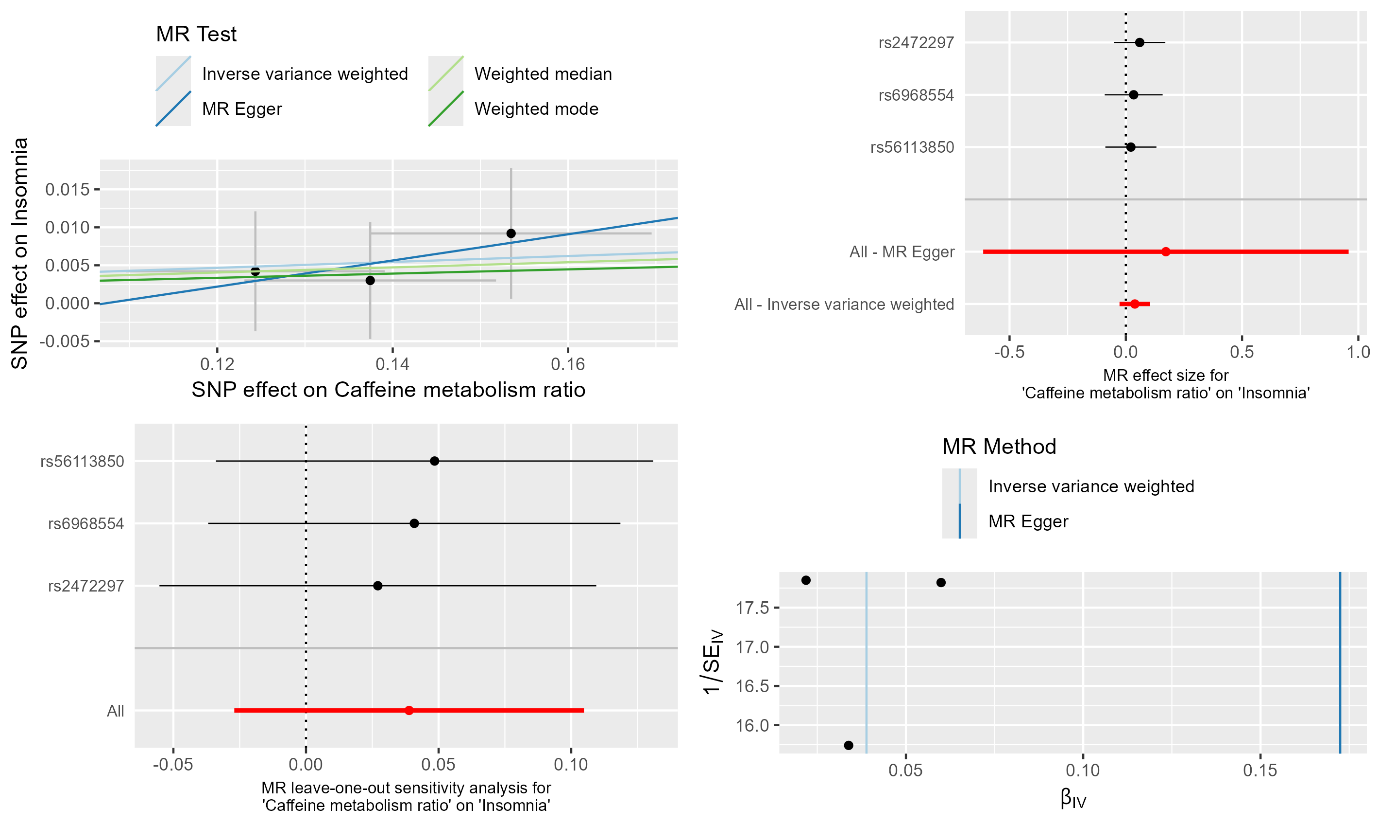

**Supplementary Figure 5e.** MR results and sensitivity analyses of caffeine metabolism ratio on insomnia.

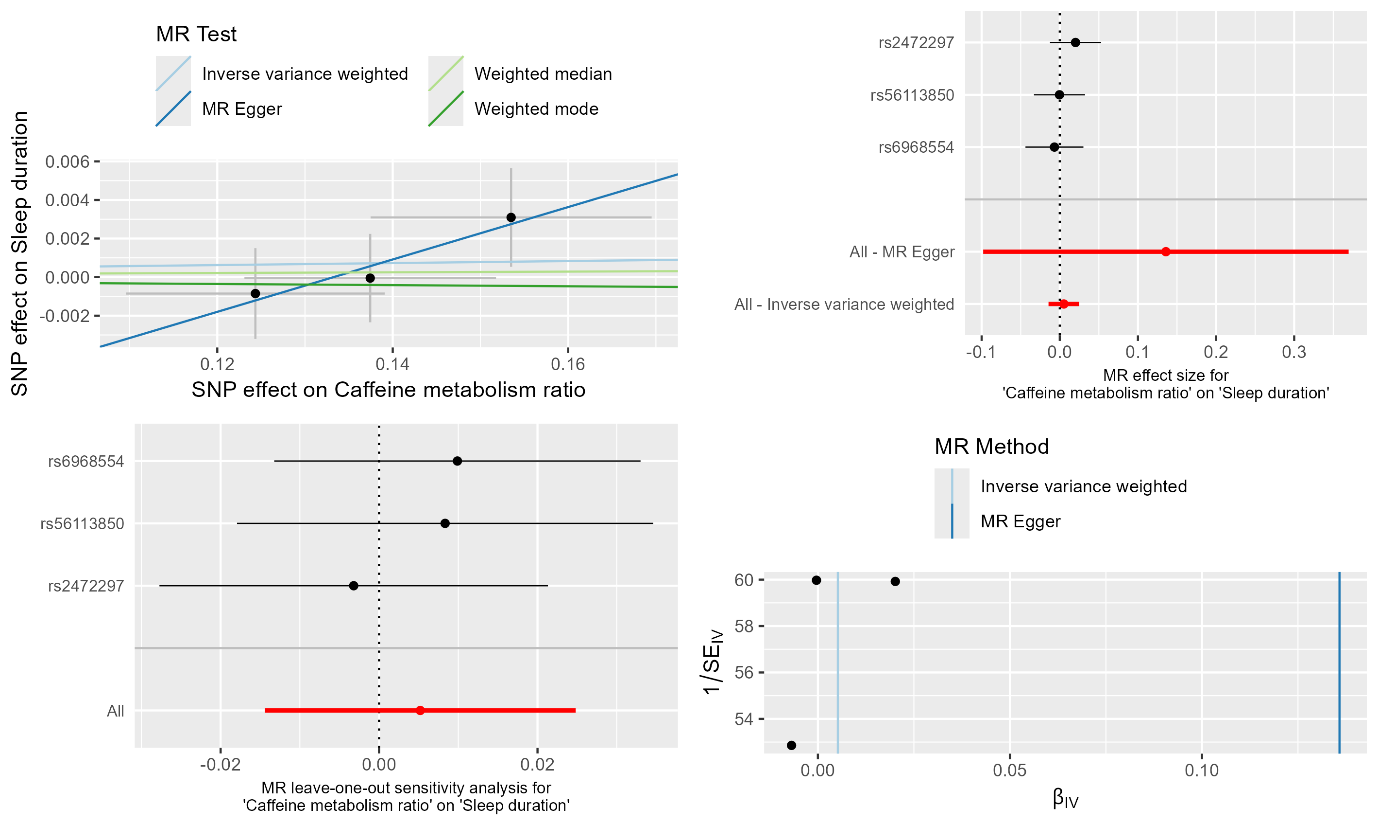

**Supplementary Figure 5f.** MR results and sensitivity analyses of caffeine metabolism ratio on sleep duration.

Top left: MR results. Black dots represent individual SNP effects. Vertical and horizontal black lines correspond to standard deviations of effects. Top right: Single SNP MR. Black dots represent individual SNP effects arranged in decreasing effect sizes. Horizontal black lines illustrate 95% confidence intervals for SNP effects. Red dots represent overall effect. Horizontal red lines illustrate 95% confidence intervals for overall effect. Bottom left: Leave-one-out MR results. Black dots represent overall effects when corresponding SNP is excluded from analysis. Horizontal black lines illustrate 95% confidence intervals for each leave-one-out MR. Red dots represent total effect when including all SNPs. Horizontal red lines illustrate 95% confidence intervals for overall effect of all SNPs. Bottom right: Funnel plot. Black dots represent individual SNP effects.

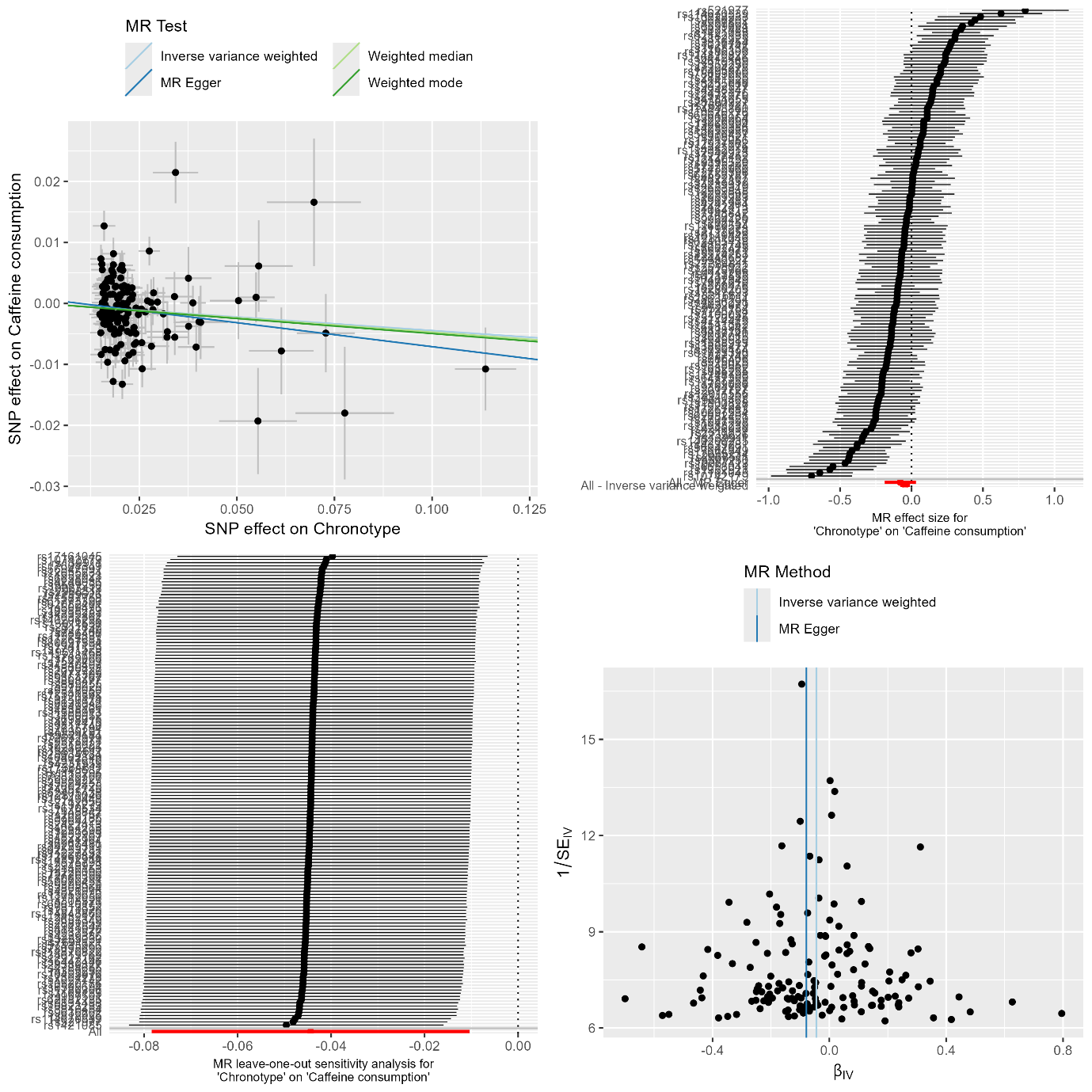

**Supplementary Figure 6a.** MR results and sensitivity analyses of chronotype on caffeine consumption.

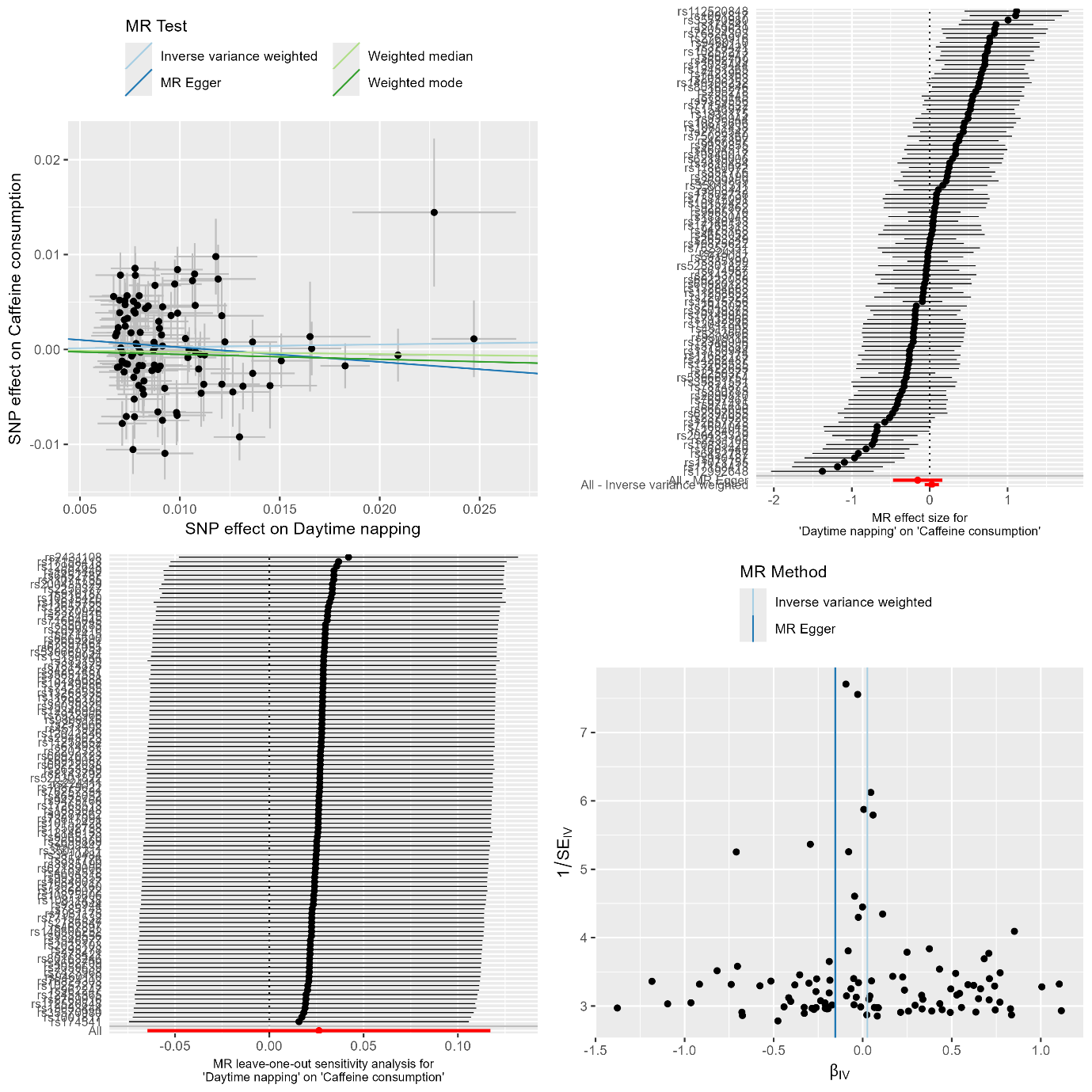

**Supplementary Figure 6b.** MR results and sensitivity analyses of daytime napping on caffeine consumption.

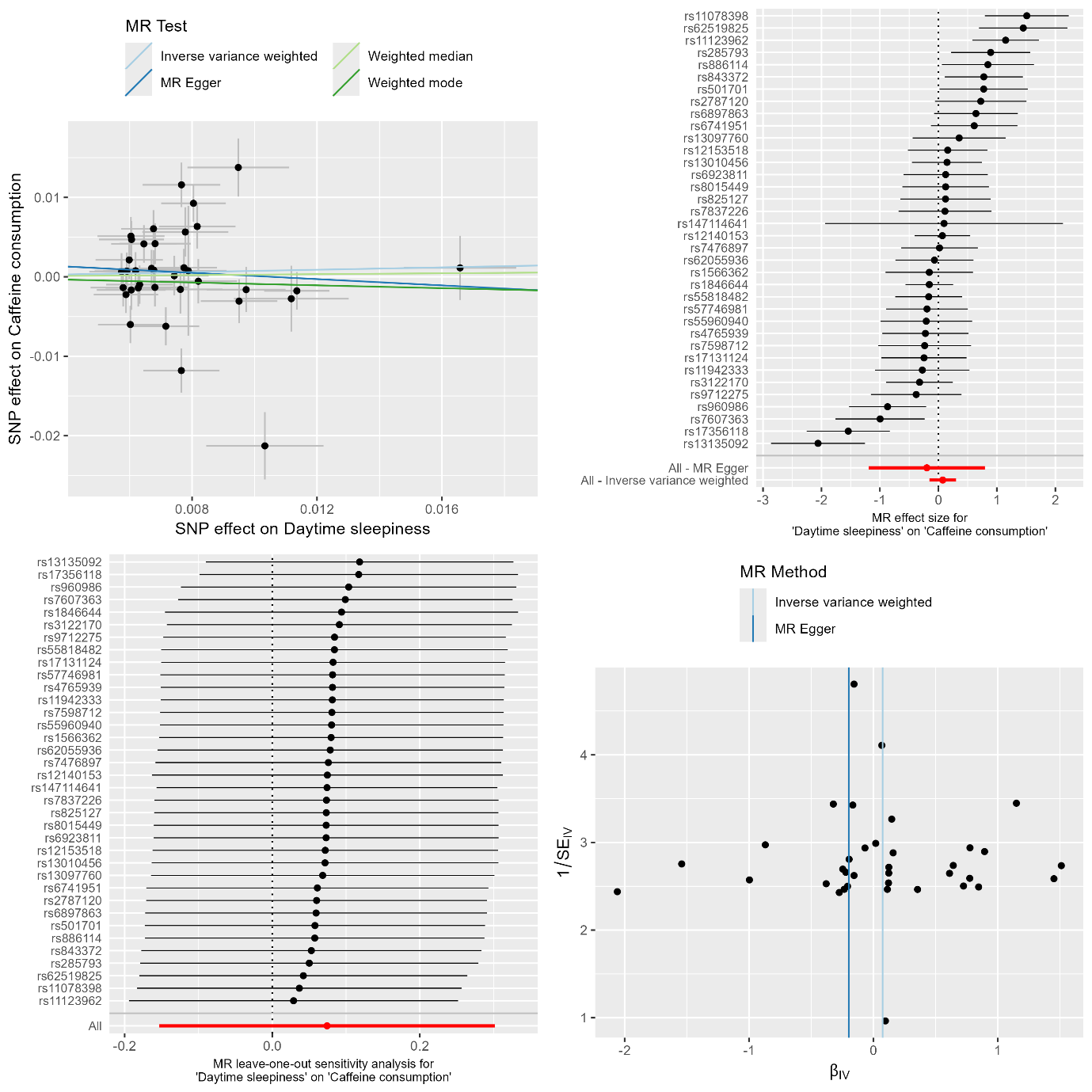

**Supplementary Figure 6c.** MR results and sensitivity analyses of daytime sleepiness on caffeine consumption.

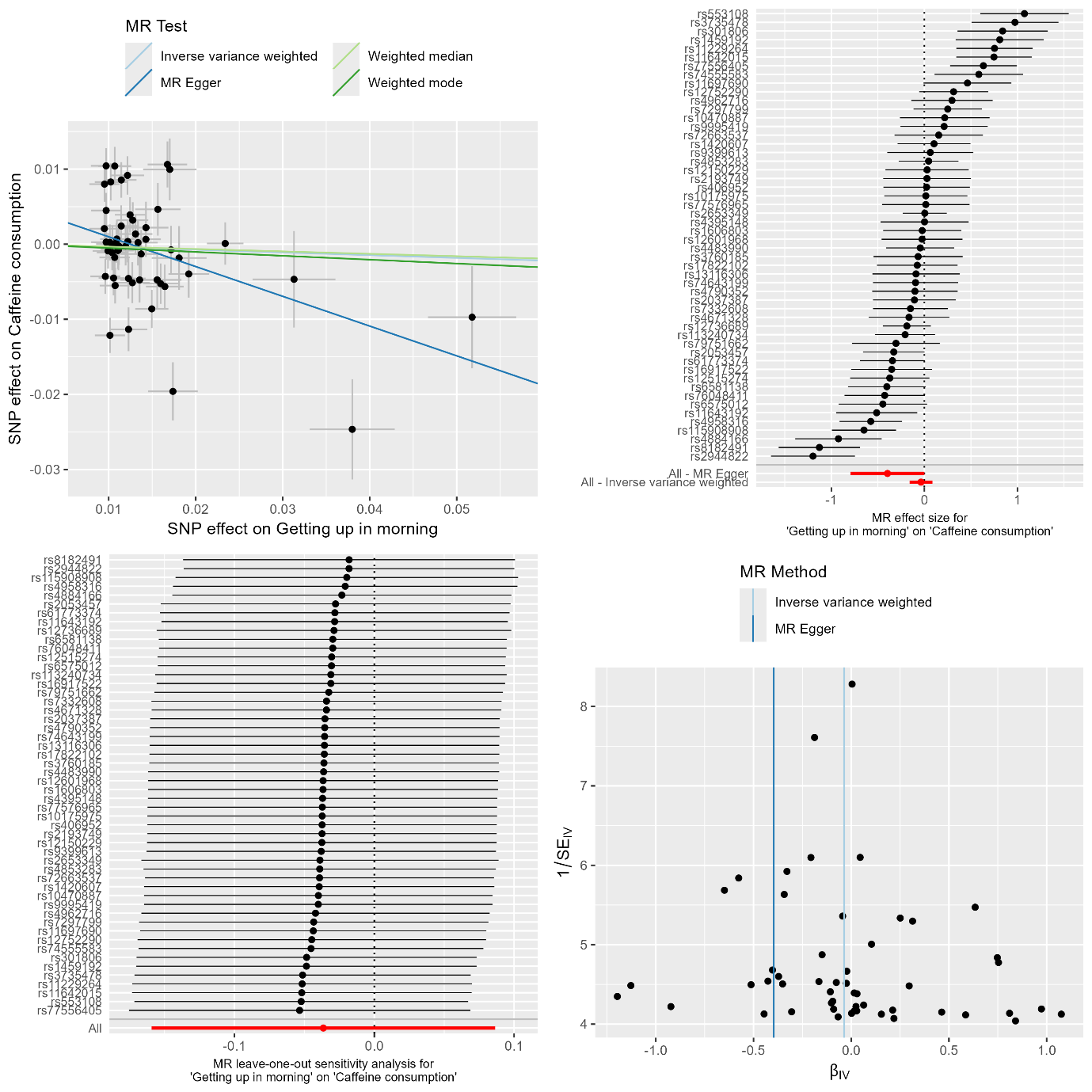

**Supplementary Figure 6d.** MR results and sensitivity analyses of getting up in morning on caffeine consumption.

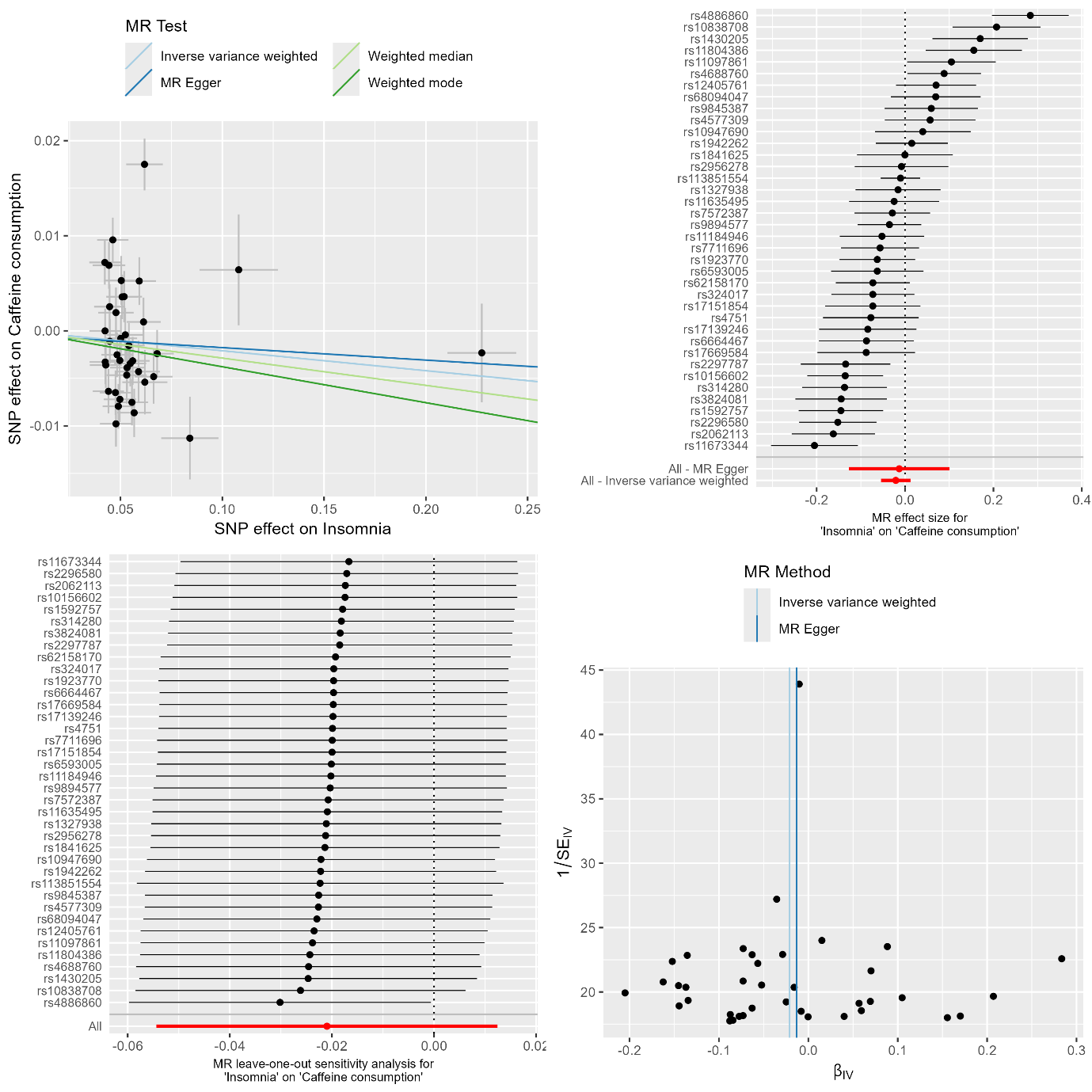

**Supplementary Figure 6e.** MR results and sensitivity analyses of insomnia on caffeine consumption.

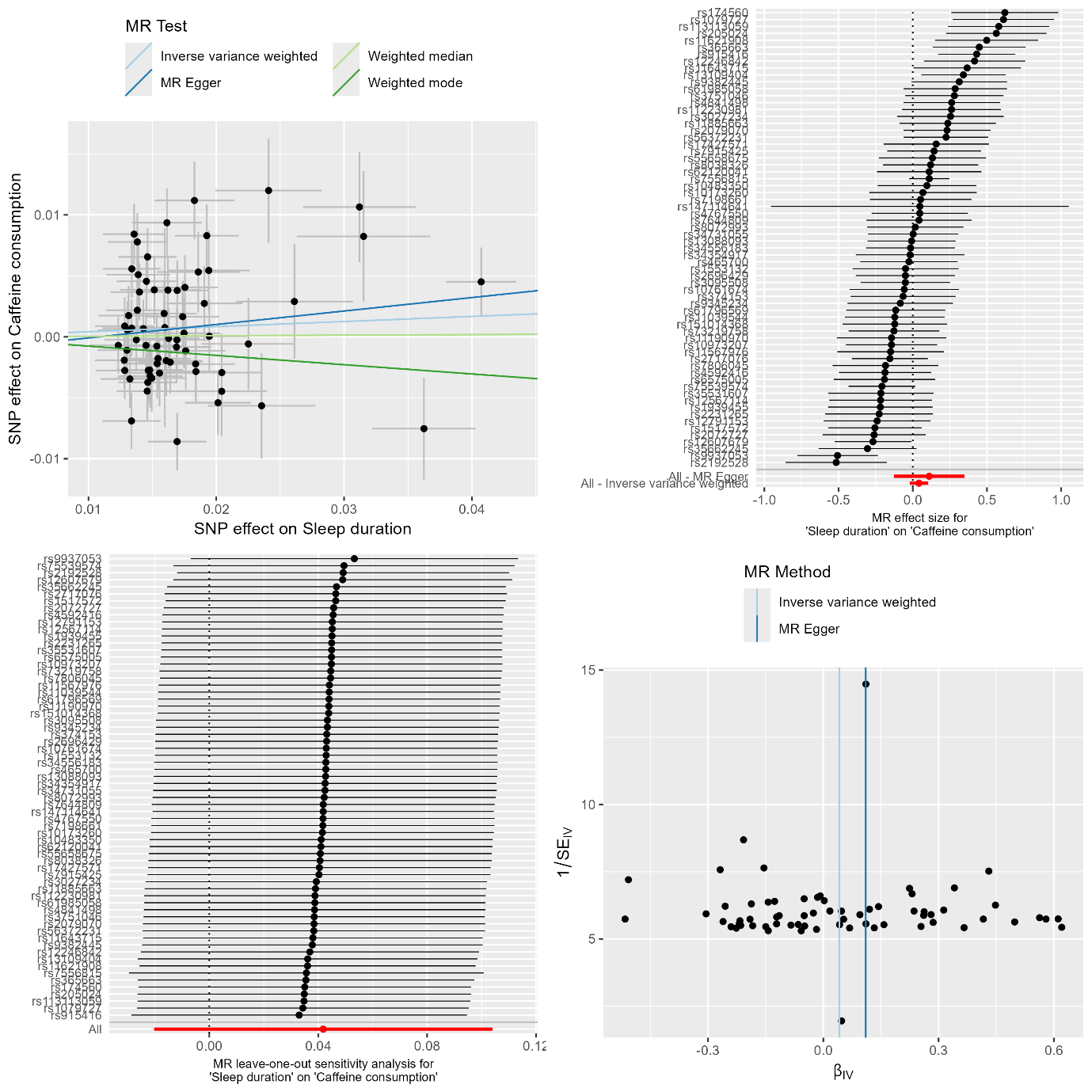

**Supplementary Figure 6f.** MR results and sensitivity analyses of sleep duration on caffeine consumption.

Top left: MR results. Black dots represent individual SNP effects. Vertical and horizontal black lines correspond to standard deviations of effects. Top right: Single SNP MR. Black dots represent individual SNP effects arranged in decreasing effect sizes. Horizontal black lines illustrate 95% confidence intervals for SNP effects. Red dots represent overall effect. Horizontal red lines illustrate 95% confidence intervals for overall effect. Bottom left: Leave-one-out MR results. Black dots represent overall effects when corresponding SNP is excluded from analysis. Horizontal black lines illustrate 95% confidence intervals for each leave-one-out MR. Red dots represent total effect when including all SNPs. Horizontal red lines illustrate 95% confidence intervals for overall effect of all SNPs. Bottom right: Funnel plot. Black dots represent individual SNP effects.

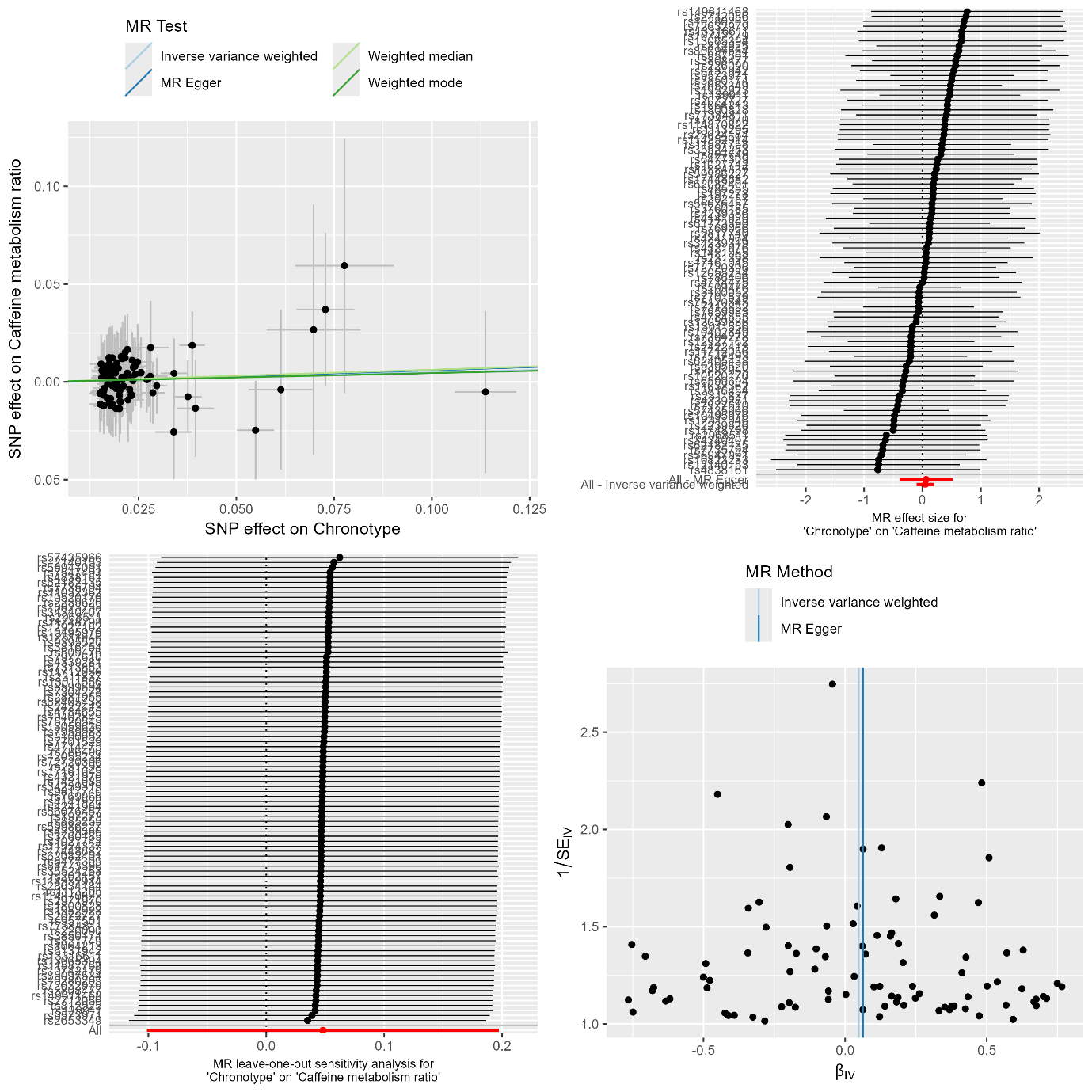

**Supplementary Figure 7a.** MR results and sensitivity analyses of chronotype on caffeine metabolism ratio.

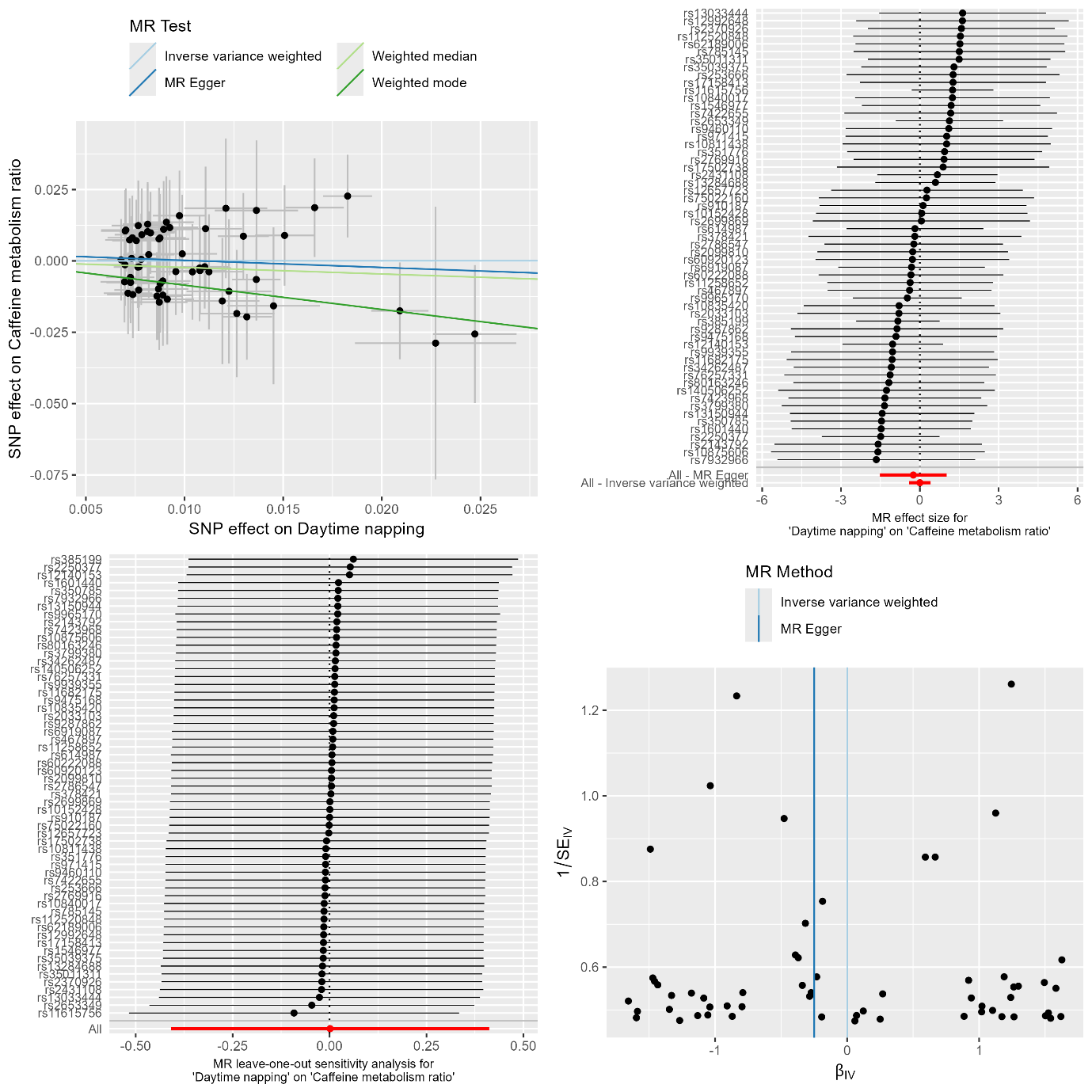

**Supplementary Figure 7b.** MR results and sensitivity analyses of daytime napping on caffeine metabolism ratio.

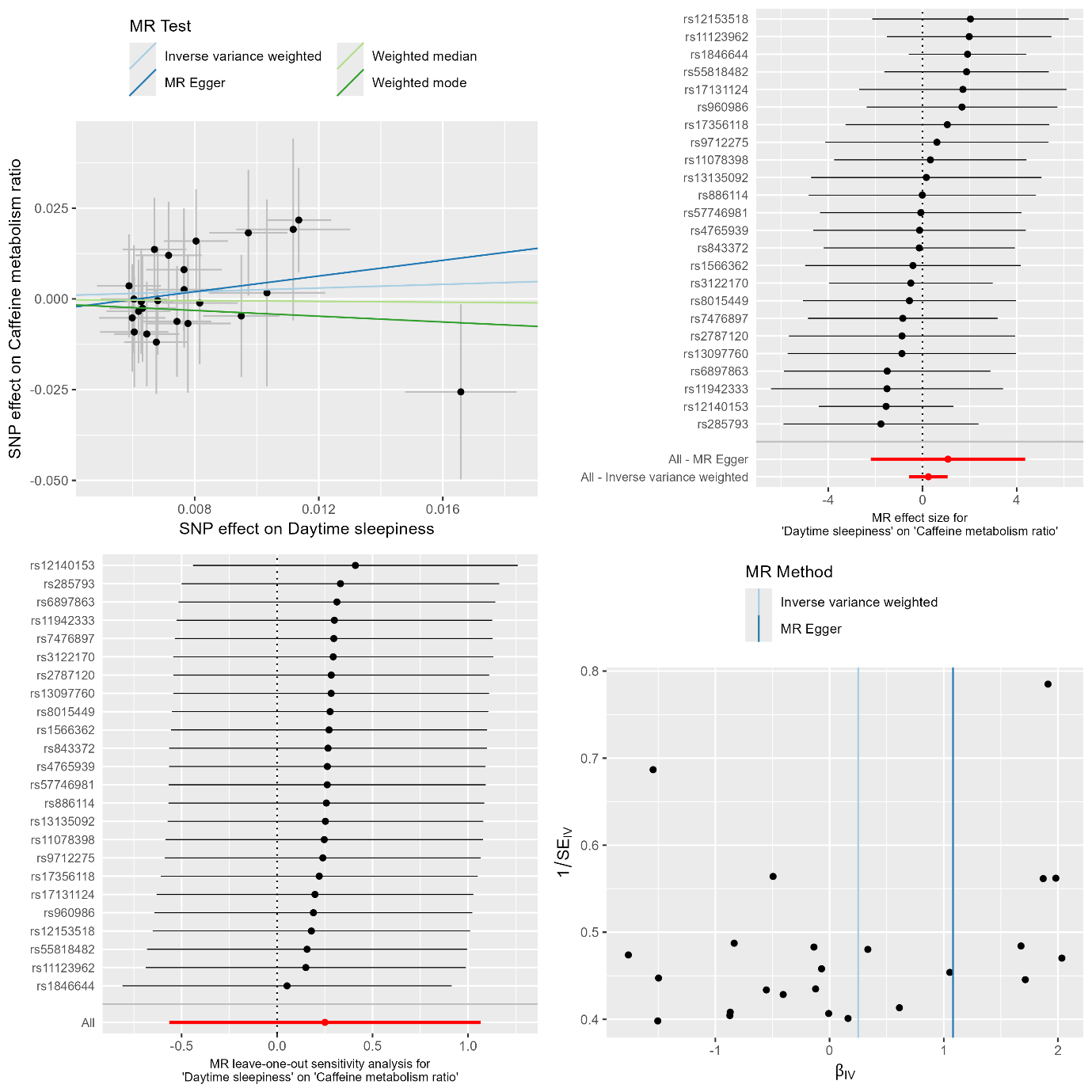

**Supplementary Figure 7c.** MR results and sensitivity analyses of daytime sleepiness on caffeine metabolism ratio.

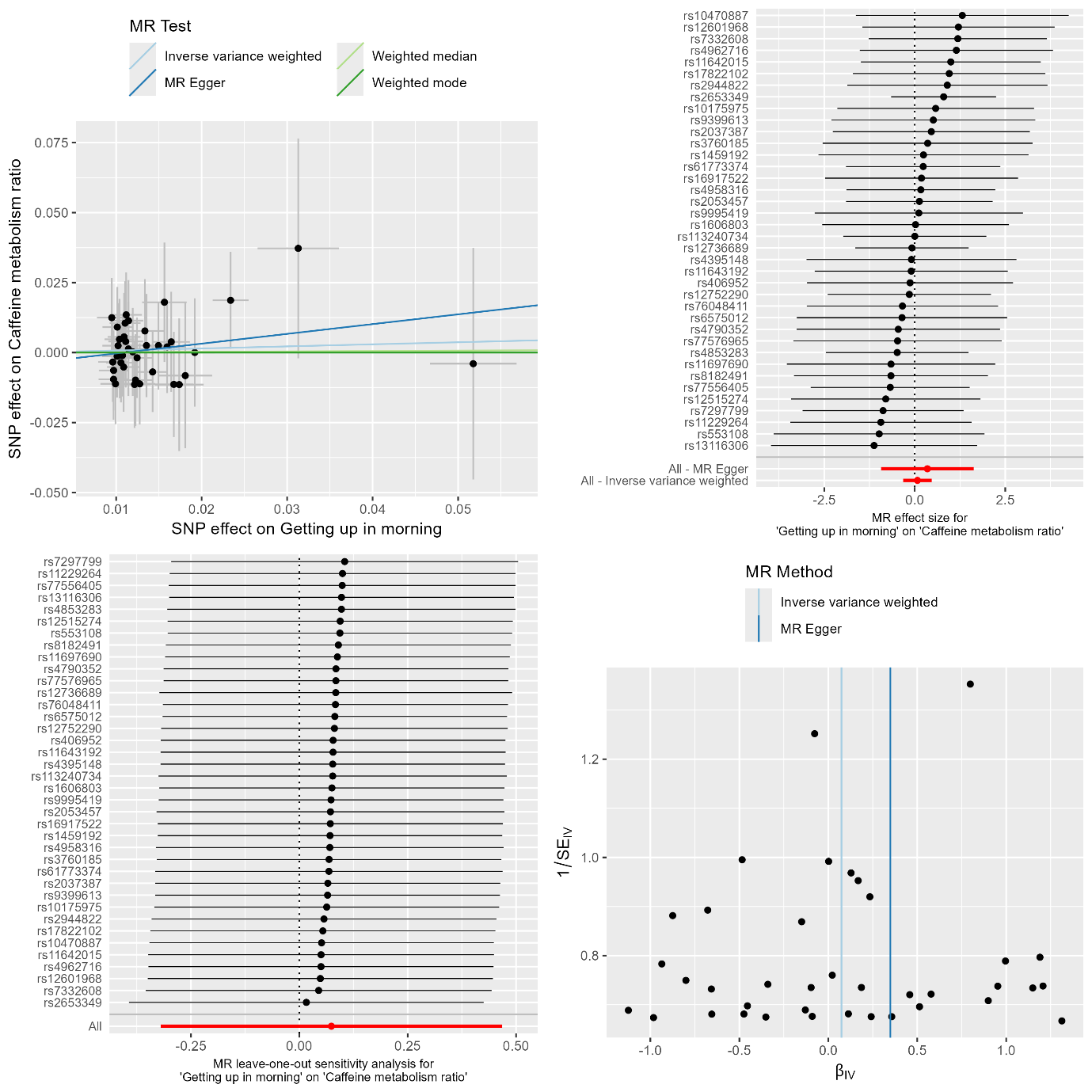

**Supplementary Figure 7d.** MR results and sensitivity analyses of getting up in morning on caffeine metabolism ratio.

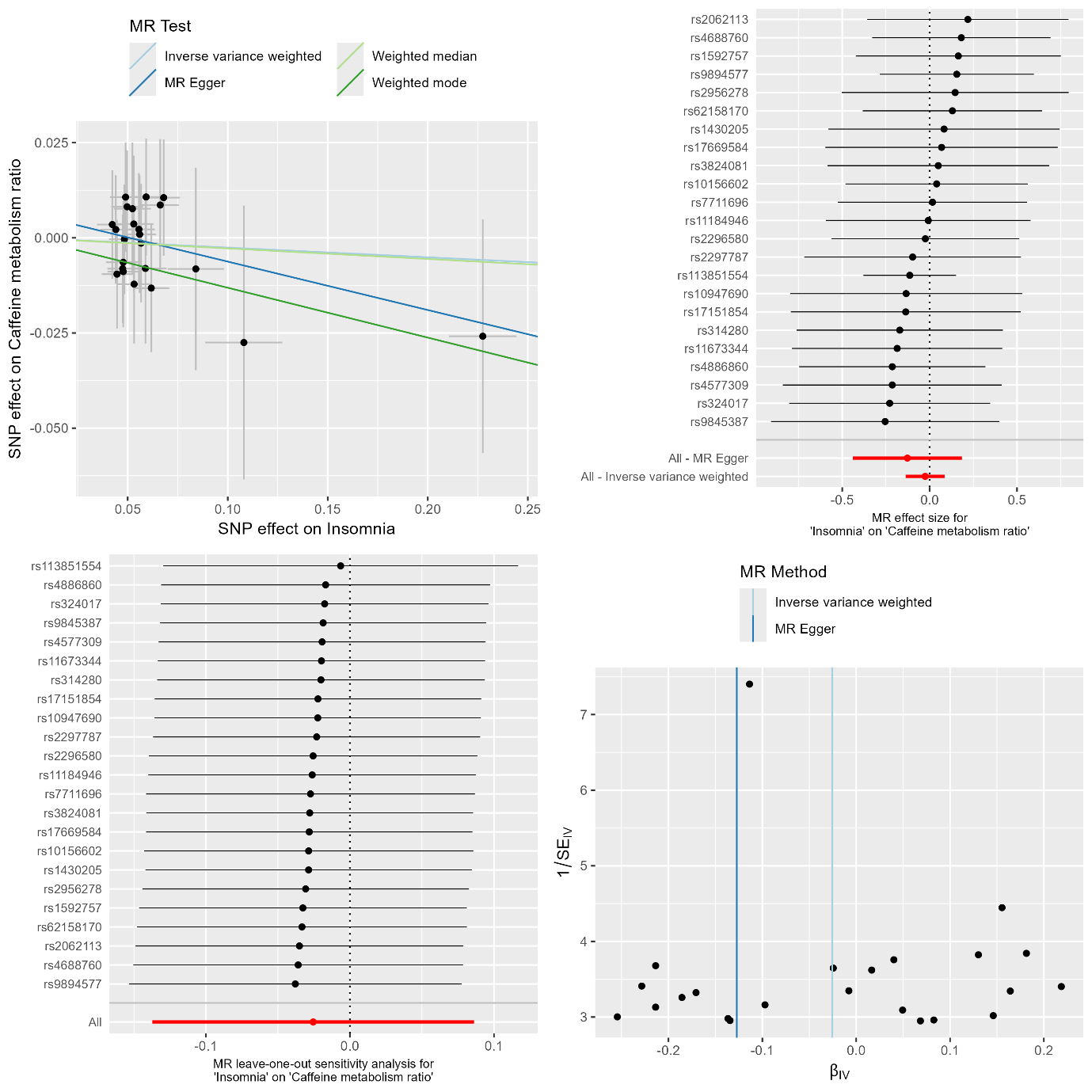

**Supplementary Figure 7e.** MR results and sensitivity analyses of insomnia on caffeine metabolism ratio.

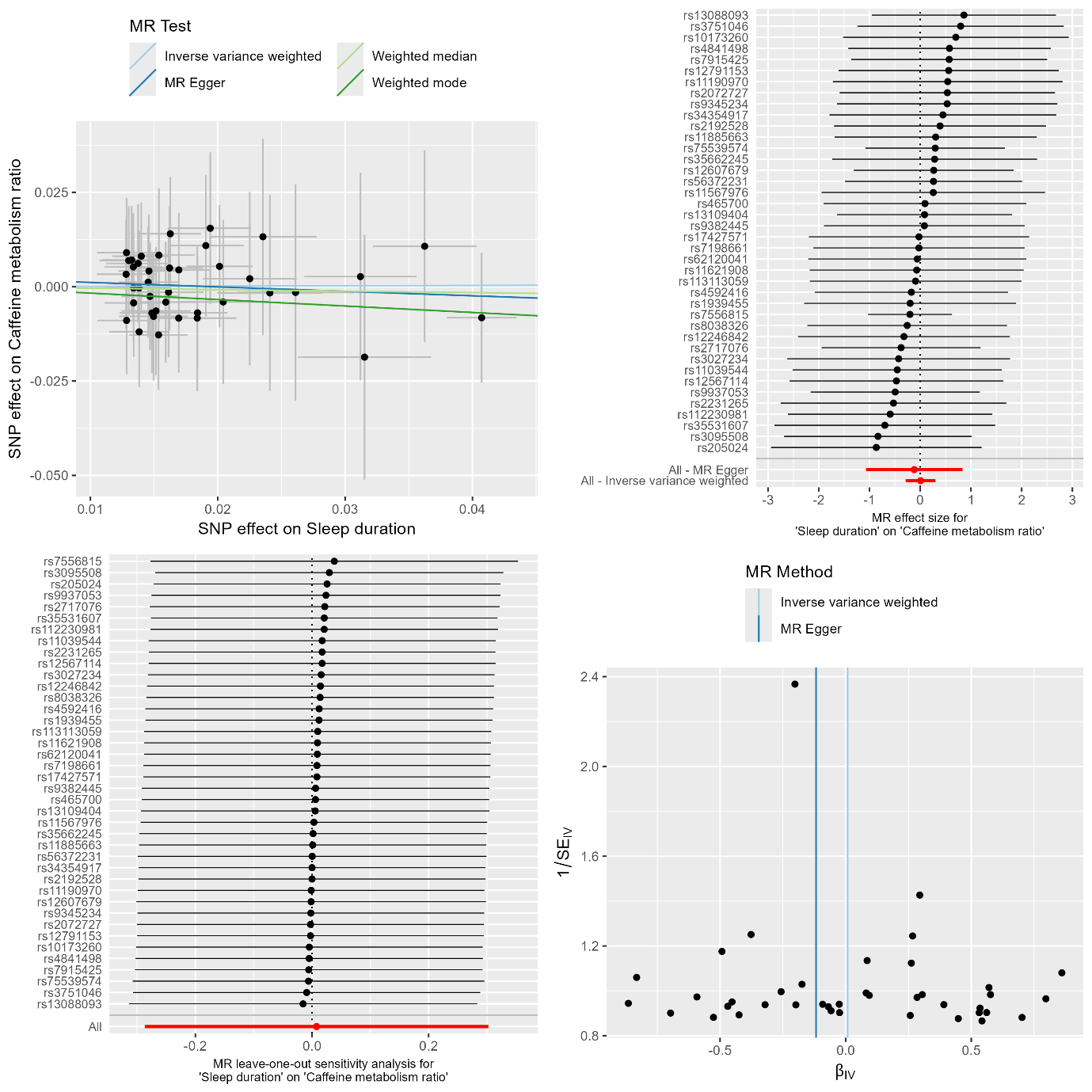

**Supplementary Figure 7f.** MR results and sensitivity analyses of sleep duration on caffeine metabolism ratio.

Top left: MR results. Black dots represent individual SNP effects. Vertical and horizontal black lines correspond to standard deviations of effects. Top right: Single SNP MR. Black dots represent individual SNP effects arranged in decreasing effect sizes. Horizontal black lines illustrate 95% confidence intervals for SNP effects. Red dots represent overall effect. Horizontal red lines illustrate 95% confidence intervals for overall effect. Bottom left: Leave-one-out MR results. Black dots represent overall effects when corresponding SNP is excluded from analysis. Horizontal black lines illustrate 95% confidence intervals for each leave-one-out MR. Red dots represent total effect when including all SNPs. Horizontal red lines illustrate 95% confidence intervals for overall effect of all SNPs. Bottom right: Funnel plot. Black dots represent individual SNP effects.

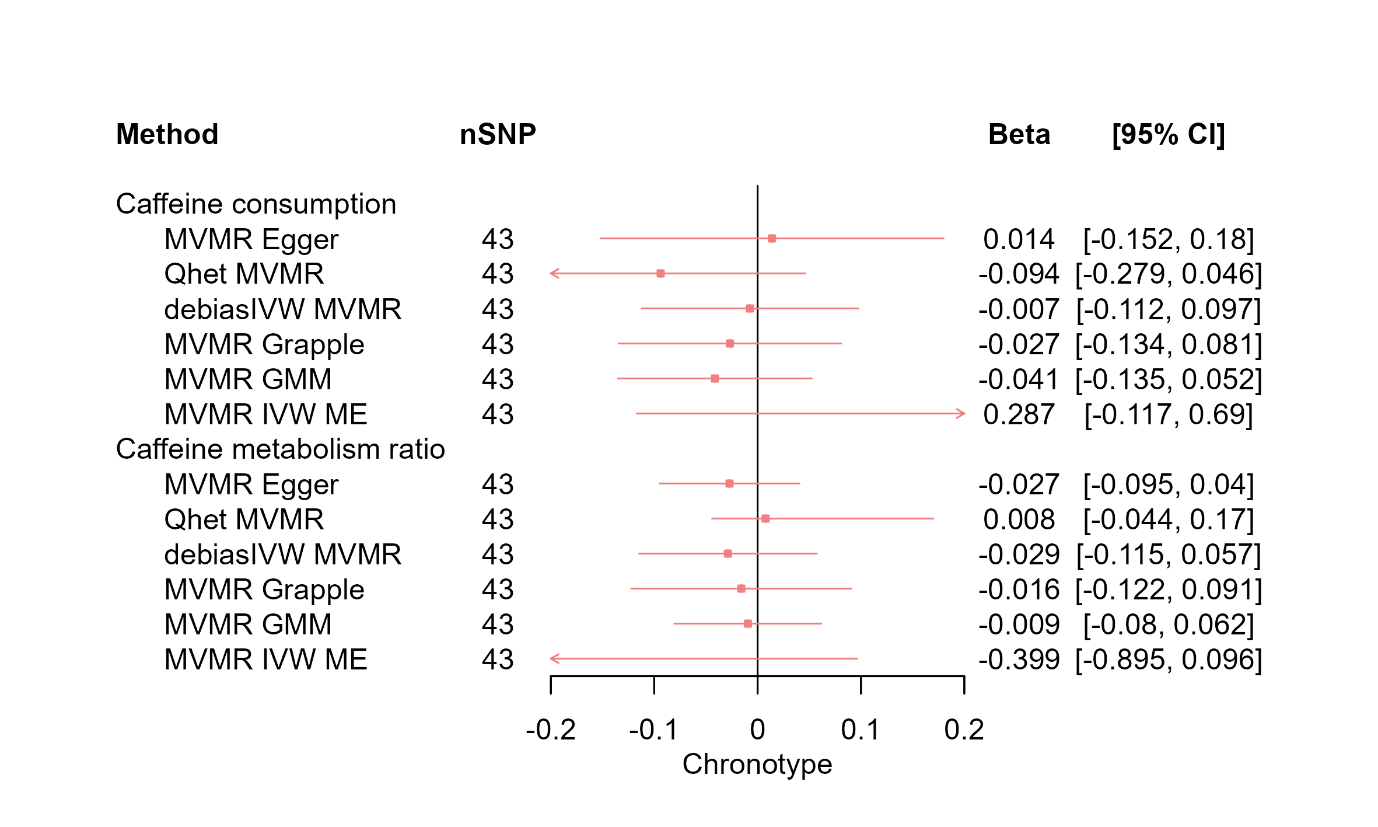

**Supplementary Figure 8a.** MR results of effects of caffeine consumption and caffeine metabolism ratio on chronotype using pleiotropy robust and weak instrument robust MVMR.

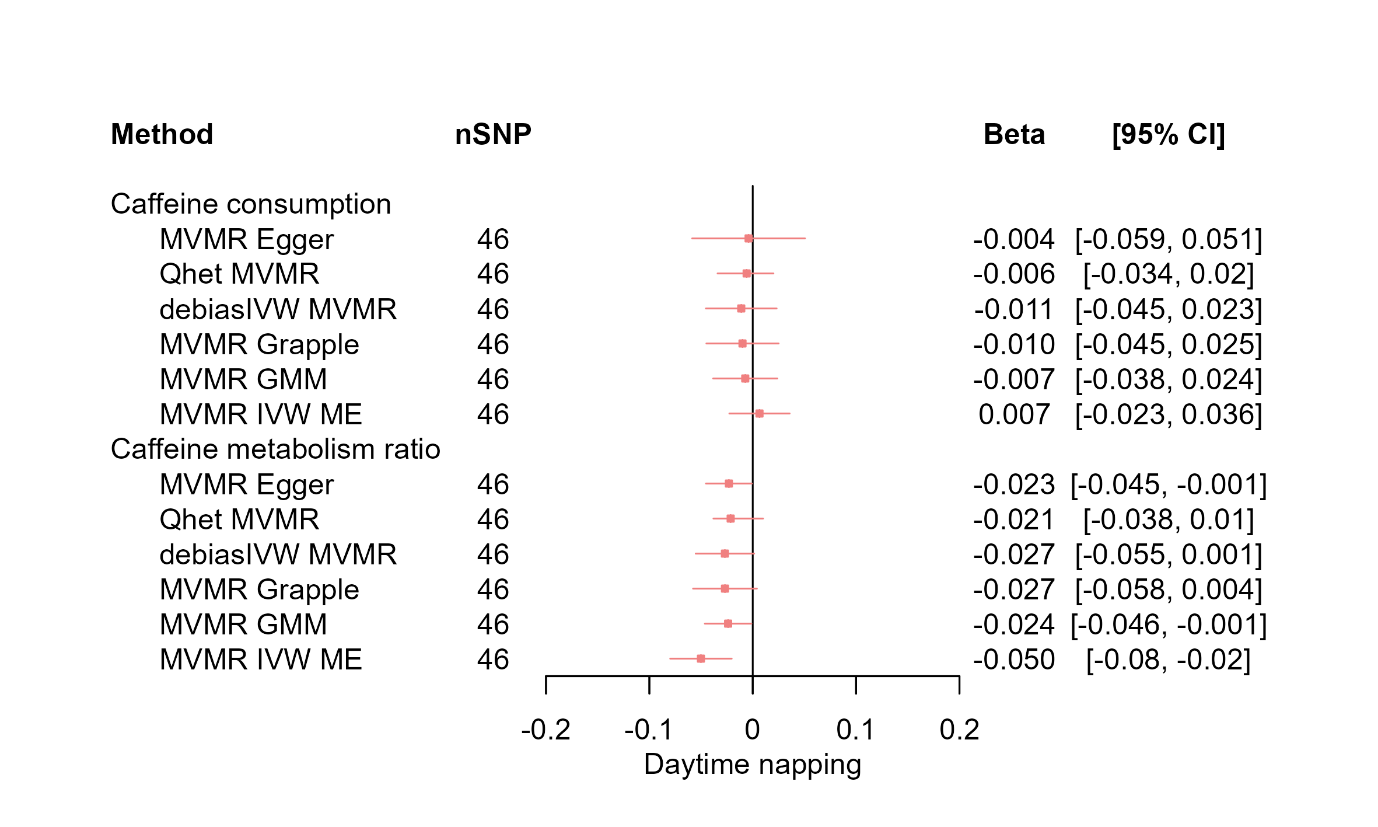

**Supplementary Figure 8b.** MR results of effects of caffeine consumption and caffeine metabolism ratio on daytime napping using pleiotropy robust and weak instrument robust MVMR.

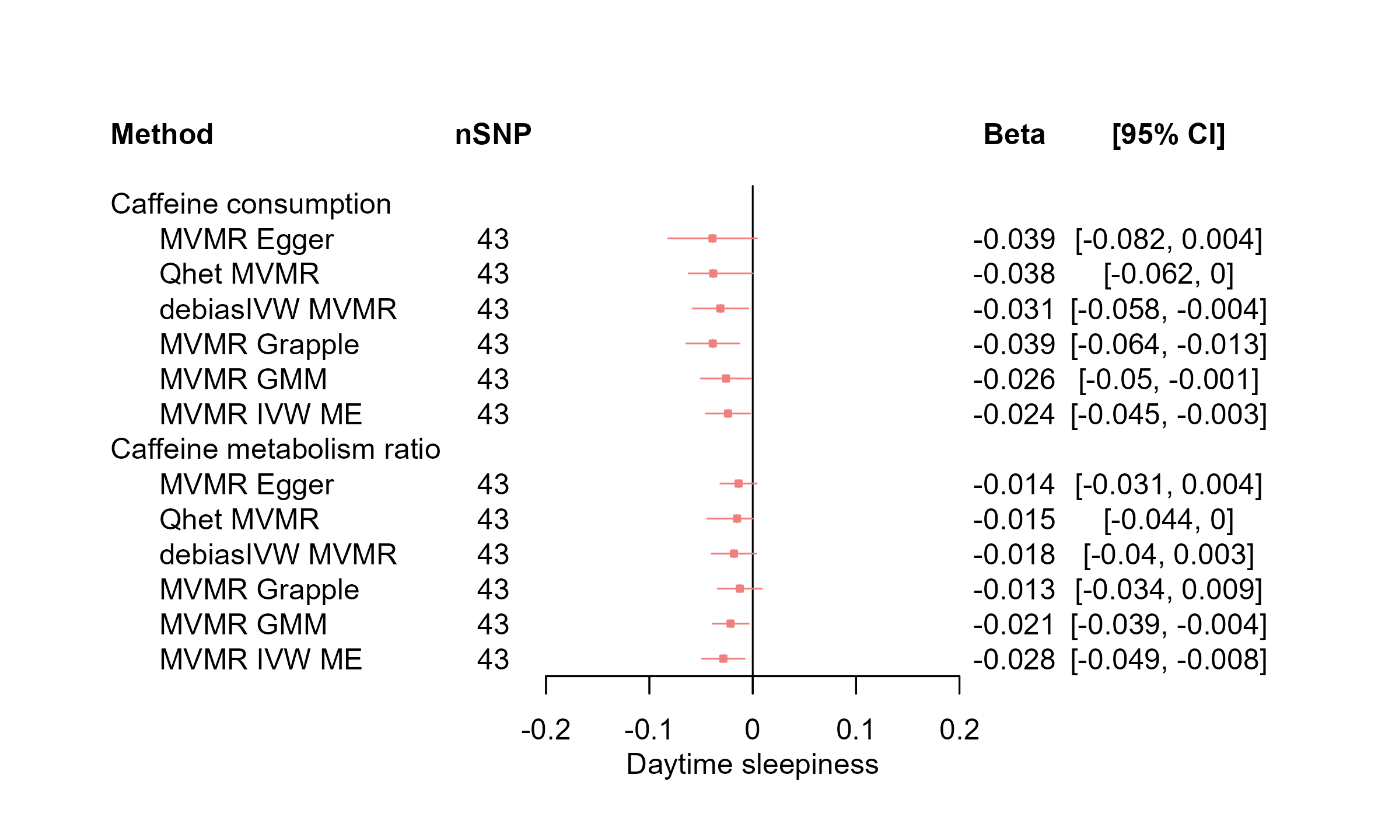

**Supplementary Figure 8c.** MR results of effects of caffeine consumption and caffeine metabolism ratio on daytime sleepiness using pleiotropy robust and weak instrument robust MVMR.

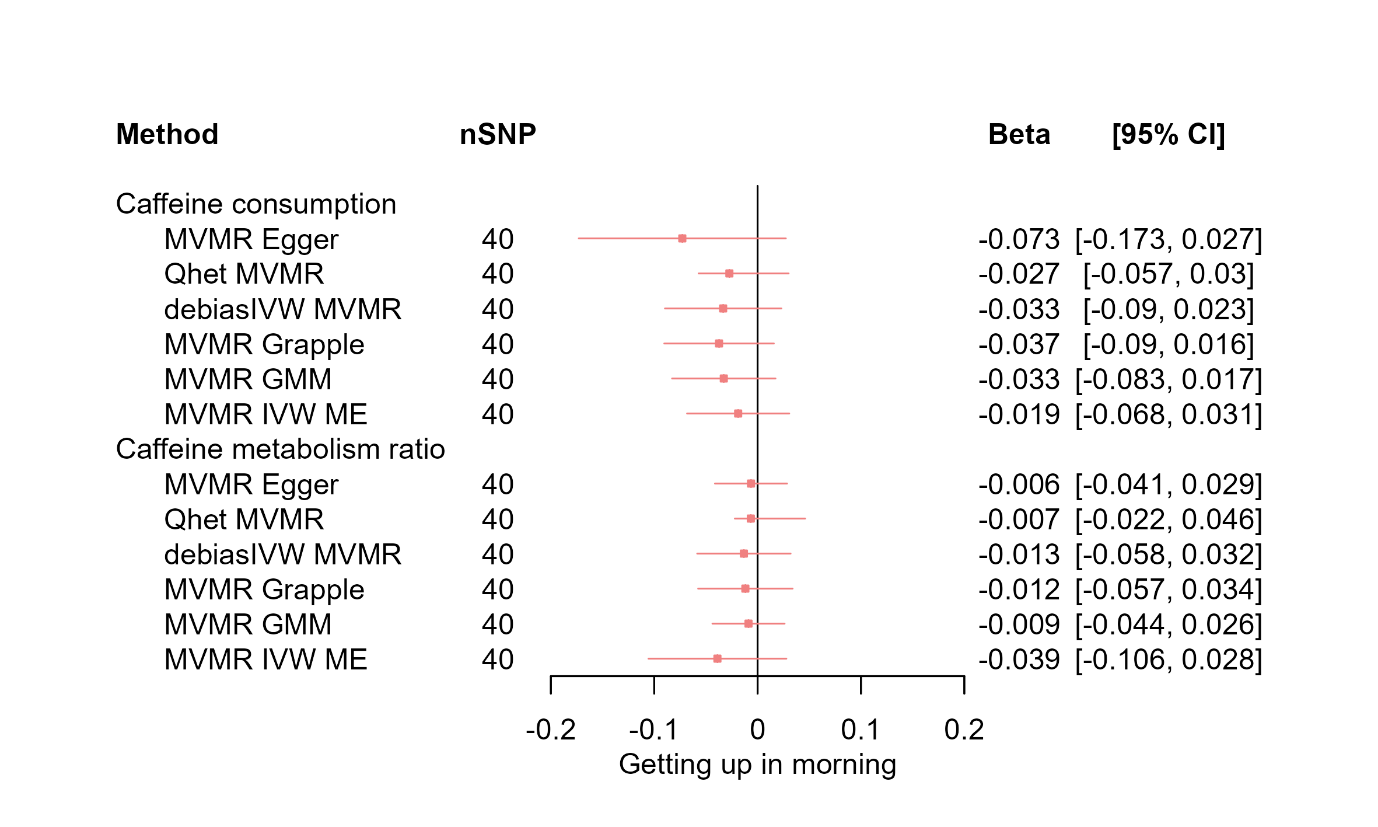

**Supplementary Figure 8d.** MR results of effects of caffeine consumption and caffeine metabolism ratio on getting up in morning using pleiotropy robust and weak instrument robust MVMR.

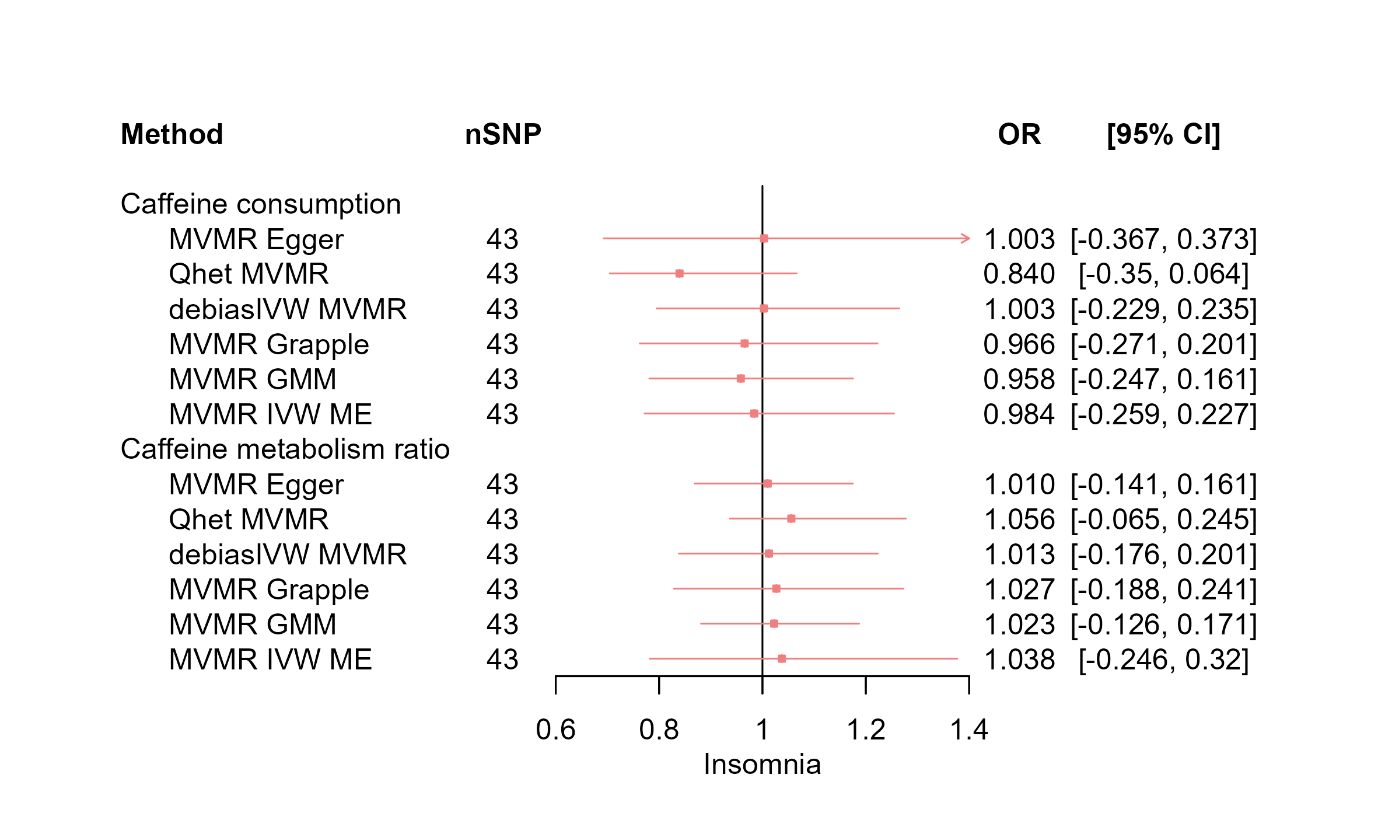

**Supplementary Figure 8e.** MR results of effects of caffeine consumption and caffeine metabolism ratio on insomnia using pleiotropy robust and weak instrument robust MVMR.

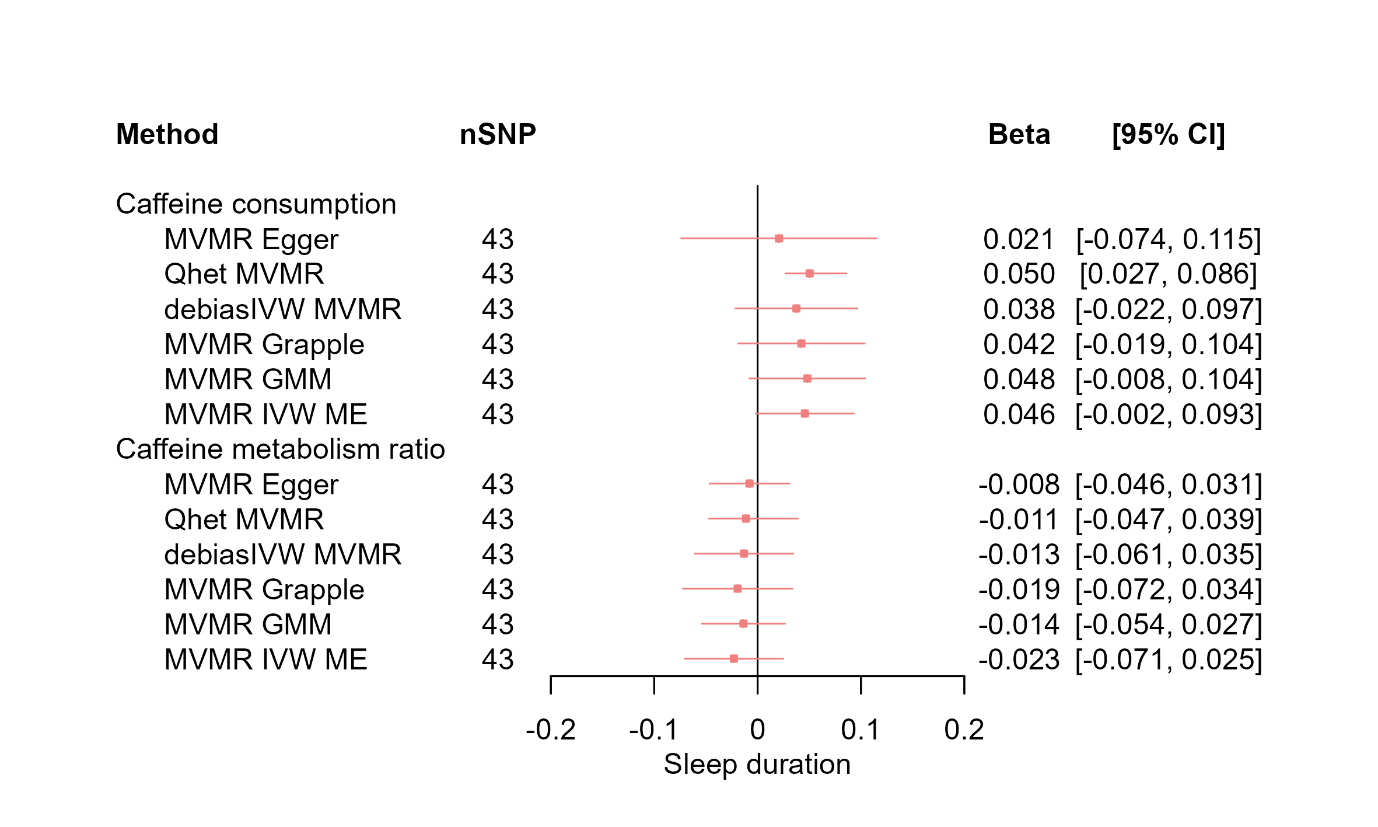

**Supplementary Figure 8f.** MR results of effects of caffeine consumption and caffeine metabolism ratio on sleep duration using pleiotropy robust and weak instrument robust MVMR.

Red squares represent the estimated effect sizes (betas or odds ratio (OR)) for each individual estimation method. The red horizontal lines represent the 95% confidence intervals (95% CI) for the estimated effects. The black vertical line represents the point of no effect. “nSNP” gives the number of single nucleotide polymorphisms (SNPs) used in each estimation method.

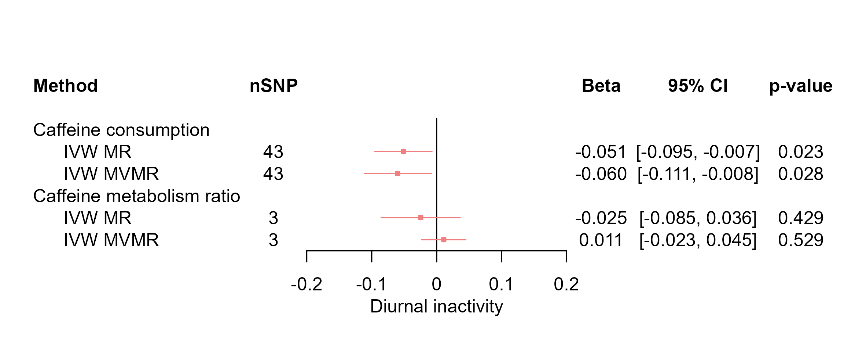

**Supplementary Figure 9a.** MR results of effects of caffeine consumption and caffeine metabolism ratio on diurnal inactivity.

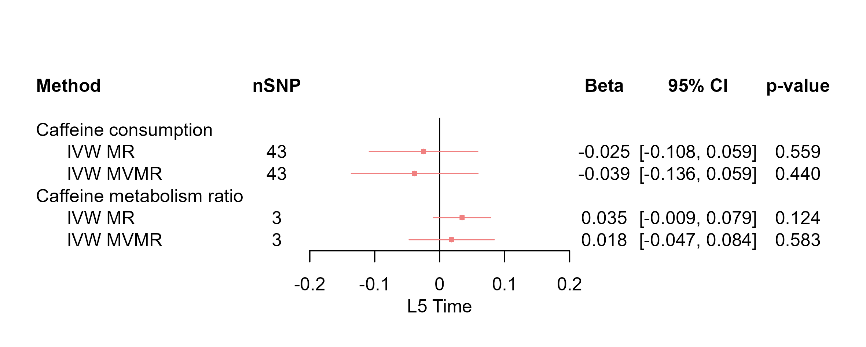

**Supplementary Figure 9b.** MR results of effects of caffeine consumption and caffeine metabolism ratio on L5 time.

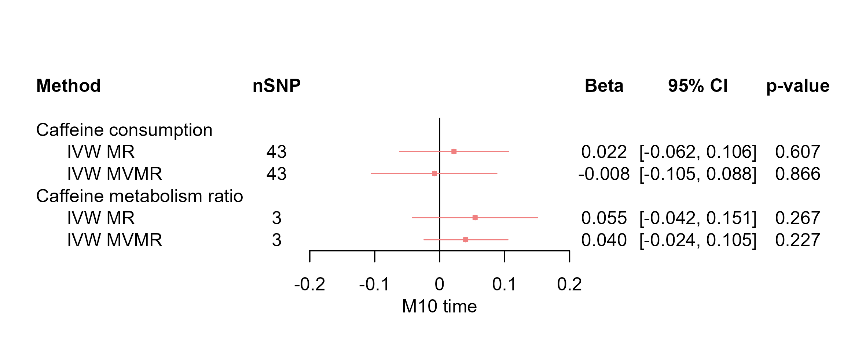

**Supplementary Figure 9c.** MR results of effects of caffeine consumption and caffeine metabolism ratio on M10 time.

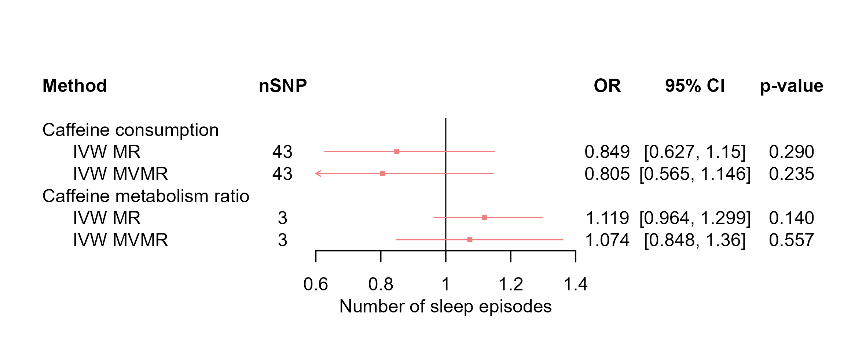

**Supplementary Figure 9d.** MR results of effects of caffeine consumption and caffeine metabolism ratio on number of nocturnal sleep episodes.

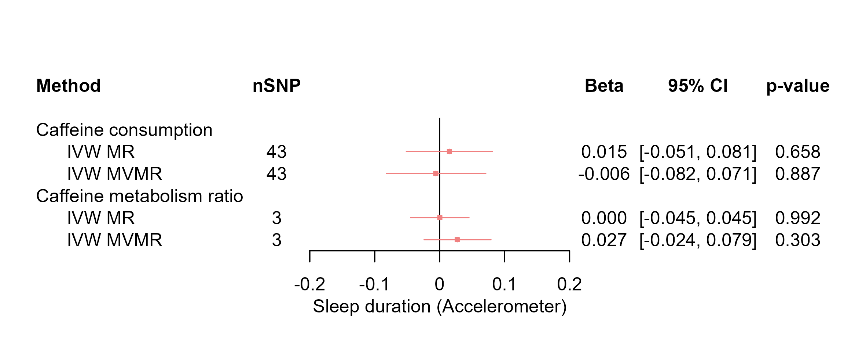

**Supplementary Figure 9e.** MR results of effects of caffeine consumption and caffeine metabolism ratio on accelerometer-derived sleep duration.

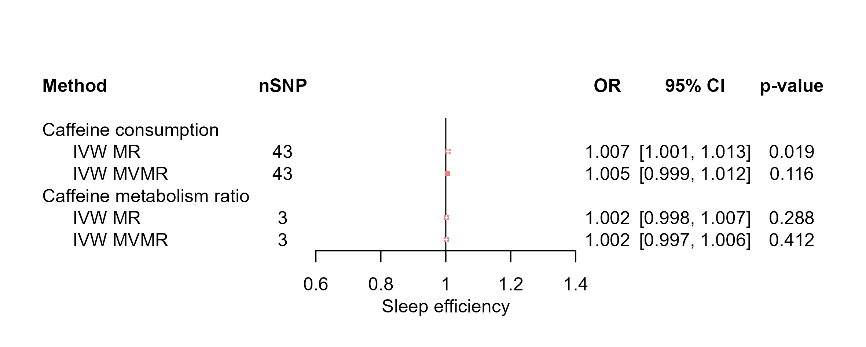

**Supplementary Figure 9f.** MR results of effects of caffeine consumption and caffeine metabolism ratio on sleep efficiency.

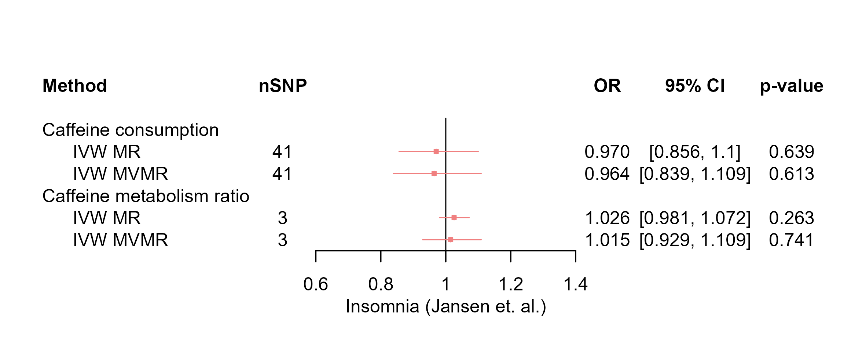

**Supplementary Figure 9g.** MR results of effects of caffeine consumption and caffeine metabolism ratio on insomnia (Jansen et al. GWAS (21)).

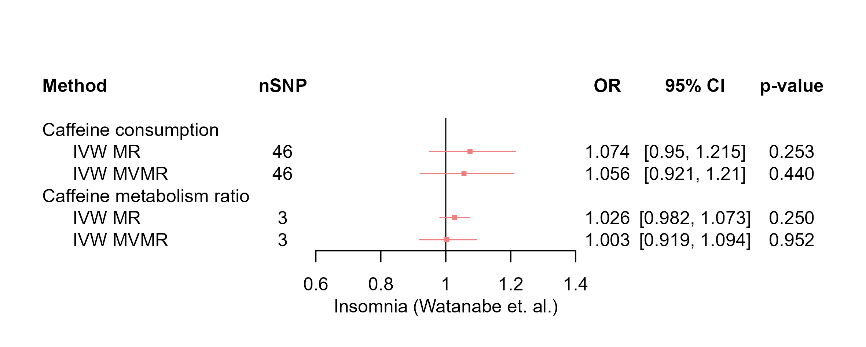

**Supplementary Figure 9h.** MR results of effects of caffeine consumption and caffeine metabolism ratio on insomnia (Watanabe et al. GWAS (22)).

Red squares represent the estimated effect sizes (betas or odds ratios (ORs)) for each individual estimation method. The red horizontal lines represent the 95% confidence intervals (95% CI) for the estimated effects. The black vertical line represents the point of no effect. “nSNP” gives the number of single nucleotide polymorphisms (SNPs) used in each estimation method. “IVW MR” gives the total effects estimated using inverse variance weighted (IVW) Mendelian Randomisation (MR). “IVW MVMR” gives the direct effects estimated using Multivariable Mendelian Randomisation (MVMR).

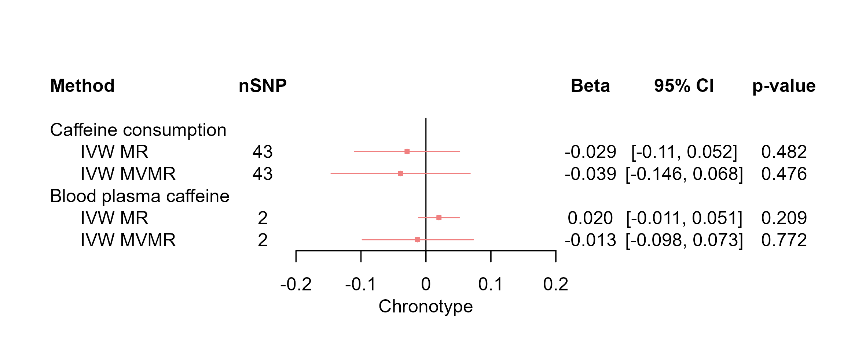

**Supplementary Figure 10a.** MR results of effects of caffeine consumption and blood plasma caffeine on chronotype.

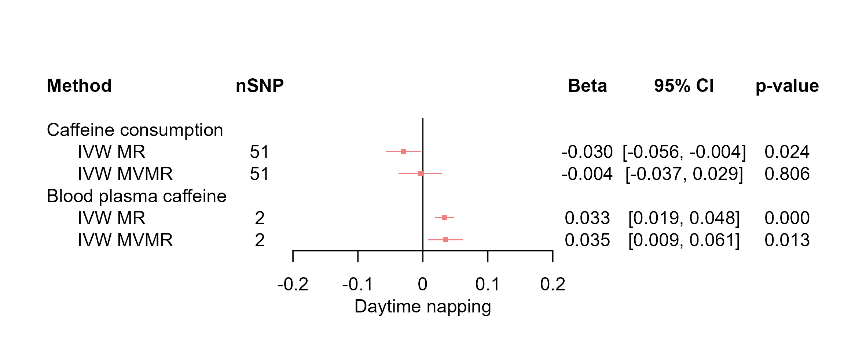

**Supplementary Figure 10b.** MR results of effects of caffeine consumption and blood plasma caffeine on daytime napping.

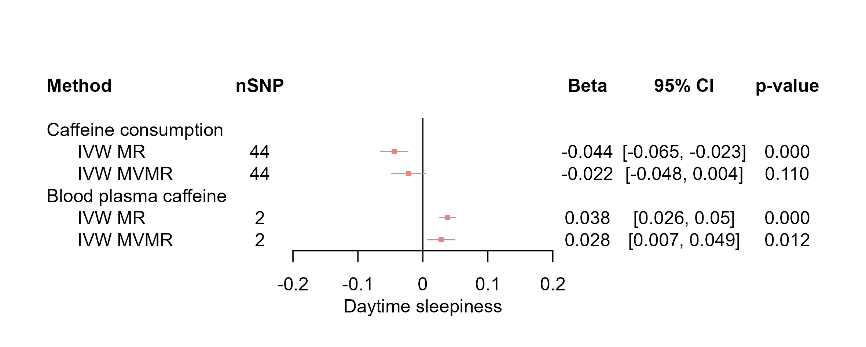

**Supplementary Figure 10c.** MR results of effects of caffeine consumption and blood plasma caffeine on daytime sleepiness.

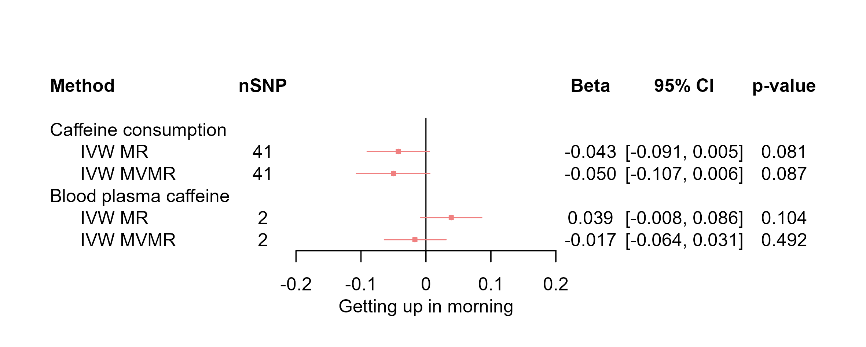

**Supplementary Figure 10d.** MR results of effects of caffeine consumption and blood plasma caffeine on getting up in morning.

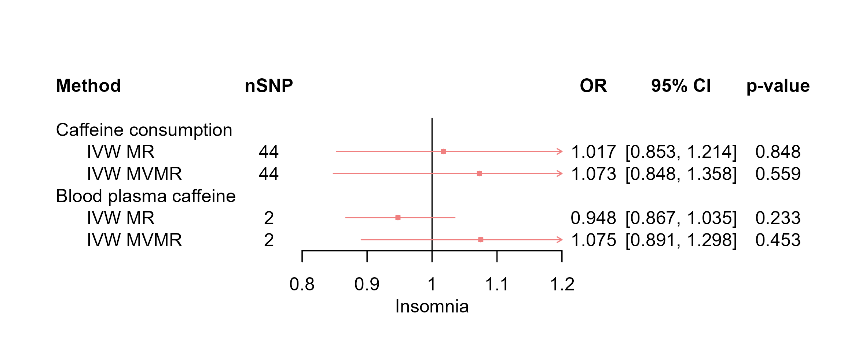

**Supplementary Figure 10e.** MR results of effects of caffeine consumption and blood plasma caffeine on insomnia.

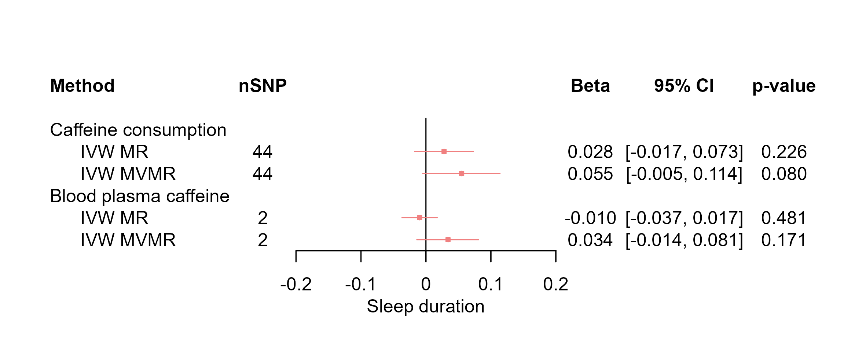

**Supplementary Figure 10f.** MR results of effects of caffeine consumption and blood plasma caffeine on sleep duration.

Red squares represent the estimated effect sizes (betas or odds ratios (ORs)) for each individual estimation method. The red horizontal lines represent the 95% confidence intervals (95% CI) for the estimated effects. The black vertical line represents the point of no effect. “nSNP” gives the number of single nucleotide polymorphisms (SNPs) used in each estimation method. “IVW MR” gives the total effects estimated using inverse variance weighted (IVW) Mendelian Randomisation (MR). “IVW MVMR” gives the direct effects estimated using Multivariable Mendelian Randomisation (MVMR).

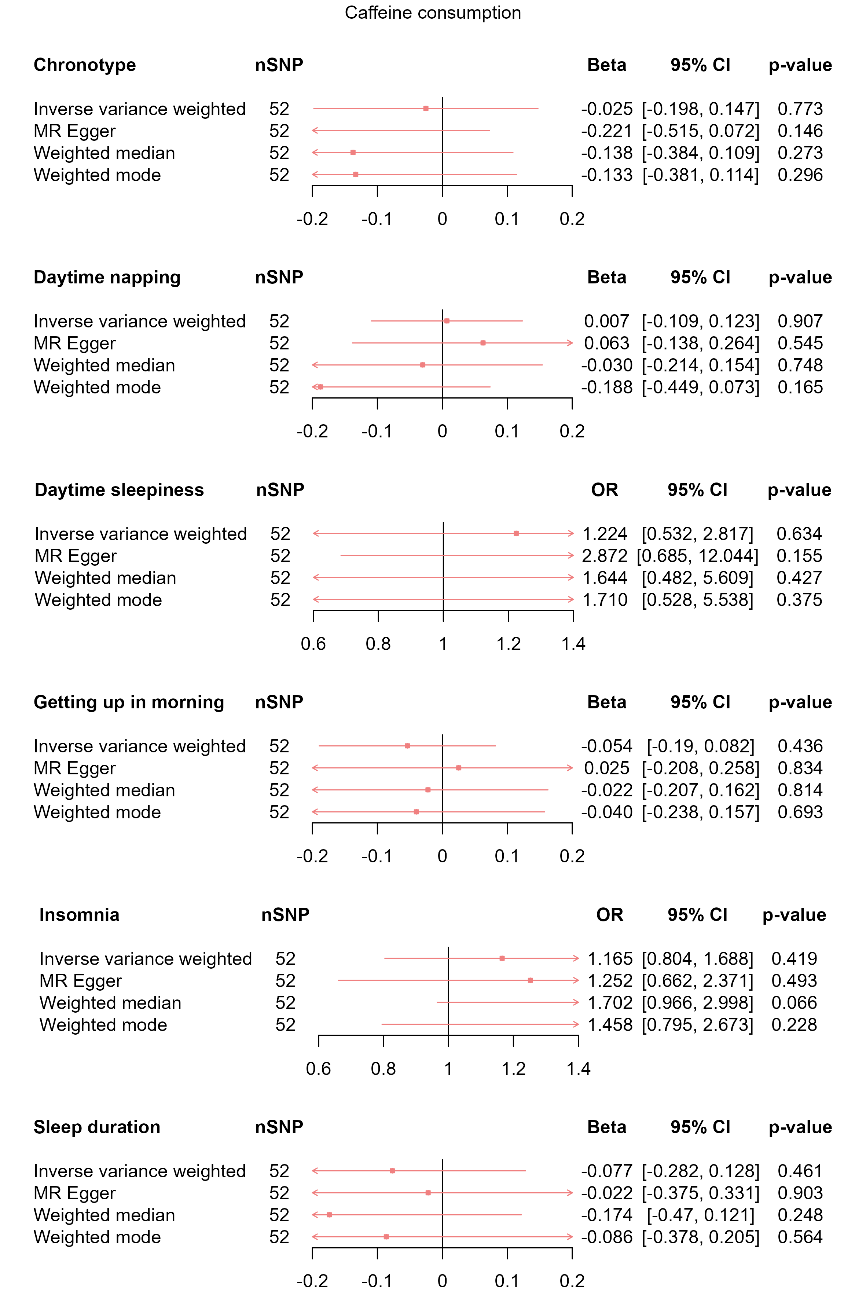

**Supplementary Figure 11a.** MR results of total effect of caffeine consumption on sleep behaviours in non-current caffeine consumers.

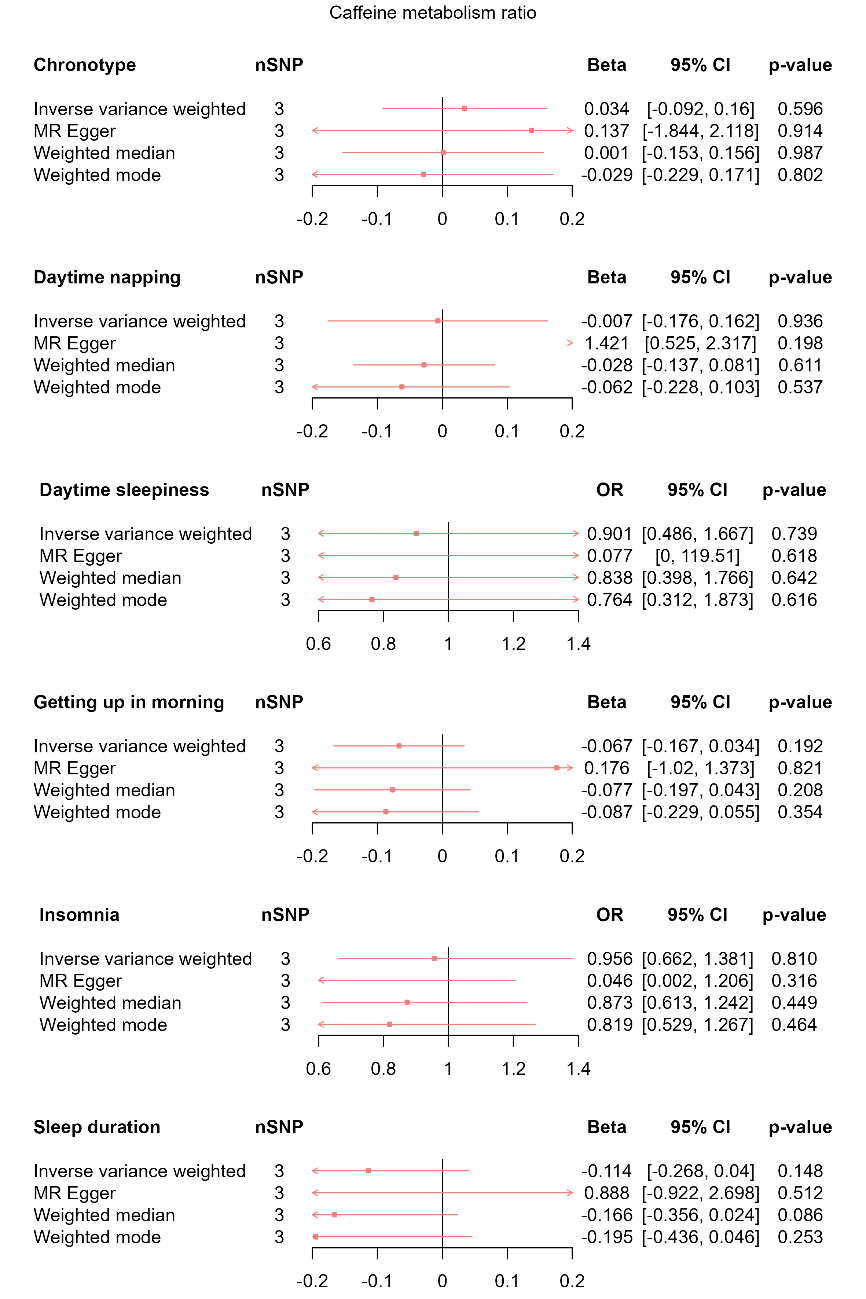

**Supplementary Figure 11b.** MR results of total effect of caffeine metabolism ratio on sleep behaviours in non-current caffeine consumers.

Red squares represent the estimated effect sizes (betas and odds ratios (ORs)) for each individual estimation method. The red horizontal lines represent the 95% confidence intervals (95% CI) for the estimated effects. The black vertical line represents the point of no effect. “nSNP” gives the number of single nucleotide polymorphisms (SNP) used in each estimation method.

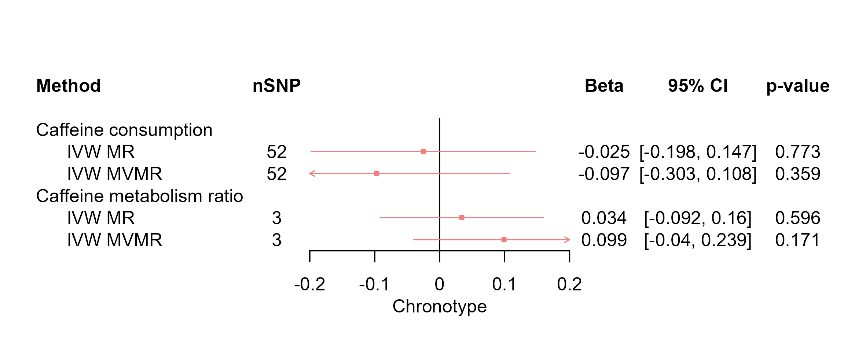

**Supplementary Figure 12a.** MR results of effects of caffeine consumption and caffeine metabolism ratio on chronotype in non-current caffeine consumers.

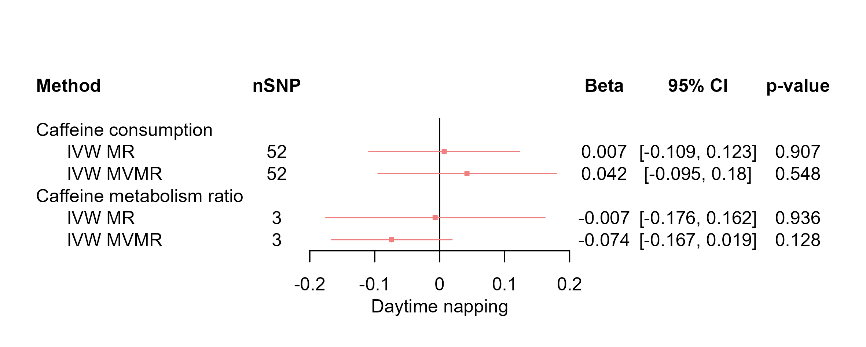

**Supplementary Figure 12b.** MR results of effects of caffeine consumption and caffeine metabolism ratio on daytime napping in non-current caffeine consumers.

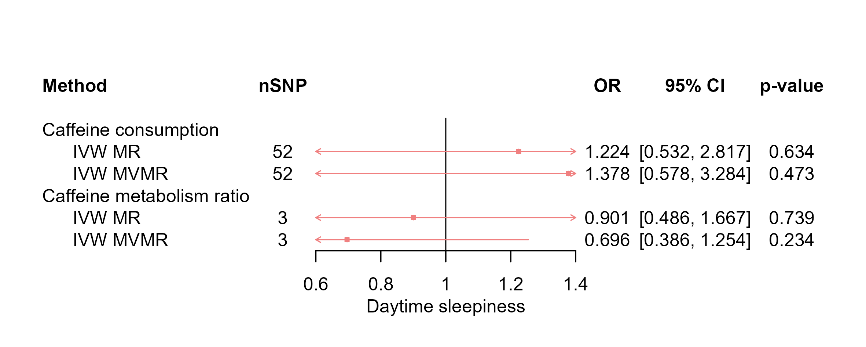

**Supplementary Figure 12c.** MR results of effects of caffeine consumption and caffeine metabolism ratio on daytime sleepiness in non-current caffeine consumers.

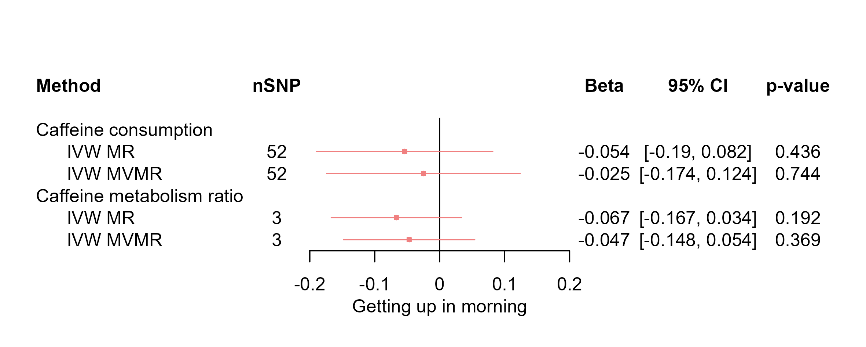

**Supplementary Figure 12d.** MR results of effects of caffeine consumption and caffeine metabolism ratio on getting up in morning in non-current caffeine consumers.

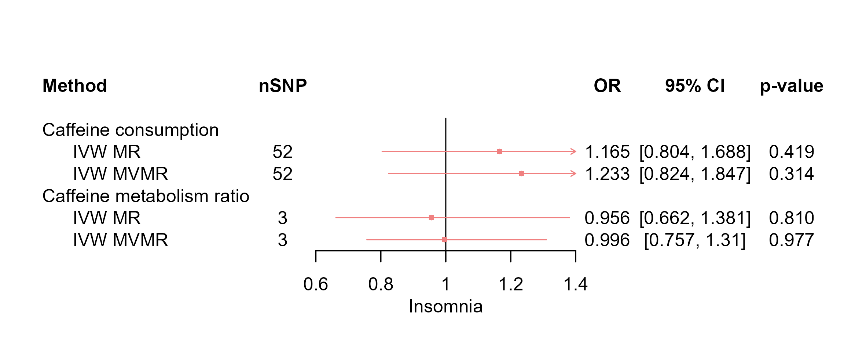

**Supplementary Figure 12e.** MR results of effects of caffeine consumption and caffeine metabolism ratio on insomnia in non-current caffeine consumers.

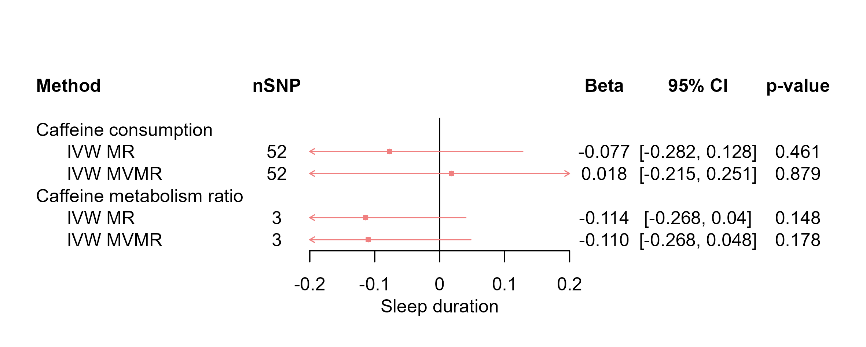

**Supplementary Figure 12f.** MR results of effects of caffeine consumption and caffeine metabolism ratio on sleep duration in non-current caffeine consumers.

Red squares represent the estimated effect sizes (betas or odds ratios (ORs)) for each individual estimation method. The red horizontal lines represent the 95% confidence intervals (95% CI) for the estimated effects. The black vertical line represents the point of no effect. “nSNP” gives the number of single nucleotide polymorphisms (SNPs) used in each estimation method. “IVW MR” gives the total effects estimated using inverse variance weighted (IVW) Mendelian Randomisation (MR). “IVW MVMR” gives the direct effects estimated using Multivariable Mendelian Randomisation (MVMR).

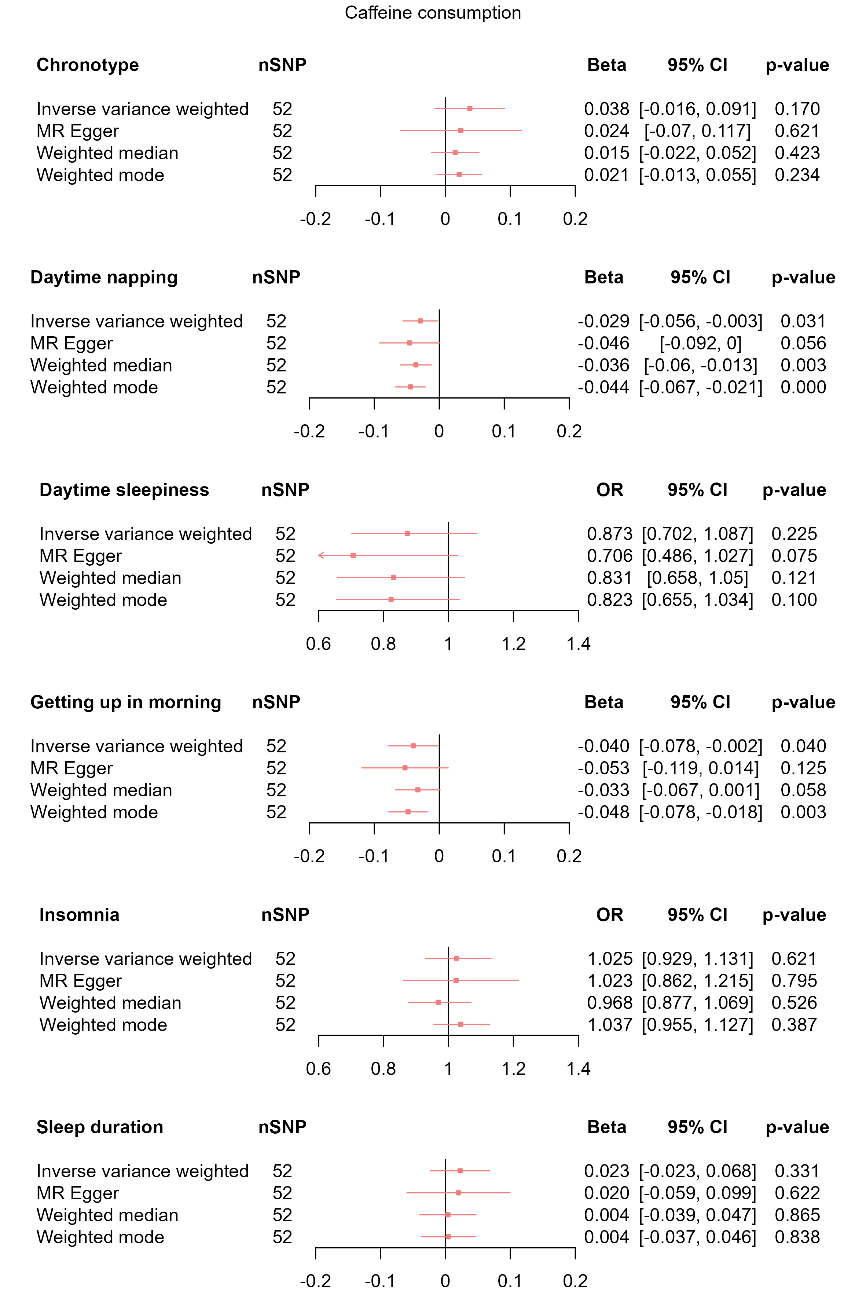

**Supplementary Figure 13a.** MR results of total effect of caffeine consumption on sleep behaviours in current caffeine consumers.

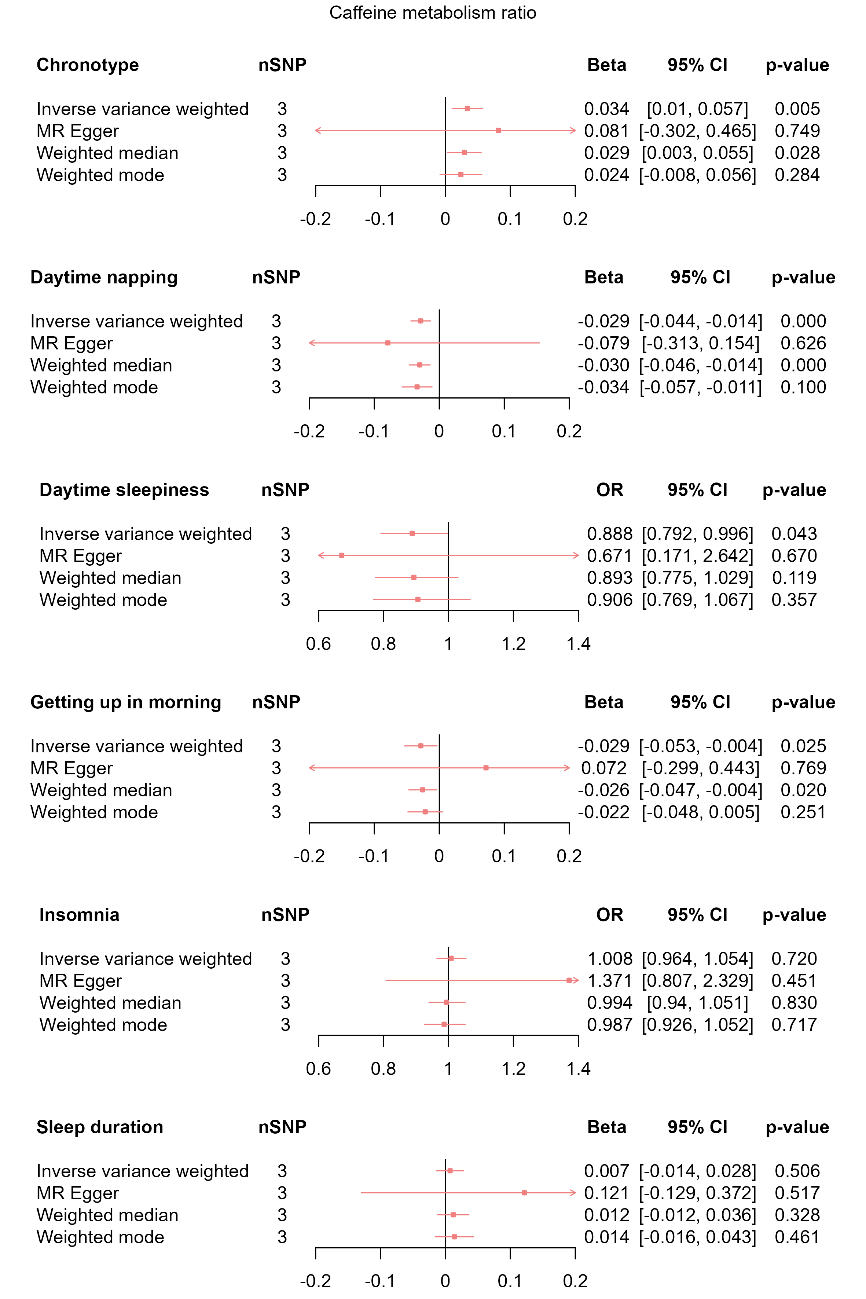

**Supplementary Figure 13b.** MR results of total effect of caffeine metabolism ratio on sleep behaviours in current caffeine consumers.

Red squares represent the estimated effect sizes (betas and odds ratios (ORs)) for each individual estimation method. The red horizontal lines represent the 95% confidence intervals (95% CI) for the estimated effects. The black vertical line represents the point of no effect. “nSNP” gives the number of single nucleotide polymorphisms (SNP) used in each estimation method.

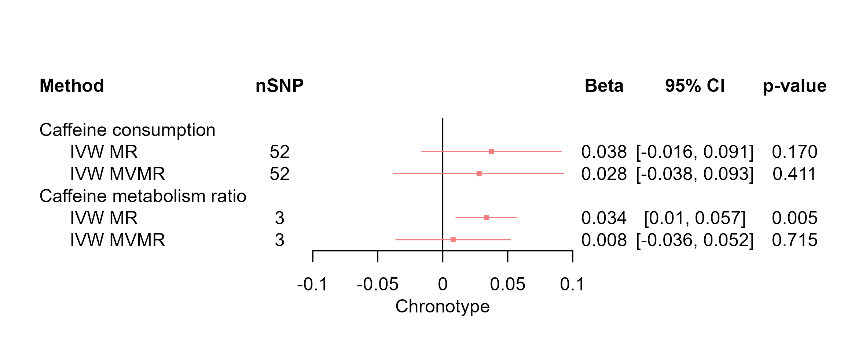

**Supplementary Figure 14a.** MR results of effects of caffeine consumption and caffeine metabolism ratio on chronotype in current caffeine consumers.

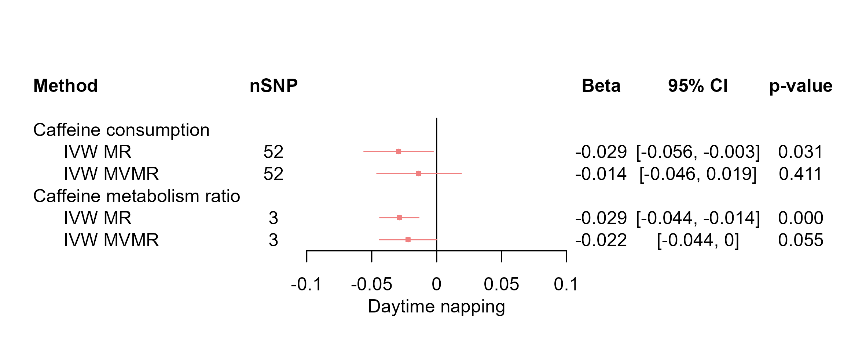

**Supplementary Figure 14b.** MR results of effects of caffeine consumption and caffeine metabolism ratio on daytime napping in current caffeine consumers.

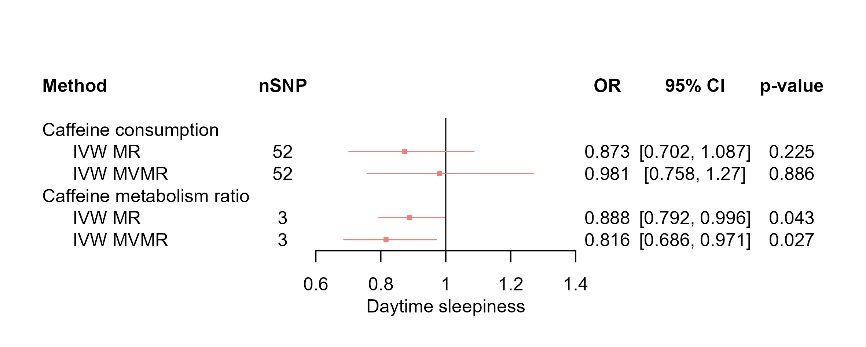

**Supplementary Figure 14c.** MR results of effects of caffeine consumption and caffeine metabolism ratio on daytime sleepiness in current caffeine consumers.

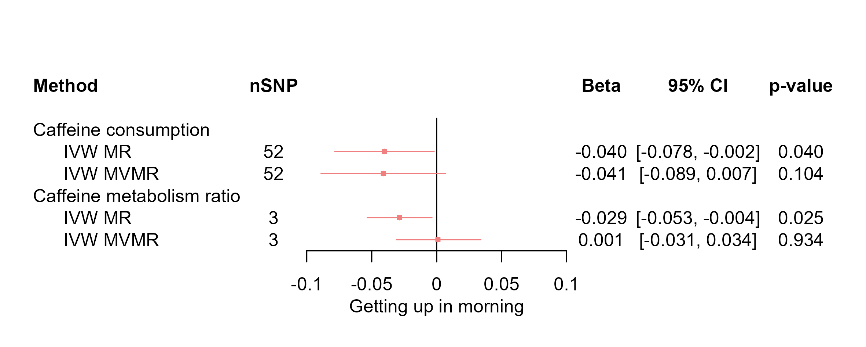

**Supplementary Figure 14d.** MR results of effects of caffeine consumption and caffeine metabolism ratio on getting up in morning in current caffeine consumers.

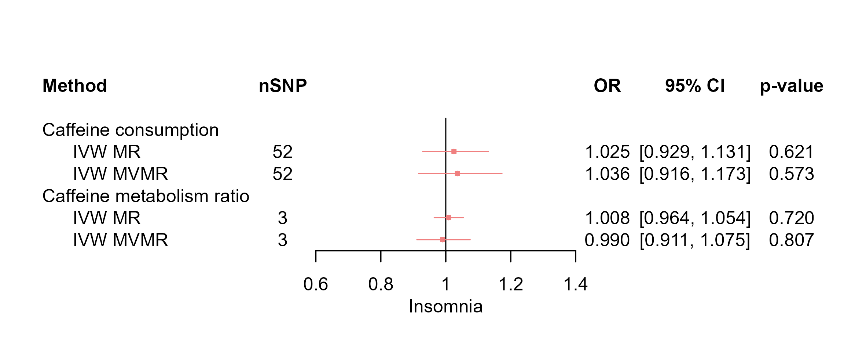

**Supplementary Figure 14e.** MR results of effects of caffeine consumption and caffeine metabolism ratio on insomnia in current caffeine consumers.

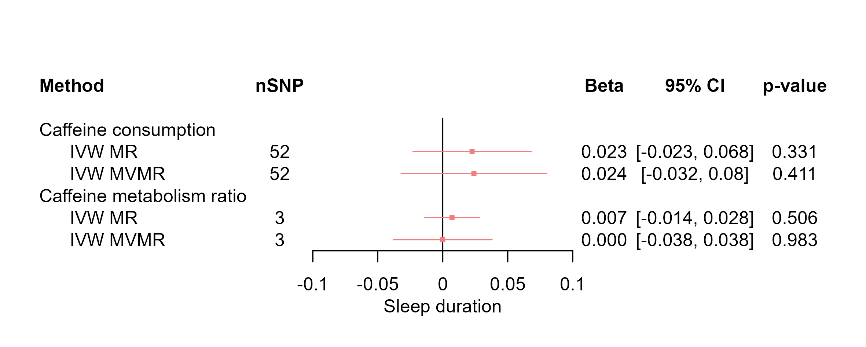

**Supplementary Figure 14f.** MR results of effects of caffeine consumption and caffeine metabolism ratio on sleep duration in current caffeine consumers.

Red squares represent the estimated effect sizes (betas or odds ratios (ORs)) for each individual estimation method. The red horizontal lines represent the 95% confidence intervals (95% CI) for the estimated effects. The black vertical line represents the point of no effect. “nSNP” gives the number of single nucleotide polymorphisms (SNPs) used in each estimation method. “IVW MR” gives the total effects estimated using inverse variance weighted (IVW) Mendelian Randomisation (MR). “IVW MVMR” gives the direct effects estimated using Multivariable Mendelian Randomisation (MVMR).

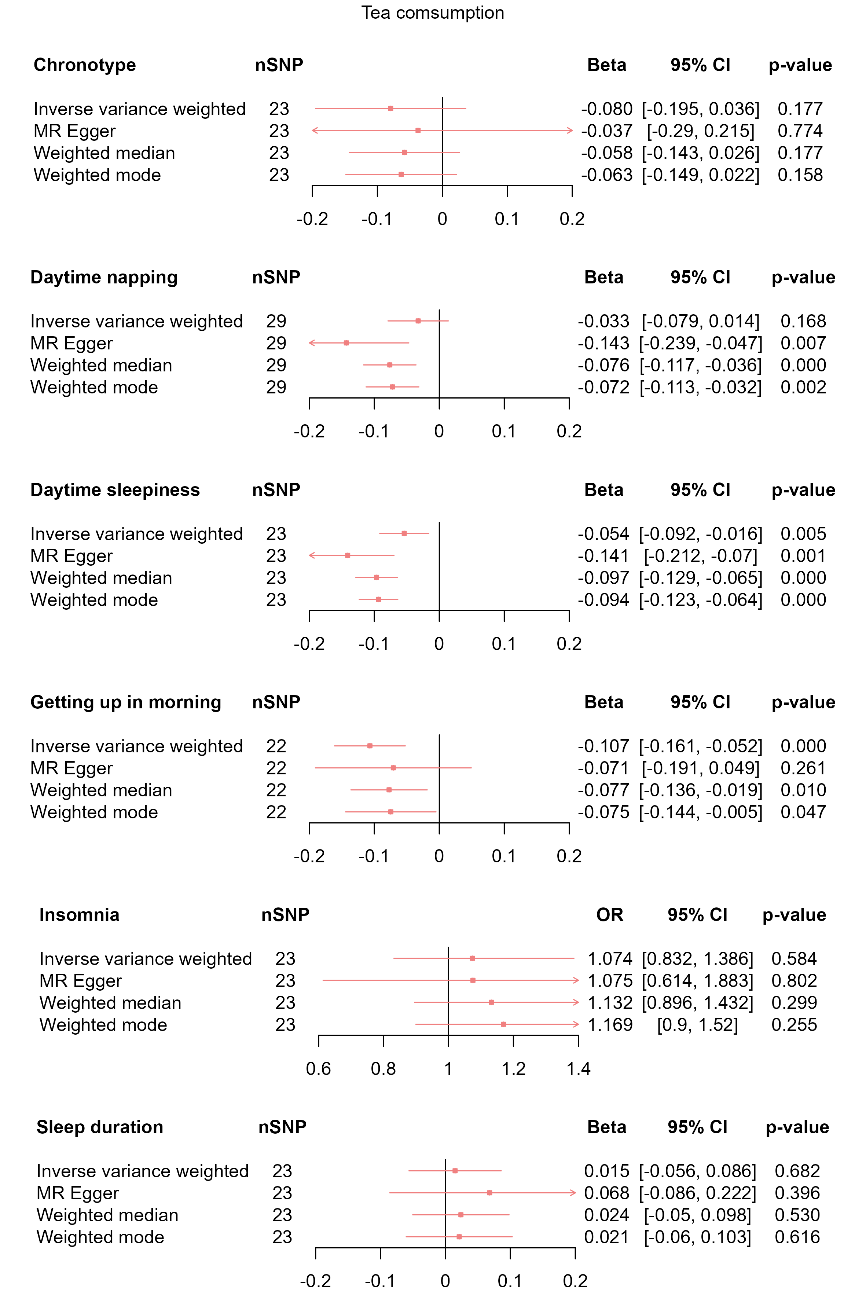

**Supplementary Figure 15a.** MR results of total effect of tea consumption on sleep behaviours.

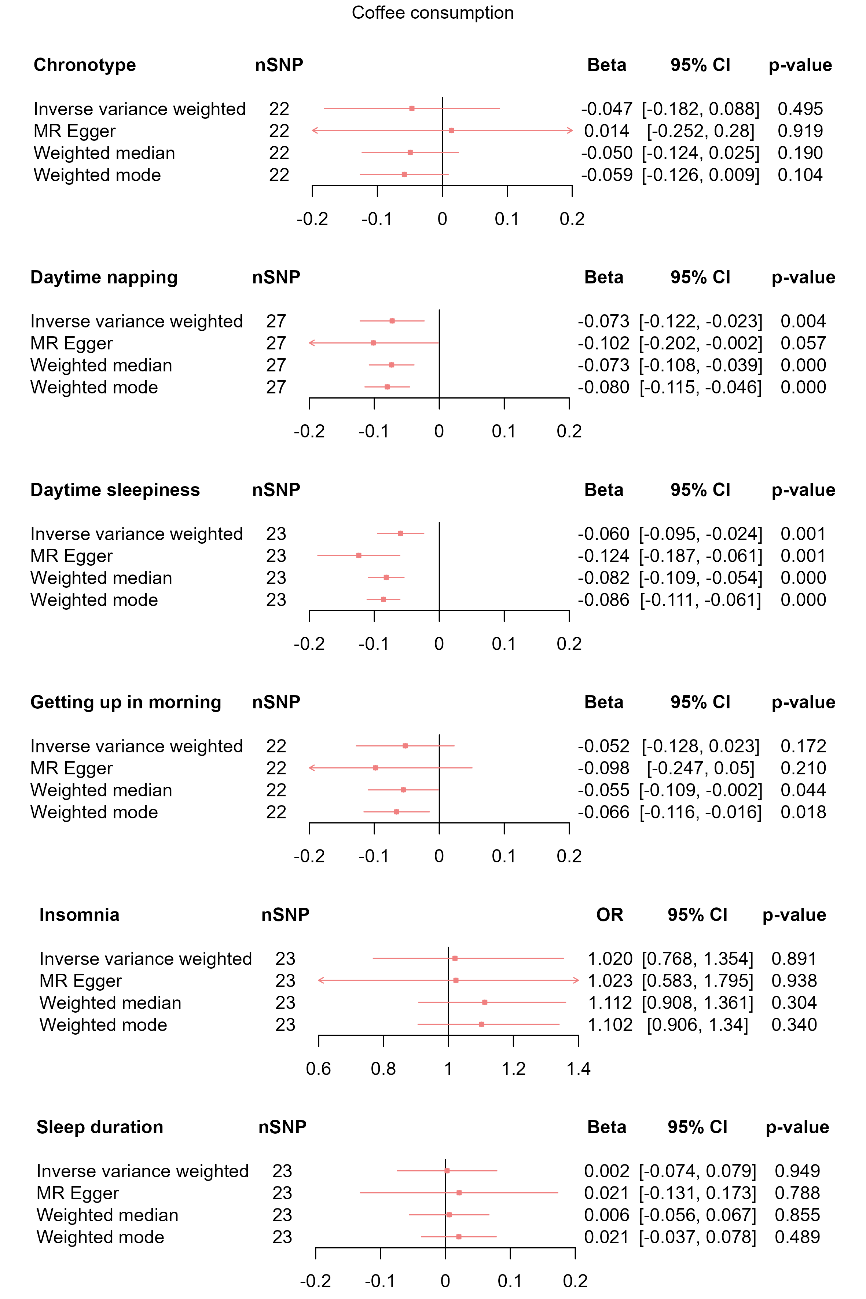

**Supplementary Figure 15b.** MR results of total effect of coffee consumption on sleep behaviours.

Red squares represent the estimated effect sizes (betas) for each individual estimation method. The red horizontal lines represent the 95% confidence intervals (95% CI) for the estimated effects. The black vertical line represents the point of no effect. “nSNP” gives the number of single nucleotide polymorphisms (SNP) used in each estimation method.

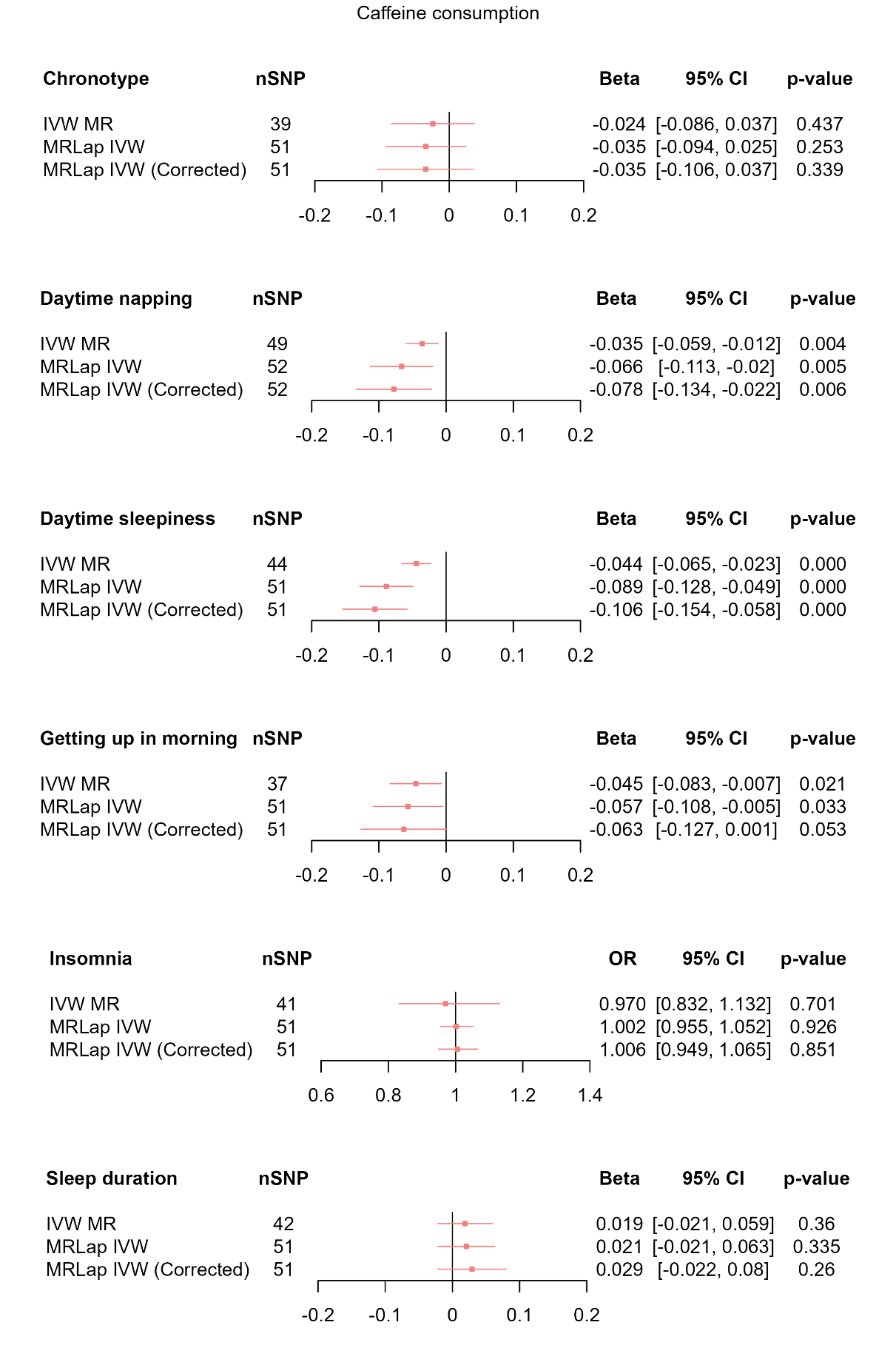

**Supplementary Figure 16.** Sample overlap-adjusted MR results of total effect of coffee consumption on sleep behaviours.

Red squares represent the estimated effect sizes (betas) for each individual estimation method. The red horizontal lines represent the 95% confidence intervals (95% CI) for the estimated effects. The black vertical line represents the point of no effect. “nSNP” gives the number of single nucleotide polymorphisms (SNP) used in each estimation method.
